# XMR: A cross-population Mendelian randomization method for causal inference using genome-wide summary statistics

**DOI:** 10.64898/2026.03.10.26348003

**Authors:** Xinrui Huang, Zitong Chao, Zhiwei Wang, Xianghong Hu, Can Yang

## Abstract

Mendelian randomization (MR) is an important tool for inferring causal relationships between exposures (like lifestyle factors or biomarkers) and health outcomes using genome-wide association study (GWAS) summary data, yet the small sample sizes of non-European populations often result in insufficient instrumental variables (IVs) and unreliable causal effect estimates. In this paper, we consider causal inference in underrepresented populations to improve global health equity. We propose a statistical method for cross-population MR, XMR, to enhance causal inference in these target populations by using auxiliary GWAS summary statistics from global biobanks. By leveraging the shared genetic basis of exposure traits in the target and auxiliary populations, XMR increases the number of IVs while maintaining robust estimates via rigorous evaluation of IV validity and accounting for confounding factors. Through extensive simulations and real-data analyses, we demonstrate that XMR can achieve greater statistical power, better control of false positive rates and more replicable results compared to existing methods. Notably, XMR successfully identifies novel causal relationships in our studies of the East Asian (including Japanese and Taiwanese), Central/South Asian, and African populations. These findings reveal potential heterogeneity in causal patterns across populations, highlighting the importance of causal inference in underrepresented populations.

## Introduction

Over the past two decades, genome-wide association studies (GWAS) have transformed our understanding of the genetic basis of complex traits and diseases [1]. To establish causal relationships between exposures and outcomes, Mendelian randomization (MR) has emerged as a crucial method for causal inference in genetic studies, using genetic variants as instrumental variables (IVs). MR analysis has been extensively applied to identify causal relationships by leveraging data from GWAS, offering new insights in both biomedical research and social sciences [2, 3].

Despite these advancements, findings are predominantly based on European populations. Although global biobanks have allowed GWAS to include diverse groups, such as East Asians [4, 5], Central/South Asians [6], Africans [7], and Hispanics [8], the majority of studies (*>* 85% as of 2024) still focus on European populations [9, 10], leaving non-European groups underrepresented. Due to ancestral differences, the causal relationships inferred in European populations may not be extrapolated to non-European populations. For instance, hypertension presents unique risks in African populations, with higher prevalence, greater severity, and more extensive organ damage at similar blood pressure levels compared to Europeans [11]. Similarly, Asian populations exhibit higher HbA1c levels, a greater prevalence of diabetes, earlier onset of the disease, and an increased risk of complications compared to Europeans [12, 13]. For cholelithiasis, the incidence and types of the disease also differ significantly between European and Asian populations [14]. Therefore, neglecting underrepresented populations may overlook genetic risk factors unique to these groups [15]. It is crucial to investigate causal relationships in these populations to address existing healthcare disparities.

To explore causal relationships in the underrepresented populations, existing MR methods, such as Inverse Variance Weighted regression (IVW) [16, 17], Egger [18], Weighted-median [19], Weighted-mode [20], MRAPSS [21], and CAUSE [22], face significant challenges. One major issue is the limited sample sizes, which complicate IV selection based on GWAS results from these populations. Using a stringent threshold for IV selection (e.g., *P* value = 5 *×* 10^*−*8^) may yield too few IVs for reliable causal inference, while a more lenient threshold (e.g., *P* value = 5 *×* 10^*−*5^) can lead to the inclusion of many invalid IVs, resulting in inflated false-positive findings. To address these challenges, two MR methods—TEMR [23] and MR-EILLS [24]—have been proposed for causal inference in cross-population settings. TEMR seeks to enhance statistical power for small-sample populations by leveraging cross-ancestry genetic correlations. However, it is prone to inflated type I errors because it does not adequately account for confounding factors, such as correlated pleiotropy and sample structure, which may be hidden in GWAS summary statistics. On the other hand, MR-EILLS explores causal relationships under the assumption that the causal effect is consistent across populations. This assumption can be overly simplistic and risks overlooking important differences among populations, limiting its applicability for understanding population-specific causal effects.

In this paper, we introduce XMR, a cross-population MR method aimed at enhancing causal inference for underrepresented populations by utilizing large-sample GWAS data. The effectiveness of XMR is based on several key components. First, XMR does not assume that causal effects are invariant across populations, enabling it to identify both population-shared and population-specific effects in an unbiased manner. Second, XMR allows for the selection of IVs based on extensive European exposure data, effectively addressing the challenge of insufficient IVs that traditional MR methods encounter in small-sample populations. Importantly, XMR does not presume that the effect sizes of IVs are identical across populations, which enables it to automatically assess the validity of each IV and include only those that are valid for causal inference. Third, XMR accounts for selection bias introduced by IV selection and linkage disequilibrium (LD) clumping, as well as confounding factors such as pleiotropy and sample structure. This is achieved through its probabilistic framework, resulting in reliable causal effect estimates accompanied by well-calibrated *P* values.

Through a comprehensive simulation study, we demonstrate that XMR achieves superior statistical power and estimation accuracy compared to existing methods, while maintaining well-calibrated *P* values. In real-data negative control studies, XMR effectively controls type I errors, highlighting the effectiveness of its various model components. We then apply XMR to three underrepresented populations—East Asians, Central/South Asians, and Africans—to uncover novel causal relationships that conventional MR methods have overlooked. For East Asians, we systematically evaluate causal inference results using two independent cohorts: BioBank Japan (BBJ) [4] and the Taiwan Precision Medicine Initiative (TPMI) [25, 26]. Our findings show that XMR outperforms related MR methods by providing consistent causal effect estimates that are validated by both BBJ and TPMI. Finally, we compare causal effects estimated from non-European populations with those from European populations. XMR not only identifies causal effects that are shared across populations but also highlights population-specific causal relationships. We believe these findings are vital for advancing global healthcare equity.

## Results

### Method overview

Conventional MR methods face significant challenges in IV selection when performing causal inference in underrepresented populations due to limited sample sizes. A stringent threshold for IV selection may yield only a small number of IVs, resulting in unreliable causal inferences. Conversely, a loose threshold can include many invalid IVs, leading to inflated false-positive rates. To address these issues, we propose XMR, a probabilistic model for cross-population MR analysis. XMR leverages large-sample GWAS data from European (EUR) populations as an auxiliary resource to enhance IV selection and improve the statistical power of causal inference (Fig. 1A), while also accounting for differences between the target and auxiliary populations to effectively control type I errors.

**Figure 1.**
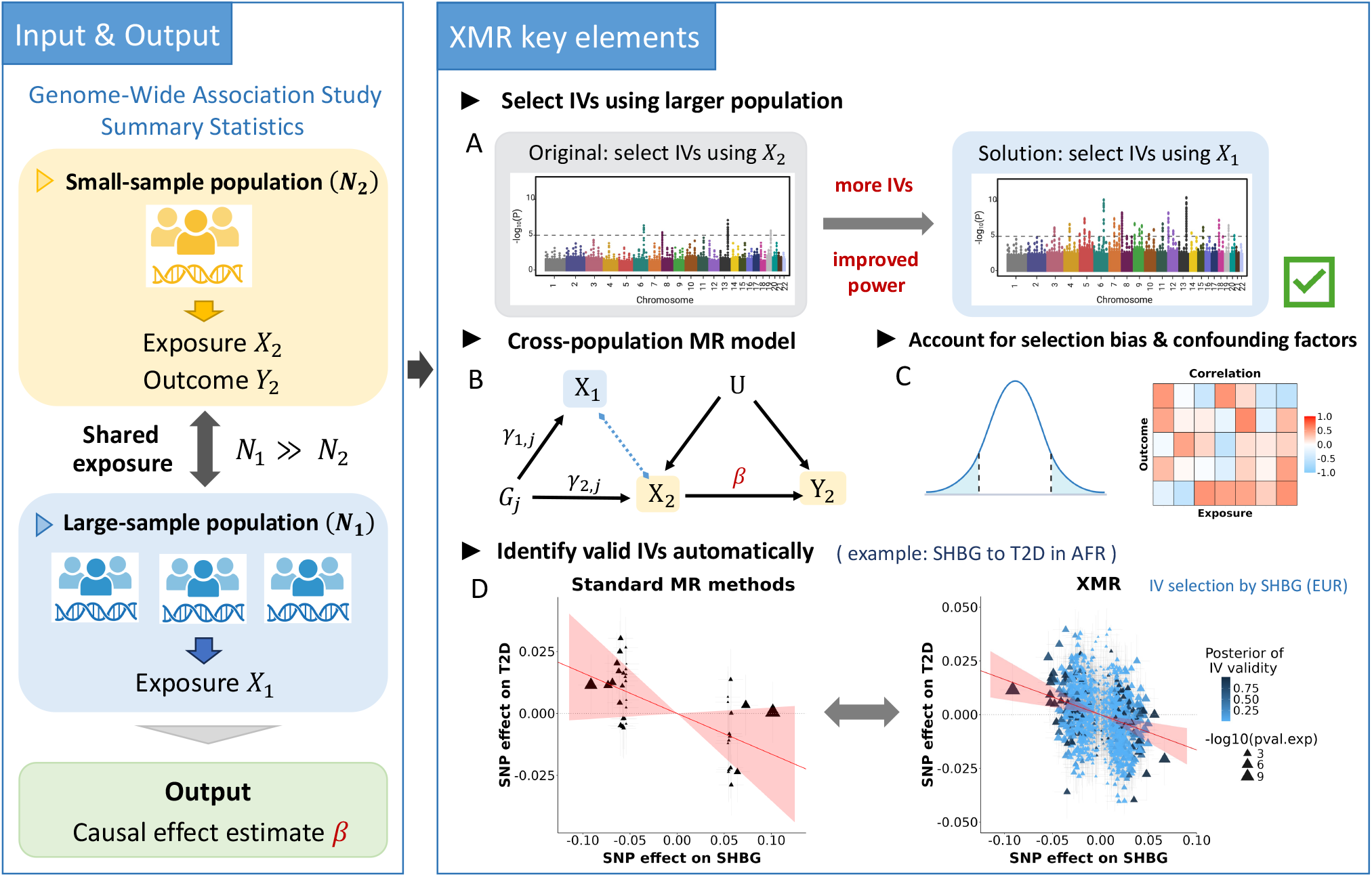
Overview of the XMR method. XMR estimates the causal effect *β* between exposure *X*_2_ and outcome *Y*_2_ in a small-sample population by leveraging data on the same exposure *X*_1_ from a large-sample population. The method involves several key elements: **(A)** IVs are selected from the large-sample population (*X*_1_) to improve power compared to the limited IVs available from the small-sample population (*X*_2_). The distributions of observed *−* log_10_(*p*) values for SNP-exposure associations across chromosomes are shown. **(B)** The XMR model diagram. The arrowed lines represent effects with directions. The blue dashed line indicates the correlation between *X*_1_ and *X*_2_. **(C)** Selection bias and confounding factors contribute to observed SNP-trait associations. **(D)** An illustrative example of causal inference between SHBG (sex hormone-binding globulin) and T2D (type 2 diabetes) in an African population, using conventional two-sample MR methods (left) and XMR (right), respectively. The estimated causal effect is shown as a red line, with the 95% confidence interval (CI) shaded in transparent red. Triangles represent observed SNP effect sizes (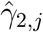 and 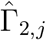), colored by their posterior probability of IV validity (*Z*_*j*_ = 1 in dark blue; *Z*_*j*_ = 0 in light blue). Figure 1 created with BioRender.com, with permission.

Suppose we aim to estimate the causal effect *β* between an exposure *X*_2_ and an outcome *Y*_2_ in a target population with sample size *N*_2_. Let *X*_1_ be the exposure from an auxiliary population which has a larger sample size *N*_1_, where *N*_1_ *≫ N*_2_. Unlike conventional MR methods which perform IV selection solely based on the target population, XMR selects IVs from the auxiliary population to increase the number of available IVs, and thus improve statistical power for causal inference (Fig. 1A, Fig. 1B). Specifically, we select IVs based on the criterion 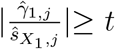, where 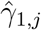 denotes the estimated effect size of the *j*-th SNP on *X*_1_ with its standard error 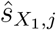 in the auxiliary population, and the threshold *t* is a predefined number to ensure that selected IVs are relevant to the exposure of interest. We also perform LD clumping to minimize dependence among the selected IVs, ensuring that the remaining IVs are nearly independent.

Let *M*_*t*_ be the remaining number of IVs for our probabilistic model. XMR takes GWAS summary statistics 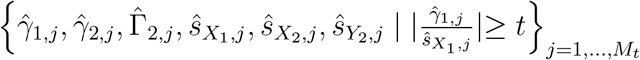 as its input for causal inference, where 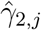 and 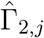 denote the estimated effect sizes of the *j*-th SNP on *X*_2_ and *Y*_2_ with their standard errors 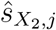 and 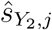. The probabilistic model of XMR is designed to address several key issues. First, while the selection of IVs from the auxiliary population increases the total number, a portion of these IVs may still be invalid. XMR effectively distinguishes valid IVs from all selected candidates and utilizes only the valid IVs for causal inference to avoid false positives. Second, it is critical to characterize the correlation between the target population and the auxiliary population. With this relationship, we are able to correct the selection bias induced during IV selection and LD clumping (Fig. 1C). Third, confounding factors—such as correlated pleiotropy, population stratification, and cryptic relatedness—often remain hidden in the GWAS summary statistics from both the target and auxiliary populations (Fig. 1C). It is crucial to account for these confounding factors in a cross-population context. By integrating these elements, XMR effectively expands the pool of valid IVs, resulting in more accurate estimates, narrower confidence intervals, and increased statistical power (Fig. 1D). Further details can be found in Methods.

### Experimental design

We compared XMR with one cross-population MR method, TEMR, and 14 conventional two-sample MR methods, including MRAPSS, Egger, Weighted-mode, Weighted-median, CAUSE, IVW (random, denoted as IVW) [16], IVW (fixed, denoted as IVW-fe) [17], dIVW [27], MRMix [28], ConMix [29], MR-PRESSO [30], cML-MA (without data perturbation) [31], MR-Robust [32] and MR-Lasso [32] (Fig. 2A). The preprocessing steps for GWAS summary statistics prior to MR analysis are outlined in Fig. 2B, including SNP quality control, aligning SNP effects of the exposure and outcome to the same allele, IV selection and LD clumping. Specifically, we examined the performance of XMR and MRAPSS using their default IV threshold of 5 *×* 10^*−*5^, and TEMR at its default threshold of 5 *×* 10^*−*8^. CAUSE became unstable in scenarios with small sample sizes under the recommended IV threshold of 1 *×* 10^*−*3^. To address this, we adopted a threshold of 5 *×* 10^*−*5^, which offered comparable but more stable performance (see discussion in Supplementary section 4.9). Other methods, designed for a stringent IV threshold of 5 *×* 10^*−*8^ to retain strong IVs, struggled with insufficient IVs in some cases. Therefore, we used 5 *×* 10^*−*8^ when the exposure sample size exceeded 20,000 and 5 *×* 10^*−*5^ otherwise. LD clumping was performed for all methods using the LD panel of the target population to obtain independent IVs, with *r*^2^ *<* 0.001. At least 4 SNPs were required to run each MR method. A more detailed description is given in Supplementary sections 2.3.1 and 2.3.2.

**Figure 2.**
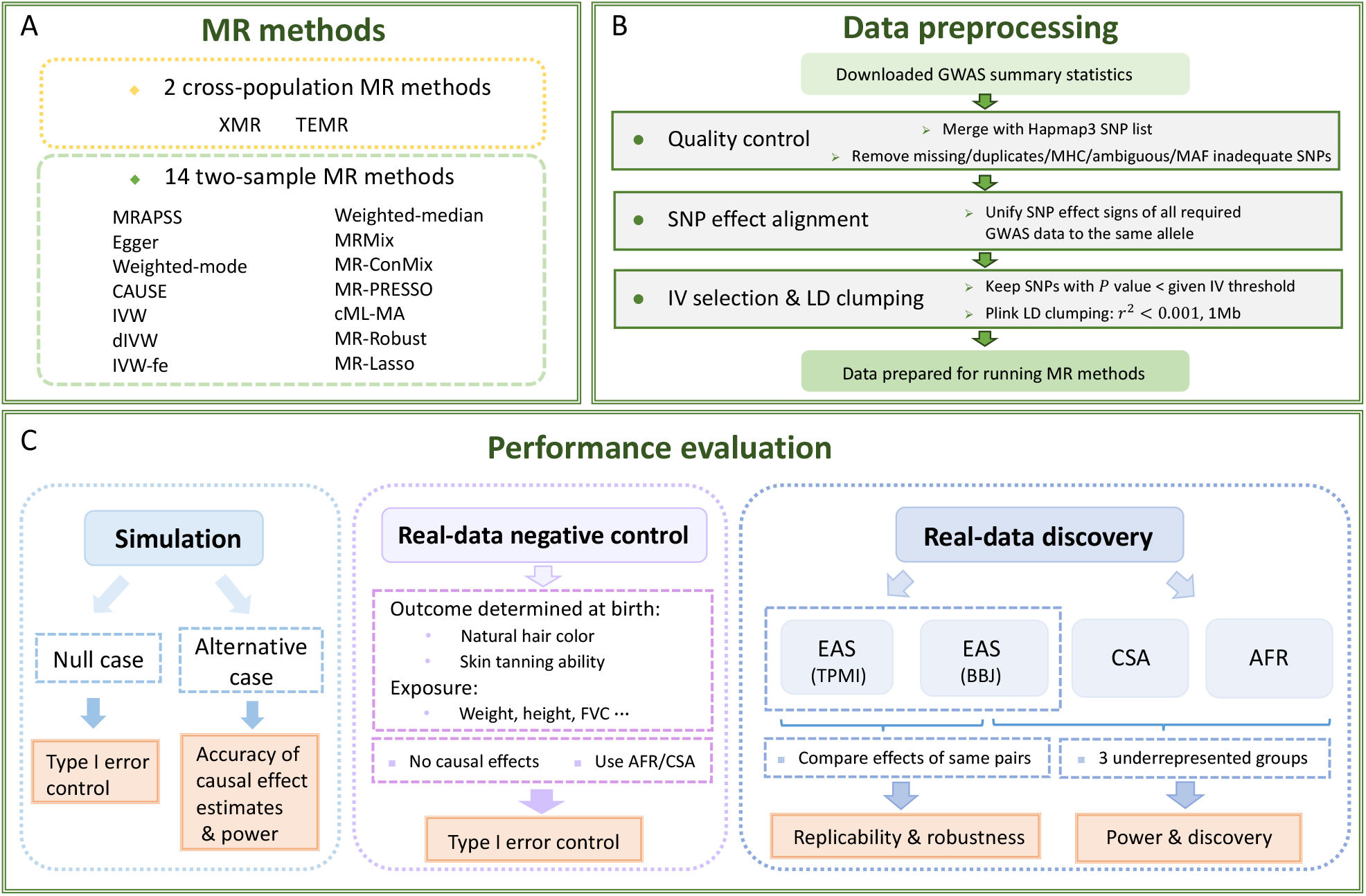
Experimental design. Pipeline for an overall evaluation of MR methods. **(A)** 16 MR methods are included in the analyses. **(B)** A three-step procedure to prepare data for running MR methods. **(C)** Performance evaluation regarding type I error control, accuracy of causal effect estimates, replicability, and power in three scenarios: simulation, real-data negative control in AFR and CSA, and real-data discovery in three underrepresented populations: EAS, CSA, AFR.

Our analysis comprised three parts to evaluate different aspects of MR methods (Fig. 2C). In simulations, we generated summary statistics for a large-sample population (*N*_1_ = 80, 000) and a small-sample population (*N*_2_ = 15, 000) under varying genetic correlations, in the presence of pleiotropy and sample structure. Type I error control of each method was assessed in null cases (*β* = 0), while scenarios with *β*≠0 were used to evaluate causal effect estimation accuracy and statistical power.

To ensure a realistic evaluation of type I error control, we further conducted a negative control study utilizing real-world datasets. We selected natural hair color before greying and skin tanning ability as negative control outcomes, with exposure being various anthropometric and physiological traits (e.g., weight, height, and forced vital capacity (FVC)). The outcome traits are primarily determined at birth and unlikely to be influenced by these exposures measured later in life. By design, no causal relationship exists between the exposure and outcome traits. Our analysis included 70 trait pairs between 35 exposures and 2 negative control outcomes, with data sourced from the Pan-UKBB repository [33] (https://pan.ukbb.broadinstitute.org/). Experiments were conducted in African and Central/South Asian populations, with sample sizes of approximately 6,000 and 8,000, respectively. The European cohort, consisting of over 400,000 individuals, served as the auxiliary large-sample population for XMR and TEMR. Details about the traits and datasets used in the negative control study can be found in Supplementary Table S4.

In the last part, we applied MR methods to three underrepresented groups—East Asian (EAS), Central/South Asian (CSA), and African (AFR) populations—to compare their statistical power and discoveries. For EAS, we analyzed 22 overlapping traits from BBJ and TPMI, including 7 exposures (low-density lipoprotein cholesterol (LDLC), hemoglobin (Hb), HbA1c, diastolic blood pressure (DBP), body mass index (BMI), alanine aminotransferase (ALT), and glucose (Glc)) and 15 disease-related outcomes. Exposure GWAS sample sizes ranged from 71,221 to 163,835 for BBJ and 90,509 to 203,766 for TPMI, while outcome sample sizes ranged from 79,550 to 178,726 for BBJ and 117,944 to 334,322 for TPMI. Auxiliary EUR GWAS data were sourced from the Neale Lab repository (http://www.nealelab.is/uk-biobank), UK Biobank [34], and FinnGen [35], with typical sample sizes of ∼350,000 for exposures and ∼450,000 for outcomes. The replicability and robustness of methods were assessed by comparing causal effect estimates using these two different EAS datasets.

In the CSA and AFR populations, we investigated the causal interplay among five high-burden diseases (type 2 diabetes (T2D), hypertension (HTN), chronic ischaemic heart disease (CIHD), disorders of lipid metabolism (DLM) and asthma), and diverse biomedical traits. To guarantee robust instrument strength, we restricted our dataset to traits with at least four valid IVs. We filtered out traits whose sample sizes were too limited for stable estimation of confounding factor parameters via LD Score Regression (LDSC), as well as traits with few causal signals. As a result, we covered 11 biomedical traits for CSA (BMI, height, HbA1c, LDLC, high-density lipoprotein cholesterol (HDLC), triglycerides (TG), neutrophil count (Neutro), C-reactive protein (CRP), insulin-like growth factor 1 (IGF-1), sex hormone-binding globulin (SHBG) and vitamin D (Vit D)), while we examined 8 biomedical traits (LDLC, TG, Neutro, CRP, IGF-1, SHBG, Vit D and albumin (ALB)) for AFR. Most datasets for CSA, AFR, and the auxiliary EUR cohort were sourced from Pan-UKBB, with sample sizes of ∼8,000 for CSA, ∼6,000 for AFR, and ∼400,000 for EUR. Albumin data for AFR was obtained from a pan-African GWAS [36], with a sample size of ∼13,000. We investigated every trait pair, excluding biomedical traits paired with other biomedical traits.

The discoveries made by each MR method reflect their statistical power and can be compared to findings in Europeans, revealing both shared and population-specific causal relationships. Detailed information about the traits and datasets used in this study can be found in Supplementary Tables S15, S20 and S23.

### XMR demonstrates robust type I error control and high power in simulations

To evaluate the performance and robustness of summary-level MR methods, we conducted simulation studies based on the XMR model framework, with some model misspecifications such as inconsistencies in the validity of IVs across two populations.

Specifically, we generated simulated summary statistics 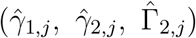 for 50,000 SNPs under the assumptions of the XMR model described in Eq. (1). However, correlated pleiotropy was only assumed for 30,000 SNPs, while all SNPs were influenced by sample structure. To mimic practical scenarios, we set the sample sizes of the large and small populations as *N*_1_ = 80, 000 and *N*_2_ = 15, 000, respectively. The variances of the sample structure terms (*ϵ*_1,*j*_, *ϵ*_2,*j*_, *ξ*_2,*j*_) were adjusted according to the sample sizes, with standard errors 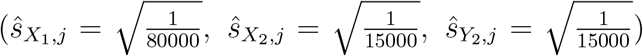 and sample structure parameters (*c*_1_ = 1.25, *c*_2_ = 1.05, *c*_3_ = 1.02). This reflects the higher uncertainty but reduced susceptibility to sample structure in the smaller population. The polygenic effects (*u*_1,*j*_, *u*_2,*j*_, *v*_2,*j*_) were simulated with variances 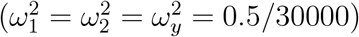, corresponding to heritabilities of 0.5 for *X*_1_, *X*_2_ and *Y*_2_. Among the 30,000 SNPs with correlated pleiotropy, only 1,500 were considered valid IVs (*Z*_*j*_ = 1) in at least one population. To ensure sufficient instrument strength, we set 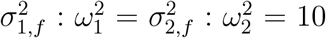, indicating that the true effects (*γ*_1,*j*_, *γ*_2,*j*_) were much stronger than confounding effects. In contrast, the direct effects *α*_*j*_ had the same magnitude as the polygenic effects, with 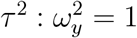. To test the robustness of each method in the presence of invalid IVs, we randomly selected 400 SNPs out of the 1,500 to introduce inconsistencies in IV validity between two populations: some SNPs were valid only in the small population (*γ*_1,*j*_ = 0), while others were valid only in the large population (*γ*_2,*j*_ = 0). Simulations were conducted for various causal effect sizes *β* under different levels of correlation *σ*_12,*f*_ */*(*σ*_1,*f*_ *σ*_2,*f*_ ) between the true effects *γ*_1,*j*_ and *γ*_2,*j*_. The correlation level reflects the degree to which information can be borrowed from the large population to improve estimation in the small population.

We compared XMR with the cross-population MR method, TEMR, and 14 single-population MR methods. Single-population methods utilized only the simulated summary statistics 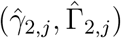 from the target population. However, TEMR also required paired outcome summary statistics from the large-sample population. For TEMR, we extended our simulation framework to generate these statistics under the assumption that the existence of causal effects was consistent across populations, a setting favorable to TEMR. Details of the simulation design are provided in Supplementary sections 2.1 and 2.2. As *N*_2_ *<* 20,000, we applied a unified criterion (*P* ≤ 5 *×* 10^*−*5^) for IV selection across most methods, except for TEMR (*P* ≤ 5 *×* 10^*−*8^).

We first evaluated the false positive rates of all methods under the null hypothesis (*β* = 0) where no causal relationship exists. Figs. 3A, B and C present quantile-quantile (QQ) plots for each method under correlations of 0, 0.3 or 0.7 based on 30 repeated experiments. In the upper panel, XMR consistently produced well-calibrated *P* values across all correlation settings, while other methods exhibited noticeable inflation. Among them, TEMR showed the most severe inflation, which might result from inadequate accounting for invalid IVs and confounding factors. Additional results for other MR methods are available in Supplementary section 4.1, Fig. S2, which similarly demonstrate inflated *P* values. The lower panel displays the results of some single-population MR methods without overly inflated *P* values across three settings, including Egger, MRAPSS and Weighted-mode. CAUSE exhibited certain levels of deflation in these experiments.

**Figure 3.**
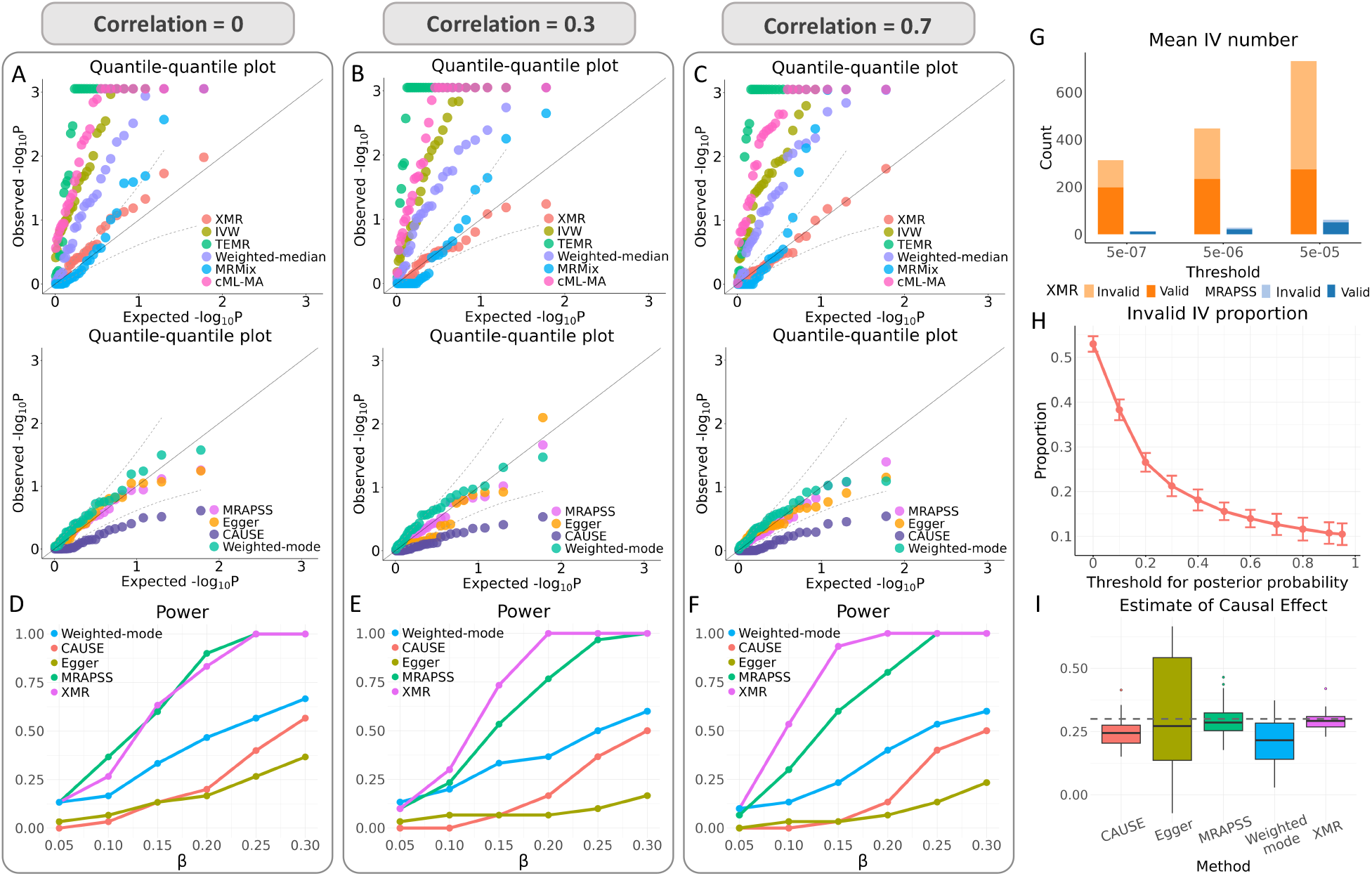
Simulation studies. Comparison of 10 summary-level MR methods using simulated data. **(A–C)** QQ plots of *−*log_10_(*p*) values from each method under the null scenario (*β* = 0) where no causal effect exists, with the correlation parameter *σ*_12,*f*_ */*(*σ*_1,*f*_ *σ*_2,*f*_ ) between two populations set to 0, 0.3 and 0.7, respectively. **(D–F)** Statistical power comparison of methods without inflated *P* values under alternative simulations with different causal effect sizes *β*. The correlation parameter still takes values of 0, 0.3, and 0.7. **(G)** Comparison of the numbers of valid and invalid IV identified by XMR and MRAPSS across various IV selection thresholds with a correlation parameter of 0.3 and *β* = 0.2. **(H)** Proportion of invalid IVs among those classified as valid by XMR, calculated for different posterior probability thresholds under the null case (*β* = 0) with a correlation parameter of 0.3. **(I)** Causal effect estimates *β* obtained by different methods under *β* = 0.3 and a correlation parameter of 0.7.

The superior performance of XMR can be attributed to its ability to identify and exclude invalid IVs during causal effect estimation. Our model assigns a posterior probability to each SNP *j* indicating its validity. By setting a threshold for the posterior probability, SNPs with values exceeding the threshold are considered to be recognized as valid by our model. Fig. 3H illustrates the error rates in inferring IV validity by summarizing the proportion of truly invalid IVs among those recognized as valid by XMR, based on posterior probability thresholds. The result at the threshold of 0 indicates that over 50% of the included IVs were invalid right after initial IV selection. However, as the threshold increased, the proportion of invalid IVs decreased sharply, dropping to approximately 15% at a threshold of 0.5 and below 10% with stricter criteria. These results underscore XMR’s ability to effectively filter out invalid IVs and minimize their influence, ensuring more accurate results. The adjustment of confounding factors and selection bias is also critical. The ablated versions of XMR, which assume no correlated pleiotropy (**Ω** = 0), no sample structure (***C*** = *I*), or no correction of selection bias, all failed to consistently control false positive rates under three settings (see Supplementary section 4.2, Fig. S3).

Next, we assessed the statistical power of methods that had no inflation under the previous null scenario. Five methods were selected for comparison: XMR, MRAPSS, Egger, CAUSE, and Weighted-mode. Simulations were conducted under correlations of 0, 0.3, and 0.7, with causal effect sizes *β* ranging from 0.05 to 0.3. As shown in Figs. 3D, E and F, XMR generally exhibited the highest power among all methods. Specifically, XMR reliably outperformed others in most scenarios, while maintaining competitive performance similar to MRAPSS in settings with zero correlation between the two populations (Fig. 3D). While the power of the other four single-population methods remained largely unaffected by correlation levels, XMR’s performance improved significantly with higher correlations. This behavior aligns with expectations, as stronger correlations indicate greater consistency in genetic effects on the exposure across populations, enabling XMR to better leverage information from the large population. MRAPSS demonstrated the second-best performance overall, benefiting from its robust design. However, XMR’s cross-population framework allowed it to surpass MRAPSS by incorporating a larger pool of candidate IVs from the auxiliary population. Fig. 3G compares the number of valid and total IVs identified by XMR and MRAPSS under *β* = 0.2 and a correlation of 0.3. Regardless of the IV selection threshold, XMR identified more valid IVs and more total IVs than MRAPSS, highlighting its advantage. Furthermore, the number of valid IVs increased steadily as the selection threshold was relaxed, further showcasing the robustness of the method.

We further examined the estimation accuracy of causal effects for these five methods. Fig. 3I shows the results under *β* = 0.3 and a correlation of 0.7. CAUSE and Weighted-mode underestimated the causal effects, in line with their relatively lower power observed in earlier experiments. Among the three unbiased methods—XMR, MRAPSS, and Egger— Egger displayed the greatest estimation variance. While MRAPSS reduced estimation error substantially, it still lagged behind XMR. Additional results for different *β* and correlation settings can be found in Supplementary section 4.3, where the main performance patterns remain the same.

### XMR maintains well-calibrated *P* values in real-data negative control studies

Our goal is to achieve high power while ensuring type I error remains under control in real data analysis. As a first step before comparing the power of different methods, we evaluated their false positive rates. Beyond simulations, we conducted a more realistic negative control experiment using real-world data. Following Sanderson et al. [37] and Hu et al. [21, 38], we selected two traits determined at birth—natural hair color before greying and ease of skin tanning—as outcomes. For exposures, we considered 35 physiological or biochemical indicators, such as weight, arm fat percentage, and albumin. On the one hand, these indicators, measured later in life, cannot causally influence traits established at birth. On the other hand, the outcomes are prone to confounding factors such as population structure. They serve as ideal examples for negative control studies.

Using GWAS data from Pan-UKBB, we performed experiments on African and Central/South Asian populations, with Europeans serving as the auxiliary large-sample population. The sample sizes for the target populations were approximately 6,000 and 8,000, respectively, while the EUR cohort consisted of over 400,000 individuals, reflecting a substantial disparity in sample sizes between European and non-European populations. To assess the robustness of MR methods, we tested their performance under three different thresholds, as shown in Fig. 4.

**Figure 4.**
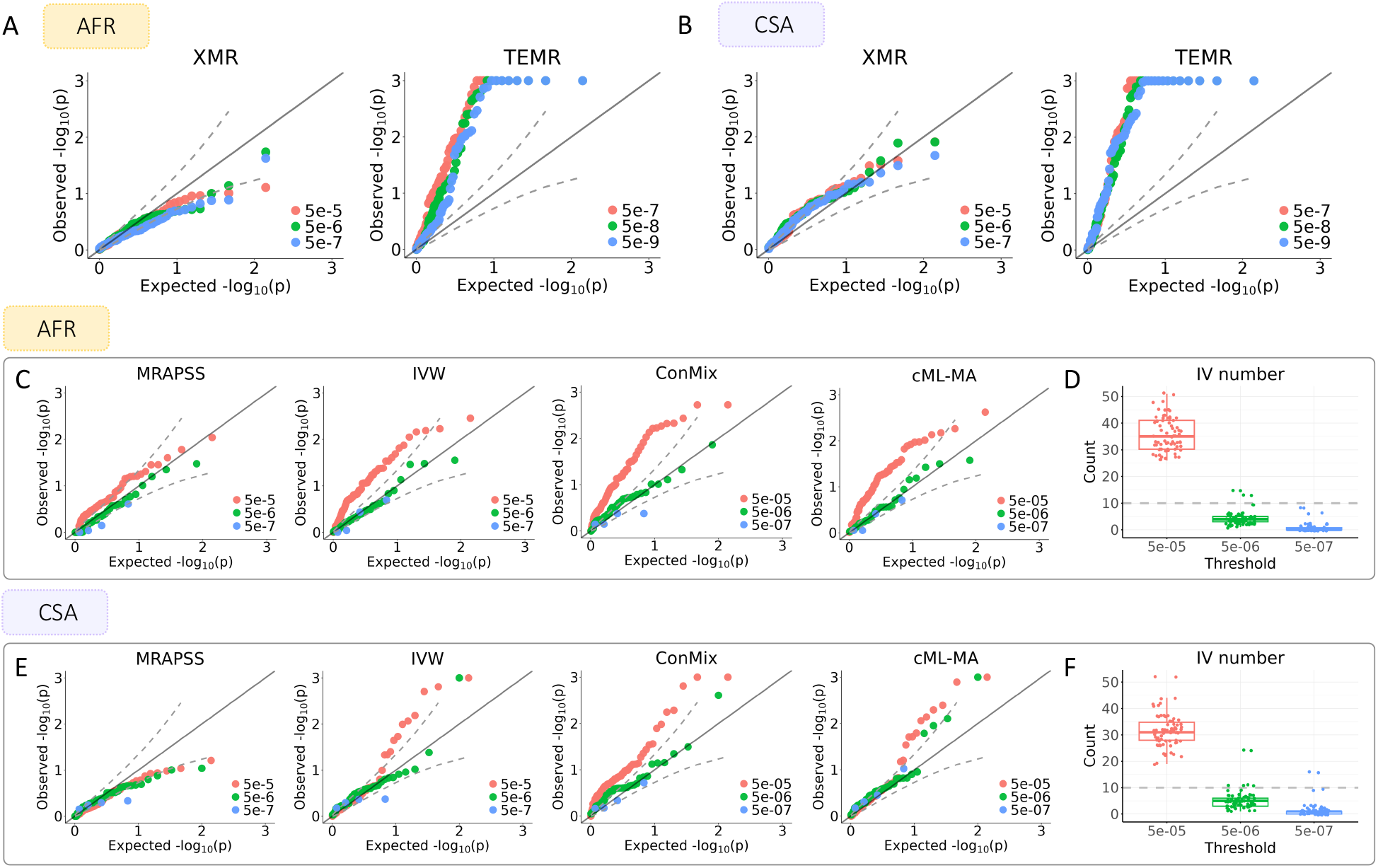
Real-data negative control studies. Evaluation of the false positive rates of MR methods using real-world negative control data. **(A, B)** QQ plots of *−*log_10_(*p*) values from two cross-population MR methods, XMR and TEMR, under different IV selection thresholds in AFR and CSA populations, respectively. **(C, E)** QQ plots of *−*log_10_(*p*) values from several single-population MR methods under different IV selection thresholds in AFR and CSA populations, respectively. **(D, F)** Box plots of the number of IVs retained after selection under different thresholds in AFR and CSA populations, respectively.

First, we compared XMR with the other cross-ancestry MR method, TEMR. In Figs. 4A, B, XMR generated well-calibrated *P* values for both AFR and CSA populations. Its performance remained stable across different IV selection thresholds, demonstrating its robustness to varying criteria. In contrast, TEMR yielded overly inflated *P* values in both populations, regardless of the threshold applied. This discrepancy likely stems from the inclusion of invalid IVs in TEMR’s augmented IV set. While the large EUR cohort enhances statistical power, TEMR’s inability to properly account for these invalid IVs introduces significant bias. Additionally, its simplified modeling framework fails to adequately correct for confounding factors in both target and auxiliary populations. These shortcomings contribute to its seemingly high statistical power, achieved at the expense of elevated false positive rates. To further investigate this, we examined ablated versions of XMR (**Ω** = 0, ***C*** = *I*, or no correction of selection bias). The results also exhibited certain degrees of inflation (see Supplementary section 4.5, Fig. S9), suggesting the importance of correcting for correlated pleiotropy, sample structure, and selection bias.

Next, we further examined the performance of 14 single-population MR methods. With sample sizes of fewer than 9,000 in each target population, the *P* values for exposure summary statistics were often not significant enough, resulting in very few SNPs passing the IV selection step. When the selection threshold was relaxed to 5 *×* 10^*−*5^, around 40 SNPs were retained, offering an acceptable number of IVs, as shown in Figs. 4D, F. However, under this relaxed threshold, inflated type I error rates were observed across most two-sample MR methods, with the notable exception of MRAPSS, Weighted-mode, Egger, CAUSE, and Weighted-median (Figs. 4C, E; Supplementary section 4.4, Figs. S7–S8). This issue likely arises from inadequate handling of pleiotropy and population structure in GWAS summary statistics [21]. Under more stringent thresholds of 5 *×* 10^*−*6^ and 5 *×* 10^*−*7^, nearly all methods produced properly calibrated *P* values. However, these estimations were typically based on no more than 5 IVs (Figs. 4D, F), far below credible standards. In some cases, the small number of IVs prevented certain methods from running altogether, as reflected by the reduced number of green and blue dots in the plots. This underscores a critical dilemma when applying single-population MR methods to small groups: strict thresholds lead to unreliable or failed estimations, while looser thresholds increase false positive rates.

Through these negative control experiments, XMR showed its capability to provide a solution for this situation by leveraging a large auxiliary population and effectively modeling confounding factors. The findings from the real-data negative control experiments largely aligned with the results observed in simulation null cases. Combining evidence from both analyses, we retained five methods that maintained reasonable *P* value calibration: XMR, MRAPSS, Weighted-mode, Egger and CAUSE. A complete list of causal effect estimations across all compared methods is given in Supplementary, Tables S5–S7 and S10–S12.

### XMR uncovers causal heterogeneity in East Asians and demonstrates superior replicability

To demonstrate XMR’s utility in characterizing causal associations within underrepresented populations, we first applied the method to the EAS population using BBJ data, with EUR data serving as an auxiliary. We analyzed 105 pairs linking 7 risk factors to 15 diseases. The Benjamini-Hochberg (BH) procedure was applied at an FDR level of 0.05. We mainly compared the statistical power of XMR with that of MRAPSS, Weighted-mode, Egger and CAUSE, as other methods exhibited higher type I errors than the nominal level in prior experiments.

XMR identified 30 significant causal effects after BH correction (Fig. 5A), substantially outperforming the alternative methods (Fig. 5B). Notably, XMR captured 15 of the 16 pairs identified by MRAPSS, Weighted-mode, Egger and CAUSE, and detected 15 unique signals (see Supplementary section 4.6.2, Fig. S11). These unique discoveries are supported by existing literature. For instance, XMR identified 9 significant effects of BMI; 8 are supported by established evidence, including associations with T2D [39], three cardiovascular diseases (myocardial infarction (MI) [40], angina pectoris (AP) [40], and unstable angina pectoris (UAP) [41]), asthma [42], cholelithiasis (GS) [43], cataract (Cat) [44], and breast cancer (BC) [45, 46]. The remaining pair indicated a negative effect of BMI on prostate cancer (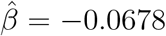, *P* = 7.51 *×* 10^*−*3^), a protective association observed in some epidemiological studies [47, 48]. In contrast, among these BMI-related pairs, only Weighted-mode and CAUSE detected the link to T2D. Furthermore, XMR was the sole method to identify the causal effects of DBP on four traits, all of which are biologically plausible and supported by prior research, including peripheral artery disease (PAD) [49], MI [50], AP [50] and epilepsy (Epi) [51]. The inability of MRAPSS, Weighted-mode, Egger, and CAUSE to detect these pairs highlights the superior statistical power of XMR in sample-limited populations. A comprehensive comparison of discoveries across all methods is presented in Supplementary section 4.6, Figs. S10, S12.

**Figure 5.**
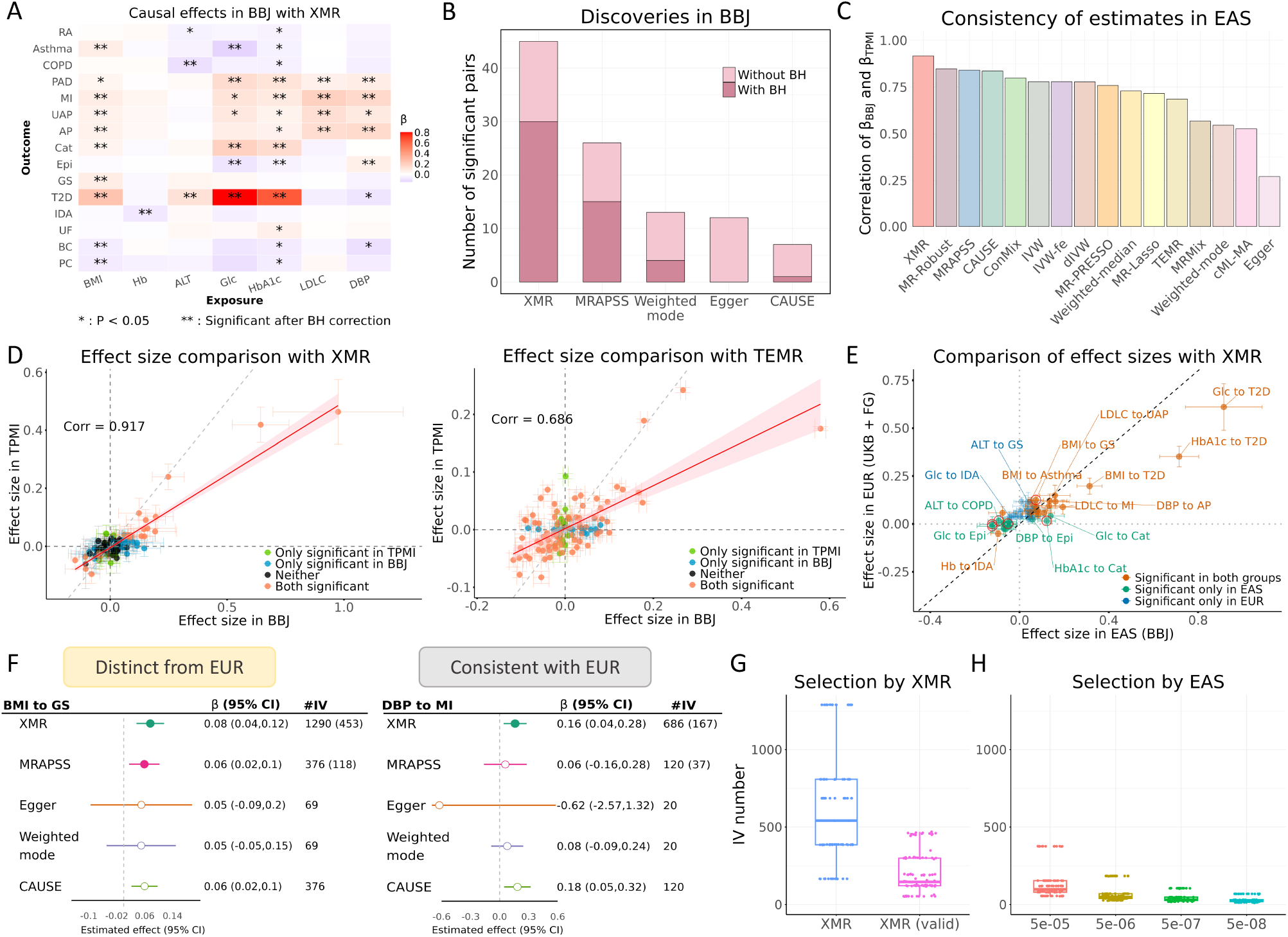
Causal relationships in the EAS population. Exploration of causal effects among various traits in East Asians. **(A)** Causal effect estimates *β* from XMR in BBJ, with red indicating positive effects and purple indicating negative effects. Significance is marked by * (*P <* 0.05) and ** (significant after BH correction). **(B)** Number of significant pairs identified by five methods, with light and dark pink representing results without and with BH correction, respectively. **(C)** Correlations of effect sizes between BBJ and TPMI across different MR methods. **(D)** Comparison of effect sizes in BBJ and TPMI estimated by XMR (left) and TEMR (right), with 95% CIs. Point colors denote significance in the two datasets; the red line shows linear regression results, with the shaded area representing 95% CIs; the grey dashed line represents *y* = *x*. **(E)** Comparison of effect sizes in EAS (by XMR) and EUR (by MRAPSS), with 95% CIs. Point colors indicate significance in two populations, the black dashed line represents *y* = *x*, and red circles highlight heterogeneous pairs validated by TPMI. **(F)** Case studies of causal effect estimates (*β*) in BBJ with 95% CIs and IV counts. Numbers of valid IVs for XMR and MRAPSS are shown in parentheses. Solid points indicate *P <* 0.05. **(G–H)** Box plots of IV counts across all analyzed trait pairs in BBJ. (G): total IVs and valid IVs from XMR; (H): IVs selected using EAS alone under different thresholds.

To rigorously validate these findings, we performed a replication analysis using an independent EAS cohort—TPMI. A robust MR method should yield consistent estimates across two cohorts of the same ancestry. XMR achieved the highest consistency among all 16 tested methods, with a correlation of 0.917 between BBJ and TPMI estimates (Fig. 5C). Among methods with controlled false-positive rates, MRAPSS and CAUSE also showed high correlations. However, Weighted-mode and Egger performed poorly (correlations of 0.546 and 0.271, respectively), likely due to their low precision and broad confidence intervals. Notably, we observed concerning inconsistencies in other high-power methods. While it is acknowledged that effect sizes across the two cohorts (BBJ and TPMI) may differ, it is highly unlikely that the direction of the effect would be reversed. XMR maintained consistent effect directions for all pairs significant in both BBJ and TPMI (*P <* 0.05), yet TEMR—despite making many discoveries—produced inflated estimates and, alarmingly, 23 pairs with conflicting directions between the two EAS cohorts (Fig. 5D). Inconsistencies were also observed in IVW-fe and ConMix (see Supplementary section 4.6.3, Fig. S14). Similarly, cML-MA yielded a correlation of only 0.527, indicating that its findings are poorly replicable. This comparison demonstrates that XMR offers a better trade-off: sufficient power to detect signals missed by conservative methods, without the instability and high false discovery rates associated with other aggressive methods.

We next investigated the cross-population architecture of these causal effects between EAS and EUR. As indicated in Fig. 5D and Supplementary section 4.6.3, Figs. S13–S14, estimates in BBJ tended to be systematically larger than in TPMI, potentially due to environmental differences or measurement heterogeneity. To obtain more robust EAS estimates for cross-ancestry comparison, we conducted a meta-analysis of the two EAS cohorts following Xiao et al. [52] (details in Supplementary section 2.3.4) and used the meta-analyzed estimates for further analysis. As shown in Fig. 5E, estimates in EAS and EUR for the 21 pairs significant in both populations generally aligned along the identity line, suggesting shared causal mechanisms. For example, the causal effects of DBP on cardiovascular diseases (e.g. MI, AP) were detected in both groups and exhibited similar effect sizes. However, XMR also revealed significant heterogeneity (highlighted in red boxes, Supplementary section 4.6.4, Fig. S15). We identified 17 trait pairs that were significant in BBJ and significantly different from EUR; notably, 6 of these pairs were successfully replicated in TPMI, representing high-confidence population-specific effects (see Supplementary section 3.1, Table S1).

Among these, we highlight two distinct patterns of heterogeneity. First, XMR identified population-specific risk factors. For example, glycemic traits, Glc (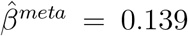, *P* = 1.05 *×* 10^*−*5^) and HbA1c (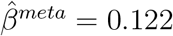, *P* = 1.14 *×* 10^*−*9^), were identified as significant risk factors for cataracts in BBJ but did not pass BH correction in EUR. The effect of HbA1c on cataracts was further validated by TPMI (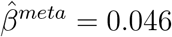, *P* = 1.13 *×* 10^*−*5^). While diabetes is a known risk factor [53], a recent longitudinal study suggests that elevated glucose accelerates cataract progression even in non-diabetic individuals [54]. The underlying mechanisms include persistent hyperglycemia causing changes in intraocular pressure (IOP) and glycoprotein composition, leading to lens clouding and reduced visual acuity [55]. Our finding, along with previous studies, underscores the importance of glycemic control for ocular health specifically in East Asians, given the substantial public health and economic burden posed by cataracts and cataract surgery [54]. Second, we observed heterogeneity in effect magnitude. The effect of BMI on cholelithiasis (GS) was stronger in EUR (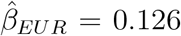, *P*_*EUR*_ = 7.20 *×* 10^*−*31^) than in EAS 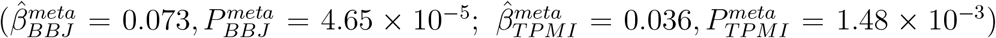. This may partially explain the higher prevalence of cholelithiasis in Europeans and different patterns of the disease, where 80–90% of gallstones in Western populations are cholesterol stones with obesity as a strong risk factor, while pigment stones dominate gallstones found in Asian populations, which are mainly associated with parasitic infections and bacterial infections [56, 57, 58]. Additionally, several pairs showing unique heterogeneity in BBJ— though not strictly replicated in TPMI—are consistent with established literature. For example, the causal effect of LDLC on MI, a type of atherosclerotic cardiovascular disease (ASCVD), was markedly stronger in EAS 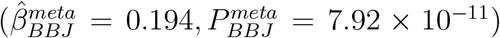 than in EUR (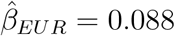, *P*_*EUR*_ = 2.58 *×* 10^*−*6^). This finding aligns with evidence that suggests distinctive patterns of lipoprotein cholesterol distributions and incidence of ASCVD in East Asians, who tend to have lower LDLC levels than Europeans and distinct ASCVD risk factors [59, 60]. A meta-analysis by Li et al. also demonstrated that East Asians may require only small reductions in LDLC to achieve regression of coronary atherosclerotic plaque compared to Western populations [61], supporting population-specific treatment strategies. Collectively, these results demonstrate XMR’s capability to characterize both shared and distinct causal architectures in underrepresented populations relative to Europeans. Notably, even when leveraging the augmented power of meta-analysis, alternative methods—including MRAPSS, Egger, Weighted-mode, and CAUSE—identified significantly fewer signals than XMR (Supplementary section 4.6.4, Fig. S16, and section 4.6.5, Fig. S17).

Finally, we illustrate the power of XMR through two specific examples in BBJ: BMI to GS (heterogeneous between EAS and EUR) and DBP to MI (shared between the two populations). As shown in Fig. 5F, Egger suffered from wide confidence intervals and low statistical power, while CAUSE yielded narrow intervals but potentially deflated *P* values. Only XMR consistently detected significant effects with high precision in both cases. This precision is driven by the effective expansion of the instrument pool; as shown in Figs. 5G, H, even the valid IVs utilized by XMR significantly outnumber those available when using EAS summary statistics alone, confirming the benefit of incorporating auxiliary EUR data. A full list of causal effect estimates from all 16 methods across BBJ, TPMI, and EUR is provided in Supplementary Tables S16–S18. The meta-analyzed results for the two EAS cohorts are detailed in Supplementary Table S19.

### XMR displays robust power to reproduce causal findings in under-represented populations

We extended our analysis to Central/South Asian and African populations to evaluate XMR’s performance under conditions of even more limited sample sizes. Specifically, we investigated bidirectional causal effects among five diseases (T2D, HTN, CIHD, DLM, and asthma) and between these diseases and 11 quantitative traits in CSA (8 in AFR). Despite the restricted sample sizes (approximately 8,000 for CSA and 6,000 for AFR), XMR successfully identified 15 significant causal relationships in CSA after BH correction (Fig. 6A). In contrast, among compared methods, only Egger detected a single signal (T2D on HbA1c, 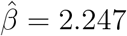, *P* = 1.68 *×* 10^*−*5^), while MRAPSS, Weighted-mode and CAUSE failed to report any significant findings. The disparity was even more pronounced in AFR: all four traditional MR methods failed to identify any significant causal relationship, suggesting their limited statistical power in underrepresented cohorts (Fig. 6B). However, XMR remained robust, identifying 3 significant pairs passing BH correction. When the significance threshold was relaxed to nominal *P <* 0.05, XMR uncovered 31 and 14 potential causal pairs in CSA and AFR, respectively. While few unique relationships were identified—likely due to sample size constraints—the direction and magnitude of effect estimates showed high concordance with those observed in EUR (Figs. 6C, D). This consistency, particularly for pairs significant in both populations, demonstrates XMR’s superior capability to replicate established European findings in small-sample diverse cohorts.

**Figure 6.**
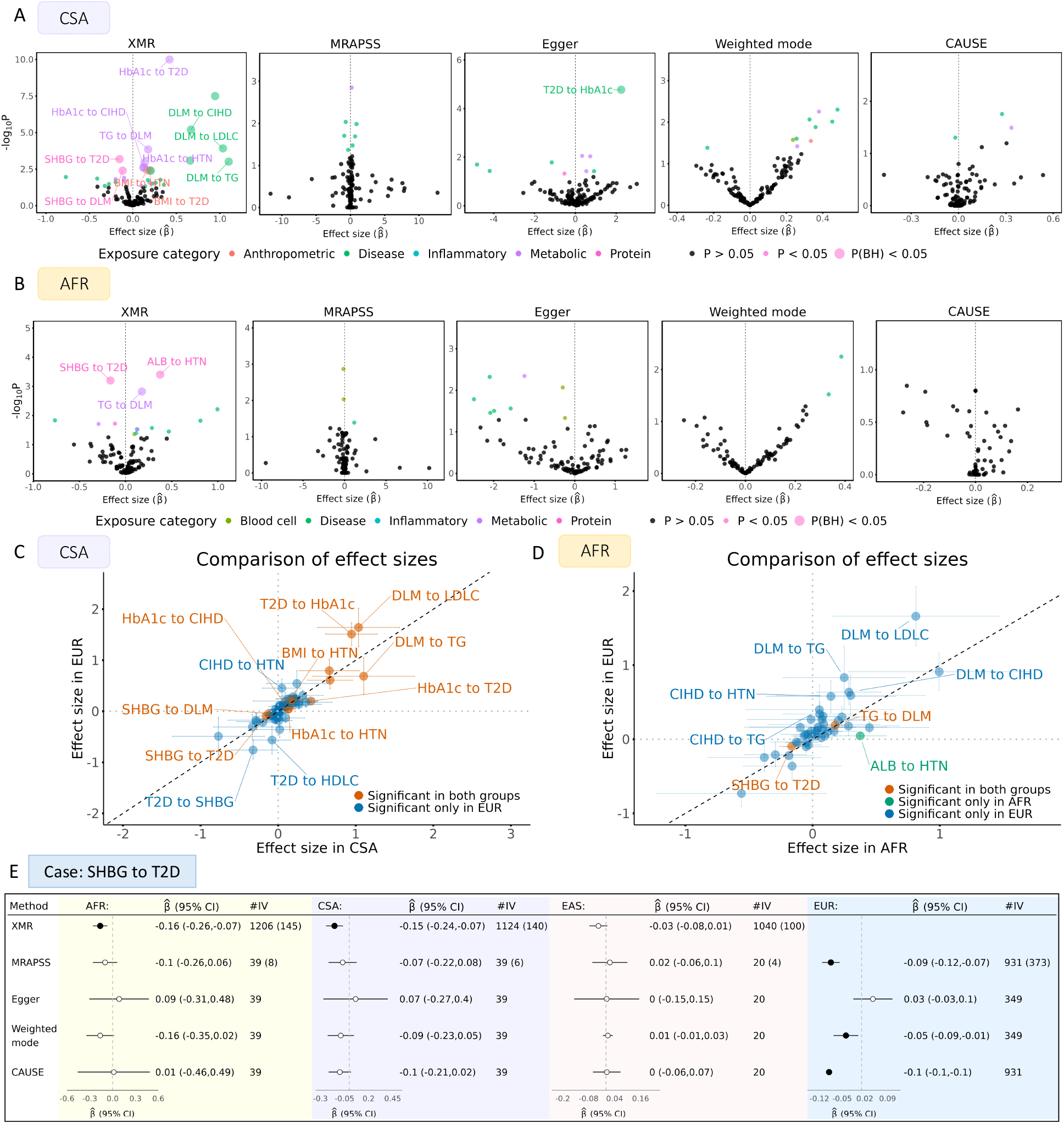
Causal relationships in CSA and AFR populations. Exploration of causal effects among various traits in Central/South Asians and Africans. **(A–B)** Scatter plots of causal effect estimates from five methods showing effect sizes 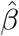 against *−*log_10_(*p*) values of each trait pair in CSA and AFR, respectively. Points with *P <* 0.05 are colored by exposure category and shaped based on significance after BH correction. **(C–D)** Comparison of effect sizes in CSA/AFR (by XMR) and EUR (by MRAPSS), with 95% CIs. Point colors indicate the significance after BH correction in two populations. Black dashed line: *y* = *x*. **(E)** Assessment of SHBG’s causal effect on T2D in four populations using five methods (effect size estimates and 95% CIs). Black solid dots: causal effects with *P <* 0.05. The number of valid IVs inferred by XMR or MRAPSS is indicated in parentheses.

Specifically, disorder of lipoid metabolism encompasses abnormalities in lipid synthesis, breakdown, absorption, and transport, affecting molecules such as cholesterol and triglycerides [62]. XMR effectively captured the defining characteristics of DLM in CSA and AFR. We observed bidirectional causal effects between DLM and lipid traits, including positive effects of DLM on TG (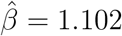, *P* = 9.73 *×* 10^*−*4^) and LDLC (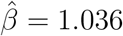, *P* = 1.20 *×* 10^*−*4^), as well as the reverse effect of TG on DLM in both populations (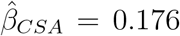, *P*_*CSA*_ = 1.43 *×* 10^*−*4^; 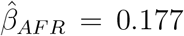, *P*_*AFR*_ = 1.50 *×* 10^*−*3^). Furthermore, XMR identified HbA1c (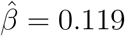, *P* = 2.51 *×* 10^*−*3^) and SHBG (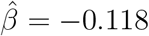, *P* = 4.01 *×* 10^*−*3^) as upstream causal factors for DLM in CSA. The protective role of SHBG was also nominally significant in AFR (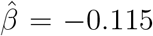, *P* = 0.019). Biologically, the link between HbA1c and dyslipidemia is well-documented across diverse ancestries [63, 64, 65], likely driven by hyperglycemia-induced disruptions in fatty acid oxidation and signaling pathways [66, 67]. Similarly, SHBG is linked to various lipid components [68, 69] and known to regulate hepatic lipogenesis; experimental evidence suggests that SHBG overexpression downregulates key lipogenic enzymes, thereby improving lipid profiles [70, 71], consistent with our findings.

Next, for CIHD, XMR identified HbA1c as a risk factor in CSA (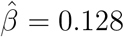, *P* = 2.37*×*10^*−*3^), aligning with established literature [72, 73]. Additionally, a strong causal effect of DLM on CIHD was detected (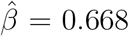, *P* = 6.32 *×* 10^*−*6^), reflecting the canonical pathway where dyslipidemia drives atherosclerosis through plaque deposition, inflammation, and endothelial dysfunction [74]. Notably, this DLM-CIHD signal was also replicated in AFR at a nominal significance level (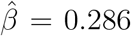, *P* = 0.027), suggesting a shared etiology across populations despite the reduced statistical power.

Although no risk factors passed BH correction for asthma, nominal significance (*P <* 0.05) revealed potential relationships with C-reactive protein (CRP). Specifically, we observed a positive effect of CRP on asthma in AFR (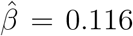, *P* = 0.040) and a reverse effect of asthma on CRP in CSA (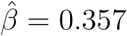, *P* = 0.038), hinting at the inflammatory nature of the disease [75].

For hypertension, XMR identified BMI (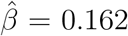, *P* = 3.66 *×* 10^*−*3^) and HbA1c (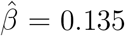, *P* = 1.15*×*10^*−*3^) as risk factors in CSA. Notably, the effect size of HbA1c on hypertension in CSA was markedly stronger than in EUR (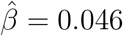, *P* = 6.78 *×* 10^*−*4^). While the impact of BMI is extensively documented and widely attributed to pathways such as sympathetic activation and renal dysfunction [76, 77], the link between HbA1c and hypertension has been less studied and is gaining recognition. Hyperglycemia may elevate blood pressure through insulin resistance-induced endothelial impairment, sodium retention, and oxidative stress [78, 79]. Clinical observations suggest a unique pattern in which South Asians exhibit higher HbA1c levels, higher rates of diabetes, and higher prevalence of hypertension compared to Europeans [80, 81]. Our finding of a stronger effect in CSA aligns with studies revealing a greater susceptibility of South Asians to hyperglycemia-related cardiovascular complications compared to Europeans [82, 83]. Uniquely, in the African population, XMR identified a positive causal effect of serum albumin levels on hypertension (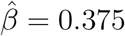, *P* = 3.95 *×* 10^*−*4^). While the ALB-hypertension relationship is complex, a positive association has been reported in some past studies [84, 85, 86]. This finding is particularly intriguing given that the albumin GWAS data used here identified African-specific genetic variants that were extremely rare in other ancestries [36]. This reinforces the unique causal signal detected by XMR and underscores the importance of leveraging population-specific genetic architecture to uncover distinct causal signals.

For T2D, XMR confirmed well-known positive causal effects of BMI (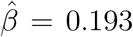, *P* = 3.90 *×* 10^*−*3^) and HbA1c (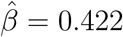, *P* = 1.37 *×* 10^*−*20^) in CSA, alongside a protective effect of SHBG in both CSA (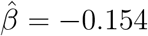, *P* = 6.51 *×* 10^*−*4^) and AFR (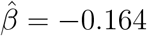, *P* = 6.30 *×* 10^*−*4^). To rigorously benchmark XMR’s performance against other methods, we conducted a four-population case study on the SHBG-T2D relationship (Fig. 6E). The SHBG data for EAS were sourced from Pan-UKBB, with a sample size of 2,327. In EUR (*N* = 363, 650), nearly all methods correctly identified the protective effect of SHBG, supported by extensive genetic and functional evidence linking SHBG to glucose homeostasis and insulin sensitivity [87, 88, 89]. Two pivotal genetic studies, primarily conducted in European populations, have significantly strengthened the evidence supporting SHBG’s involvement in the disease’s etiology [87, 88]. These studies identified germline variants in the SHBG gene that were directly associated with both plasma SHBG levels and the risk of T2D. SHBG appears to influence T2D development by modulating the biological activity of sex hormones in peripheral tissues. Moreover, recent findings suggest that the association between low SHBG levels and increased T2D risk may be partially mediated by increased visceral adipose tissue and liver fat accumulation [89].

However, as sample sizes decreased in non-European cohorts, the performance of traditional methods deteriorated. In AFR (*N* ≈ 5, 700) and CSA (*N* ≈ 7, 600), XMR was the only method to detect a significant causal effect. Even in East Asians (*N* = 2, 327), where sample sizes were insufficient for any method to reach significance, XMR still estimated a negative effect direction consistent with the other populations. As clinical studies on SHBG in these specific populations are limited and still emerging [90, 91, 92], our results provide strong statistical evidence reinforcing the universality of this biological pathway. This case study exemplifies the critical advantage of XMR: while other methods require massive sample sizes (available primarily in EUR) to function, XMR maintains sufficient power to uncover accurate causal relationships in underrepresented, small-sample populations, thereby bridging a critical gap in global health equity and precision medicine.

Additional results for MRAPSS, Egger, Weighted-mode, and CAUSE are provided in Supplementary sections 4.7.1 and 4.8.1, Figs. S18–S19. A comprehensive list of causal effect estimates from all 16 methods is available in Supplementary Tables S21–S22 and S24–S25. Trait pairs exhibiting effect heterogeneity compared to EUR by XMR are summarized in Supplementary sections 3.2 and 3.3, Tables S2–S3.

## Discussion

In this paper, we proposed a cross-population MR method, XMR, designed to enhance summary-level causal inference for underrepresented populations. This is achieved mainly by utilizing summary statistics for the same exposure from a large-sample auxiliary population. By assuming that the genetic basis of an exposure trait may be shared across populations, XMR expands the pool of IVs and incorporates the auxiliary exposure into a probabilistic model to refine causal effect estimates. To ensure robust inference, XMR is designed to mitigate biases from invalid IVs that may be introduced, as well as those arising from confounding factors and selection bias. Through extensive simulations and real-data analyses, XMR demonstrated superior performance in balancing high statistical power with low type I error rates, outperforming traditional two-sample MR methods and recently developed trans-ethnic MR methods.

In light of the scarcity of genetic data and research for non-European populations, we hope XMR can provide valuable insights into causal relationships in these groups, thereby helping to address medical disparities and advancing global health equity. Therefore, we applied XMR to a series of trait pairs for East Asians, Central/South Asians and Africans, using Europeans as the auxiliary population. XMR consistently identified significant causal relationships undetected by previous MR methods. Many of these findings agreed with results from Europeans, which aligns with expectations given the substantially shared genetic architecture and causal mechanisms across populations. This helps bridge the gap in scientific evidence for the transportability of many findings from European studies, which is increasingly supported by emerging studies in target populations. Notably, XMR also uncovered causal relationships that appeared unique to underrepresented populations or exhibited distinct effect sizes. These findings are particularly noteworthy and may reveal population-specific genetic structures or causal pathways, potentially raising important considerations for precision treatment. We attempted to explore potential biological mechanisms underlying our findings and contextualized them through comparisons with established literature, despite the general lack of studies focusing on the target populations. For instance, the smaller effect of BMI on cholelithiasis in East Asians aligns with the distinct etiology of cholelithiasis in this population, where infection-driven pigment stones are relatively more common compared to Western populations. The stronger effect of HbA1c on hypertension in Central/South Asians also corresponds to the higher susceptibility to hypertension and more severe outcomes observed in this group. Such concordance emphasizes the importance of XMR’s findings and motivates further in-depth studies in underexplored populations.

Despite the advantages of XMR, we are aware of several limitations in our study. First, MR findings should be interpreted cautiously and ideally paired with additional experimental validation, such as randomized controlled trials (RCTs) or mechanistic studies to confirm causal relationships. This is particularly critical for cross-population differences, as observed heterogeneity may stem from differences in allele frequency, LD patterns, or genetic effects, rather than true causal differences [93]. Thus, our findings should serve as a starting point for deeper exploration rather than conclusive evidence. Second, although XMR is designed for target populations with limited sample sizes, an adequate sample size is still required for reliable LDSC estimation of confounding correction parameters. When the target sample is sufficient for stable LDSC estimation but far smaller than large auxiliary populations—as with the East Asian cohort in this study—XMR demonstrates clear advantages over methods relying solely on the target population data. Third, our method can exclude invalid IVs automatically, which has been shown to be effective in simulations and works well empirically. Nevertheless, further investigation is needed to determine whether the selected IVs fully satisfy all MR assumptions. Additionally, our IV selection is based only on the auxiliary population to facilitate selection bias correction. However, incorporating unique IVs from the target population could further enhance statistical power and facilitate the detection of population-specific causal signals. Furthermore, while approaches incorporating explicit fine-mapping strategies, such as XMAP [94], offer an alternative for addressing LD structures, they often entail substantial computational burdens and rely on complex modeling assumptions. Therefore, we adopted a more practical strategy that effectively accounts for LD patterns without incurring these additional complexities.

In conclusion, XMR offers a robust and innovative framework for improving causal inference in underrepresented populations. By addressing the challenges posed by limited data and leveraging cross-population genetic information, XMR bridges critical gaps in genetic research, paving the way for more inclusive and equitable precision medicine.

## Methods

### The XMR framework

As introduced in the Method overview section, XMR is designed to enhance causal inference between an exposure *X*_2_ and an outcome *Y*_2_ in a small-sample target population by leveraging GWAS data from the same exposure *X*_1_ in a large-sample population. The method integrates summary statistics from three GWAS datasets: 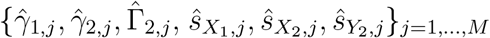, where 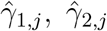 and 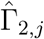 denote the estimated effect sizes of the *j*-th SNP on *X*_1_, *X*_2_, and *Y*_2_, respectively, and 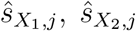 and 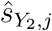 are their corresponding standard errors. Standard MR methods rely on three core assumptions for valid causal inference: (I) Relevance: The instrumental variables (IVs) are strongly associated with the exposure; (II) Independence: The IVs are independent of any confounding factors affecting the outcome; and (III) Exclusion Restriction: The IVs influence the outcome only through the exposure, not via alternative pathways. The XMR framework operates in three key stages to ensure these assumptions are satisfied: (1) IV selection, (2) confounding correction, and (3) causal effect estimation. During the IV selection stage, SNPs strongly associated with *X*_1_ are selected based on a significance threshold 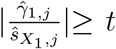, resulting in a set of *M*_t_ candidate SNPs that satisfy Assumption (I). These SNPs are subsequently analyzed using a probabilistic model that disentangles causal effects from confounding influences. By incorporating cross-population correlation between SNP effects in *X*_1_ and *X*_2_, XMR improves the robustness and statistical power of the analysis compared to traditional MR approaches that rely solely on single-population data.

The relationship between SNP effect sizes across the three datasets is expressed using the following probabilistic model, which consists of a causal inference module and a confounding correction module:

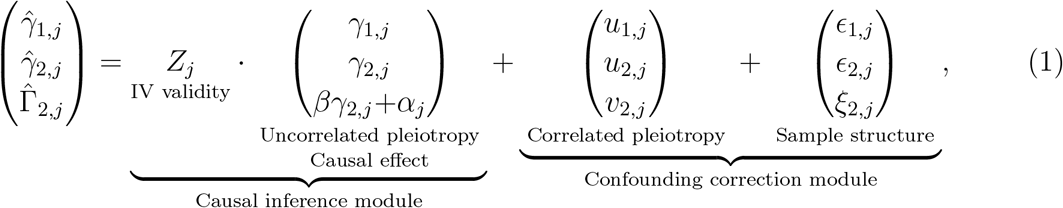

where *β* represents the causal effect of interest. To address Assumption (II), the model explicitly accounts for confounding effects, including correlated pleiotropy and population structure, via the terms (*u*_1,*j*_, *u*_2,*j*_, *v*_2,*j*_) and (*ϵ*_1,*j*_, *ϵ*_2,*j*_, *ξ*_2,*j*_). Furthermore, we introduce a binary latent variable *Z*_*j*_, which indicates whether SNP *j* is a valid IV (*Z*_*j*_ = 1) or not (*Z*_*j*_ = 0). This mechanism automatically identifies and excludes IVs that violate Assumptions (II) or (III) (e.g., through horizontal pleiotropy represented by *α*_*j*_), thereby ensuring robust causal estimation.

### Confounding correction module

The confounding correction module addresses two primary sources of bias in MR analyses: correlated pleiotropy and sample structure effects, which can obscure causal relationships. By explicitly modeling these confounders, XMR ensures that the observed SNP-trait associations more accurately reflect causal relationships rather than spurious correlations.

Correlated pleiotropy occurs when SNPs influence multiple traits through shared biological mechanisms, leading to non-causal associations between *X*_1_, *X*_2_ and *Y*_2_. This source of bias is accounted for by assuming that the pleiotropic effects (*u*_1,*j*_, *u*_2,*j*_, *v*_2,*j*_) follow a multivariate normal distribution:

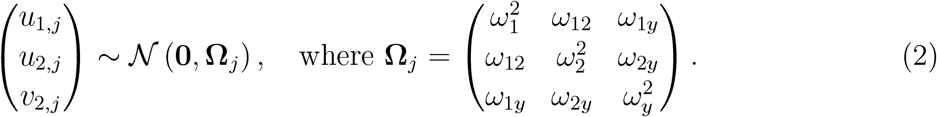

Here 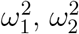 and 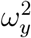 are the variances of polygenic effects on *X*_1_, *X*_2_ and *Y*_2_, respectively, while *ω*_12_, *ω*_1*y*_ and *ω*_2*y*_ quantify the genetic covariances between these traits. Given that linkage disequilibrium influences genetic correlations, the covariance matrix **Ω**_*j*_ is adjusted to reflect local genetic architecture:

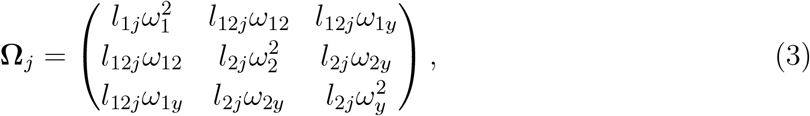

where *l*_1*j*_, *l*_2*j*_ and *l*_12*j*_ are the LD scores of SNP *j*, representing its correlation structure within the large-sample population, the small-sample population, and across the two populations, respectively.

Sample structure is independent of LD and can therefore be distinguished from correlated pleiotropy using LD Score Regression (LDSC) [95]. We model sample structure as:

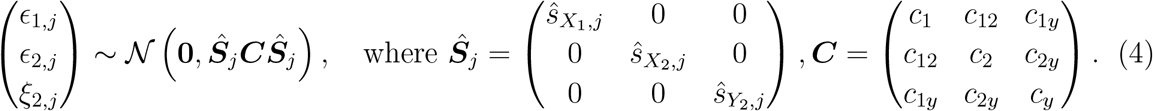

Here ***Ŝ***_*j*_ is a diagonal matrix of the standard errors 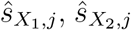 and 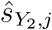, which accounts for the varying uncertainty across datasets. The matrix ***C*** describes residual correlations due to shared sample structure, with the diagonal elements *c*_1_, *c*_2_ and *c*_*y*_ representing population-specific variance, and the off-diagonal elements *c*_12_, *c*_1*y*_ and *c*_2*y*_ modeling cross-dataset correlations. When there is substantial sample structure, the diagonal parameters *c*_1_, *c*_2_ and *c*_*y*_ are expected to deviate significantly from 1, which is more common in large-sample populations. The off-diagonal term *c*_2*y*_ can be attributed to factors like sample overlap between *X*_2_ and *Y*_2_, while nonzero values for *c*_12_ and *c*_1*y*_ often arise from shared environmental, demographic, or socioeconomic factors that contribute to correlations across the two populations. The diagonal matrix ***Ŝ***_*j*_ adjusts for differences in sample size, with smaller-sample populations typically exhibiting greater uncertainty. This adjustment ensures that the effects of ***C*** are appropriately balanced.

Both **Ω**_*j*_ and ***C*** are pre-estimated from genome-wide summary statistics using LDSC prior to the IV selection process. The use of genome-wide SNPs for background parameter estimation, distinct from the selected IVs used for causal inference, contributes to the identifiability of the model. In summary, the combined variance component **Ω**_*j*_ + ***Ŝ***_*j*_ ***CŜ***_*j*_ provides a comprehensive background adjustment for confounding effects, ensuring reliable causal estimation. See more details in Supplementary section 1.2.

### Causal inference module

With confounding effects accounted for by the pre-estimated parameters, XMR estimates the causal effect *β* using the subset of *M*_*t*_ SNPs selected as IVs. We assume that SNPs may have direct effects on the outcome *Y*_2_ (*α*_*j*_) in addition to their indirect effects via the exposure *X*_2_. Since the correlated pleiotropy has already been accounted for in the confounding correction step, the InSIDE (INstrument Strength Independent of Direct Effect) assumption [18] can be imposed. Specifically, the direct effect *α*_*j*_ is assumed to be independent of the true SNP effects on *X*_1_ and *X*_2_, i.e. *α*_*j*_ ⊥ (*γ*_1,*j*_, *γ*_2,*j*_). However, *γ*_1,*j*_ and *γ*_2,*j*_ are assumed to be correlated due to the shared genetic structure. To capture these relationships, the effects (*γ*_1,*j*_, *γ*_2,*j*_, *α*_*j*_) are modeled as:

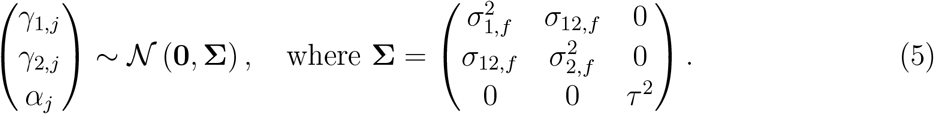

Here *σ*_12,*f*_ represents the covariance between *γ*_1,*j*_ and *γ*_2,*j*_, facilitating the integration of auxiliary information from the large-sample GWAS to improve statistical power.

To further refine causal inference, XMR introduces a binary indicator variable *Z*_*j*_, determining whether SNP *j* is a valid IV (*Z*_*j*_ = 1) or an invalid one (*Z*_*j*_ = 0). If *Z*_*j*_ = 0, the observed effects 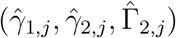 are assumed to be fully explained by the variance component derived from confounding factors, i.e. **Ω**_*j*_ + ***Ŝ***_*j*_ ***CŜ***_*j*_ . If *Z*_*j*_ = 1, SNP *j* is treated as a valid IV, and the observed SNP effects also include contributions from the true genetic effects (*γ*_1,*j*_, *γ*_2,*j*_, *βγ*_2,*j*_ + *α*_*j*_), according to Eq. (1). Under this scenario, the total variance component is given by: ***A***(*β*)**Σ*A***(*β*)^*T*^ + **Ω**_*j*_ + ***Ŝ***_*j*_ ***CŜ***_*j*_, where ***A***(*β*) encodes the causal relationship:

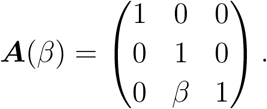

This design is particularly crucial for cross-population MR, as IV selection based on a large-sample population will inevitably include SNPs that are invalid in the target population. By incorporating *Z*_*j*_, XMR can tolerate the presence of invalid IVs during selection while automatically identifying and excluding them, thereby minimizing bias. This approach ensures that the enriched pool of filtered IVs can be fully utilized to enhance statistical power.

### Accounting for selection bias

Given the prior probability of SNP *j* being a valid IV, denoted as *π*_0_ = *p*(*Z*_*j*_ = 1), the full likelihood model integrates the causal inference and confounding correction modules. The joint probability of the observed SNP effects is expressed as:

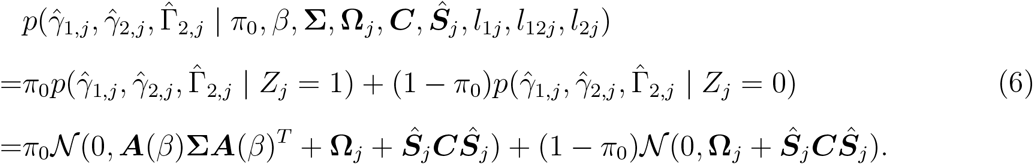

However, due to the pre-selection of IVs based on their association with 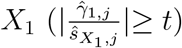, selection bias (commonly referred to as “winner’s curse” [96]) may distort the results. To address this issue, the likelihood is adjusted to account for the selection criteria:

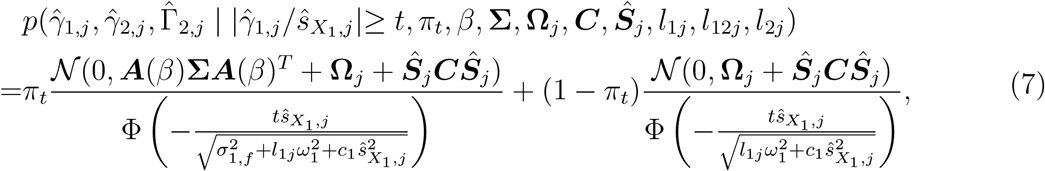

where 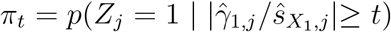. This adjustment ensures unbiased estimation of the causal effect.

In practice, LD clumping retains SNPs with smaller *P* values and thereby introduces additional selection bias. This can be mitigated by adjusting the threshold *t* to *t* + Δ*t*. This increment Δ*t* accounts for the relationship between the conditioning large-sample population *X*_1_ and the target population *X*_2_ used for LD clumping. Specifically, we demonstrate in Supplementary section 1.4 that Δ*t* depends on the sample size ratio and genetic correlation between these two populations.

### Parameter estimation and statistical inference

As mentioned in the Confounding correction module, the parameters 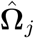 and ***Ĉ*** are pre-estimated using LDSC on genome-wide summary statistics. Fixing these components, the log-likelihood function for the observed data is given by:

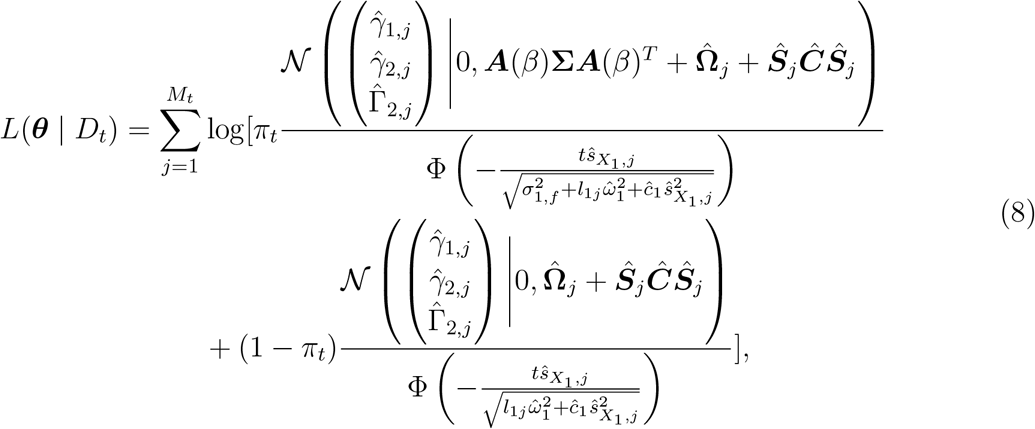

where ***θ*** = *{π*_*t*_, *β*, **Σ***}* denotes the parameters to be estimated and 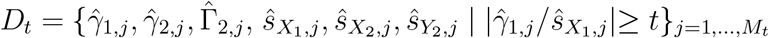 are the observed data for the *M*_*t*_ selected SNPs. To derive the maximum likelihood estimate of ***θ***, an Expectation-Maximization (EM) algorithm is employed. Details of the algorithm are provided in the Supplementary section 1.3.

To test the existence of a causal effect, we perform a likelihood ratio test:

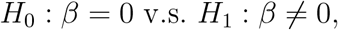

where the test statistic is computed as:

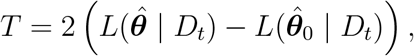

with 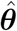 and 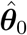 being the estimated parameters under hypotheses *H*_1_ and *H*_0_ respectively. The test statistic *T* follows a *χ*^2^ distribution with 1 degree of freedom, allowing for the computation of a *P* value. In addition to the causal effect estimation, the model also provides posterior probabilities *p*(*Z*_*j*_ = 1 | *D*_*t*_), the probability of each SNP being a valid IV. These probabilities are visualized in Fig. 1D, where SNPs are color-coded based on their posterior IV validity.

### Distinctions between XMR and MRAPSS

While XMR extends the probabilistic framework of MRAPSS to a cross-population setting, it introduces several distinct methodological advancements tailored to this context:

First, it addresses IV scarcity via auxiliary data. The most fundamental difference lies in XMR’s incorporation of an auxiliary population. MRAPSS relies solely on the target population for IV selection. In small-sample scenarios, this often leads to a severe shortage of valid IVs, resulting in insufficient statistical power (as shown in Figs. 3, 5, and 6) or even complete inapplicability (Fig. 4). By leveraging the auxiliary population, XMR ensures a sufficient number of IVs to enable robust inference.

Second, it enhances power through cross-population correlation. XMR explicitly models the genetic correlation, *σ*_12,*f*_, between SNP effects in the auxiliary exposure (*X*_1_) and the target exposure (*X*_2_). This parameter is essential for borrowing information from the larger auxiliary dataset to augment estimation in the smaller target dataset. As demonstrated in Supplementary section 4.10 (Figs. S28–S31), ignoring this correlation (i.e., enforcing *σ*_12,*f*_ = 0) significantly diminishes the statistical power, negating the benefit of the auxiliary data.

Third, it addresses selection bias arising from mismatched populations. A unique methodological challenge in cross-population MR is that LD clumping is applied to the target population, while IV selection is based on a threshold applied to the auxiliary population. This challenge does not exist in single-population MR analysis. If the selection bias from IV selection (based on the auxiliary population) and LD clumping (based on the target population) is not accounted for, type I error rates may not be properly controlled. To handle this, XMR introduces a specialized correction term, Δ*t*, to account for the population difference (details in Supplementary section 1.4). Neglecting this correction leads to inflated type I error rates, as evidenced by false positives in our negative control studies (Supplementary section 4.10, Fig. S27).

### Computation

The computation time for analyzing a single trait pair using XMR varies from under 3 minutes to over 1 hour, depending on data complexity. Specifically, parameter estimation for confounding factors typically requires 1–2 minutes. The runtime for the variational-EM algorithm depends on the number of IVs and the iterations required for convergence. For a pair with approximately 500 IVs, causal effect estimation is typically completed in less than 15 minutes on a Linux server equipped with an **Intel**^**®**^ **Xeon**^**®**^ **CPU E5-2699 v4 @ 2.20GHz**. For large-scale analyses involving many trait pairs, a parallel computing framework can be employed to maximize the utilization of available CPU cores, thereby achieving high scalability and significantly reducing the total runtime.

## Data Availability

All data produced are publicly available and listed at https://github.com/YangLabHKUST/XMR.

## Data and code availability

All GWAS summary statistics used in this paper are publicly available. The data sources are listed in the Experiment design section. The URLs for downloading the datasets are summarized in Supplementary Tables S4, S15, S20, and S23. XMR is implemented as an open-source R package. The XMR software, example datasets, and source code for reproducing the results are available at https://github.com/YangLabHKUST/XMR.

## Acknowledgements

This work was supported in part by the Innovation and Technology Commission (ITCPD/17-9); Research Grants Council, University Grants Committee (16301419, 16308120, 16307221, 16307322, 16302823, and 16309424); The Hong Kong University of Science and Technology Startup Grants (R9405 and Z0428 from the Big Data Institute); National Natural Science Foundation of China (NSFC) (Grant No. 12501401); Shenzhen Newly Introduced High-end Talents Scientific Research Start-up Project (Pengcheng Peacock Plan, Grant No. 892007202); and Scientific Foundation for Youth Scholars of Shenzhen University (Grant No. RC20240274).

## Author contributions

X. Huang, X.Hu, and C.Y. conceptualized and designed the study. X.Huang, Z.C., X.Hu, and C.Y. developed the methodology. X.Huang developed the software package. X.Huang performed the analysis with the assistance of Z.W. and X.Hu. X.Huang and C.Y. wrote the manuscript. Z.W., Z.C., and X.Hu provided critical feedback during the study and helped revise the manuscript.

