## Supplementary methods, notes, figures, and tables (S1-S3) for "XMR: A cross-population Mendelian randomization method for causal inference using genome-wide summary statistics"

### 1 Supplementary methods

Here, we provide a full version of our methods with fitting details.

#### 1.1 The XMR model

Let $\hat{\gamma}_{1,j}$, $\hat{\gamma}_{2,j}$ and $\hat{\Gamma}_{2,j}$ be the GWAS estimates of SNP $j$ for exposure in a large auxiliary population $X_1$, exposure in the target population $X_2$, and outcome in the target population $Y_2$, respectively, while $\hat{s}_{X_1,j}$, $\hat{s}_{X_2,j}$ and $\hat{s}_{Y_2,j}$ are their corresponding standard errors. XMR first conducts IV selection using $X_1$ with $|\frac{\hat{\gamma}_{1,j}}{\hat{s}_{X_1,j}}| \geq t$, which is followed by linkage disequilibrium (LD) clumping to further reduce dependence among remaining IVs. With the candidate IV set $\left\{\hat{\gamma}_{1,j}, \hat{\gamma}_{2,j}, \hat{\Gamma}_{2,j}, \hat{s}_{X_1,j}, \hat{s}_{X_2,j}, \hat{s}_{Y_2,j} \mid |\frac{\hat{\gamma}_{1,j}}{\hat{s}_{X_1,j}}| \geq t\right\}_{j=1,\ldots,M_t}$, XMR decomposes the observed SNP-trait associations into causal effects and confounding factors:

$$\begin{pmatrix}\hat{\gamma}_{1,j}\\ \hat{\gamma}_{2,j}\\ \hat{\Gamma}_{2,j}\end{pmatrix} = \underbrace{\underset{\text{IV validity}}{Z_j} \cdot \underset{\substack{\text{Uncorrelated pleiotropy}\\ \text{Causal effect}}}{\begin{pmatrix}\gamma_{1,j}\\ \gamma_{2,j}\\ \beta\gamma_{2,j}+\alpha_j\end{pmatrix}}}_{\text{Causal inference module}} + \underbrace{\underset{\text{Correlated pleiotropy}}{\begin{pmatrix}u_{1,j}\\ u_{2,j}\\ v_{2,j}\end{pmatrix}} + \underset{\text{Sample structure}}{\begin{pmatrix}\epsilon_{1,j}\\ \epsilon_{2,j}\\ \xi_{2,j}\end{pmatrix}}}_{\text{Confounding correction module}}, \tag{S1}$$

where $\gamma_{1,j}$ and $\gamma_{2,j}$ represent the true SNP effects on $X_1$ and $X_2$, and $\beta$ is the causal effect of interest. $Z_j$ is a binary variable indicating whether SNP $j$ is a valid IV ($Z_j = 1$) or not ($Z_j = 0$), with only valid ones used for causal effect estimation. The terms $(u_{1,j}, u_{2,j}, v_{2,j})$ and $(\epsilon_{1,j}, \epsilon_{2,j}, \xi_{2,j})$ model correlated pleiotropy and sample structure, respectively, while $\alpha_j$ denotes uncorrelated pleiotropy, altogether accounting for confounding factors.

#### 1.2 Estimation of correlated pleiotropy and sample structure

Under the assumptions of LD Score Regression (LDSC) [1], we are able to quantify and distinguish the effects of correlated pleiotropy and sample structure, as correlated pleiotropy associates with LD while sample structure doesn't.

We now specify distributional assumptions for both terms. Since LD influences genetic correlations, the pleiotropic effects $(u_{1,j}, u_{2,j}, v_{2,j})$ are assumed to follow a multivariate normal distribution whose covariance matrix scales with LD scores:

$$\begin{pmatrix}u_{1,j}\\ u_{2,j}\\ v_{2,j}\end{pmatrix} \sim \mathcal{N}\left(\mathbf{0}, \mathbf{\Omega}_j\right), \quad \text{where } \mathbf{\Omega}_j = \begin{pmatrix} l_{1j}\omega_1^2 & l_{12j}\omega_{12} & l_{12j}\omega_{1y}\\ l_{12j}\omega_{12} & l_{2j}\omega_2^2 & l_{2j}\omega_{2y}\\ l_{12j}\omega_{1y} & l_{2j}\omega_{2y} & l_{2j}\omega_y^2\end{pmatrix}. \tag{S2}$$

Here $\omega_1^2$, $\omega_2^2$ and $\omega_y^2$ are the variances of polygenic effects on $X_1$, $X_2$ and $Y_2$, respectively, while $\omega_{12}$, $\omega_{1y}$ and $\omega_{2y}$ quantify the genetic covariances between these traits. $l_{1j}$, $l_{2j}$ and $l_{12j}$ are the LD scores of SNP $j$, representing its correlation structure within the large-sample population, the small-sample population, and across the two populations, respectively.

Sample structure is independent of LD and modeled as:

$$\begin{pmatrix}\epsilon_{1,j}\\ \epsilon_{2,j}\\ \xi_{2,j}\end{pmatrix} \sim \mathcal{N}\left(\mathbf{0}, \hat{\boldsymbol{S}}_j \boldsymbol{C} \hat{\boldsymbol{S}}_j\right), \quad \text{where } \hat{\boldsymbol{S}}_j = \begin{pmatrix}\hat{s}_{X_1,j} & 0 & 0\\ 0 & \hat{s}_{X_2,j} & 0\\ 0 & 0 & \hat{s}_{Y_2,j}\end{pmatrix}, \boldsymbol{C} = \begin{pmatrix}c_1 & c_{12} & c_{1y}\\ c_{12} & c_2 & c_{2y}\\ c_{1y} & c_{2y} & c_y\end{pmatrix}. \quad \text{(S3)}$$

Here $\hat{\boldsymbol{S}}_j$ is a diagonal matrix of the standard errors $\hat{s}_{X_1,j}$, $\hat{s}_{X_2,j}$ and $\hat{s}_{Y_2,j}$, which accounts for the varying uncertainty across datasets. The matrix $\boldsymbol{C}$ describes residual correlations due to shared sample structure, with the diagonal elements $c_1$, $c_2$ and $c_y$ representing population-specific variance, and the off-diagonal elements $c_{12}$, $c_{1y}$ and $c_{2y}$ modeling cross-dataset correlations.

Both $\boldsymbol{\Omega}_j$ and $\boldsymbol{C}$ are pre-estimated from genome-wide summary statistics instead of just the $M_t$ SNPs passing IV selection criteria, denoted as $\hat{\boldsymbol{\Omega}}_j$ and $\hat{\boldsymbol{C}}$, respectively. For $\hat{\boldsymbol{\Omega}}_j$, the diagonal terms $\hat{\omega}_1^2$, $\hat{\omega}_2^2$ and $\hat{\omega}_y^2$ are the estimated per-SNP heritabilities from the slopes of the corresponding single-trait LDSC. The off-diagonal terms are the estimates of the per-SNP co-heritability obtained via bivariate LDSC, regarding the two corresponding traits. Similarly, for $\hat{\boldsymbol{C}}$, the diagonal terms $\hat{c}_1$, $\hat{c}_2$ and $\hat{c}_y$ are the intercepts estimated from single-trait LDSC, while off-diagonal terms $\hat{c}_{12}$, $\hat{c}_{1y}$ and $\hat{c}_{2y}$ are estimated by the intercepts of bivariate LDSC. The derivation procedure is an extension of that in SI Appendix, section 1.1 in MRAPSS [2]. We will fix these estimates when proceeding to the next fitting steps.

#### 1.3 Causal effect estimation

We adopt the InSIDE assumption for the causal inference module, where the direct effect $\alpha_j$ is assumed to be independent of the true SNP effects on $X_1$ and $X_2$, i.e., $\alpha_j \perp (\gamma_{1,j}, \gamma_{2,j})$. Accordingly,

$$\begin{pmatrix}\gamma_{1,j}\\ \gamma_{2,j}\\ \alpha_j\end{pmatrix} \sim \mathcal{N}(\mathbf{0}, \boldsymbol{\Sigma}), \text{with } \boldsymbol{\Sigma} = \begin{pmatrix}\sigma_{1,f}^2 & \sigma_{12,f} & 0\\ \sigma_{12,f} & \sigma_{2,f}^2 & 0\\ 0 & 0 & \tau^2\end{pmatrix}, \boldsymbol{\Sigma}_e = \begin{pmatrix}\sigma_{1,f}^2 & \sigma_{12,f}\\ \sigma_{12,f} & \sigma_{2,f}^2\end{pmatrix},$$

where $\mathbf{\Sigma}_e$ denotes the top-left $2 \times 2$ submatrix of $\mathbf{\Sigma}$. With $\boldsymbol{A}(\beta) = \begin{pmatrix} 1 & 0 & 0 \\ 0 & 1 & 0 \\ 0 & \beta & 1 \end{pmatrix}$, we have

$$\begin{pmatrix} \hat{\gamma}_{1,j} \\ \hat{\gamma}_{2,j} \\ \hat{\Gamma}_{2,j} \end{pmatrix} \Bigg| Z_j, \begin{pmatrix} \gamma_{1,j} \\ \gamma_{2,j} \\ \alpha_j \end{pmatrix} \sim \mathcal{N}\left( \begin{pmatrix} \hat{\gamma}_{1,j} \\ \hat{\gamma}_{2,j} \\ \hat{\Gamma}_{2,j} \end{pmatrix}; Z_j \boldsymbol{A}(\beta) \begin{pmatrix} \gamma_{1,j} \\ \gamma_{2,j} \\ \alpha_j \end{pmatrix}, \hat{\boldsymbol{\Omega}}_j + \hat{\boldsymbol{S}}_j \hat{\boldsymbol{C}} \hat{\boldsymbol{S}}_j \right).$$

To obtain the maximum likelihood estimate for $\beta$, we employ a variational EM algorithm. Firstly, we need to deal with the selection bias introduced by IV selection.

Defining $\pi_t = p(Z_j = 1 \mid |\hat{\gamma}_{1,j}/\hat{s}_{X_1,j}| \geq t)$, we have:

$$\begin{aligned} & p(\hat{\gamma}_{1,j}, \hat{\gamma}_{2,j}, \hat{\Gamma}_{2,j} \mid |\hat{\gamma}_{1,j}/\hat{s}_{X_1,j}| \geq t) \\ =& \pi_t p(\hat{\gamma}_{1,j}, \hat{\gamma}_{2,j}, \hat{\Gamma}_{2,j} \mid Z_j = 1, |\hat{\gamma}_{1,j}/\hat{s}_{X_1,j}| \geq t) + (1-\pi_t) p(\hat{\gamma}_{1,j}, \hat{\gamma}_{2,j}, \hat{\Gamma}_{2,j} \mid Z_j = 0, |\hat{\gamma}_{1,j}/\hat{s}_{X_1,j}| \geq t) \\ =& \pi_t \frac{\mathcal{N}(0, \boldsymbol{A}(\beta)\mathbf{\Sigma}\boldsymbol{A}(\beta)^T + \hat{\boldsymbol{\Omega}}_j + \hat{\boldsymbol{S}}_j \hat{\boldsymbol{C}} \hat{\boldsymbol{S}}_j)}{2\Phi\left(-\frac{t\hat{s}_{X_1,j}}{\sqrt{\sigma_{1,f}^2 + l_{1j}\hat{\omega}_1^2 + \hat{c}_1 \hat{s}_{X_1,j}^2}}\right)} + (1-\pi_t) \frac{\mathcal{N}(0, \hat{\boldsymbol{\Omega}}_j + \hat{\boldsymbol{S}}_j \hat{\boldsymbol{C}} \hat{\boldsymbol{S}}_j)}{2\Phi\left(-\frac{t\hat{s}_{X_1,j}}{\sqrt{l_{1j}\hat{\omega}_1^2 + \hat{c}_1 \hat{s}_{X_1,j}^2}}\right)}, \end{aligned} \tag{S4}$$

where $\Phi(\cdot)$ is the cumulative distribution function (CDF) of the standard normal distribution.

Let $\boldsymbol{\theta} = (\beta, \pi_t, \mathbf{\Sigma})$, $\hat{\boldsymbol{\gamma}}_1 = \{\hat{\gamma}_{1,j}\}_{j=1,\ldots,M_t}$, $\hat{\boldsymbol{\gamma}}_2 = \{\hat{\gamma}_{2,j}\}_{j=1,\ldots,M_t}$, $\hat{\mathbf{\Gamma}} = \{\hat{\Gamma}_{2,j}\}_{j=1,\ldots,M_t}$, $\boldsymbol{\gamma}_1 = \{\gamma_{1,j}\}_{j=1,\ldots,M_t}$, $\boldsymbol{\gamma}_2 = \{\gamma_{2,j}\}_{j=1,\ldots,M_t}$, $\boldsymbol{\alpha} = \{\alpha_j\}_{j=1,\ldots,M_t}$, and $\mathbf{Z} = \{Z_j\}_{j=1,\ldots,M_t}$. By treating $\boldsymbol{\gamma}_1, \boldsymbol{\gamma}_2, \boldsymbol{\alpha}$, and $\mathbf{Z}$ as latent variables, the complete data likelihood can be obtained as follows:

$$\begin{aligned} & p\left(\hat{\boldsymbol{\gamma}}_1, \hat{\boldsymbol{\gamma}}_2, \hat{\mathbf{\Gamma}}, \boldsymbol{\gamma}_1, \boldsymbol{\gamma}_2, \boldsymbol{\alpha}, \mathbf{Z} \mid t; \boldsymbol{\theta}\right) \\ =& \prod_{j=1}^{M_t} p\left(\hat{\gamma}_{1,j}, \hat{\gamma}_{2,j}, \hat{\Gamma}_{2,j}, \gamma_{1,j}, \gamma_{2,j}, \alpha_j \mid Z_j, |\hat{\gamma}_{1,j}/\hat{s}_{X_1,j}| \geq t; \boldsymbol{\theta}\right) p\left(Z_j \mid |\hat{\gamma}_{1,j}/\hat{s}_{X_1,j}| \geq t; \boldsymbol{\theta}\right) \\ =& \prod_{j=1}^{M_t} \frac{p\left(\hat{\gamma}_{1,j}, \hat{\gamma}_{2,j}, \hat{\Gamma}_{2,j}, \gamma_{1,j}, \gamma_{2,j}, \alpha_j \mid Z_j; \boldsymbol{\theta}\right)}{p\left(|\hat{\gamma}_{1,j}/\hat{s}_{X_1,j}| \geq t \mid Z_j; \boldsymbol{\theta}\right)} p\left(Z_j \mid |\hat{\gamma}_{1,j}/\hat{s}_{X_1,j}| \geq t; \boldsymbol{\theta}\right) \\ =& \prod_{j=1}^{M_t} \frac{p\left(\hat{\gamma}_{1,j}, \hat{\gamma}_{2,j}, \hat{\Gamma}_{2,j} \mid \gamma_{1,j}, \gamma_{2,j}, \alpha_j, Z_j; \boldsymbol{\theta}\right) p\left(\gamma_{1,j}, \gamma_{2,j}, \alpha_j \mid Z_j; \boldsymbol{\theta}\right)}{p\left(|\hat{\gamma}_{1,j}/\hat{s}_{X_1,j}| \geq t \mid Z_j; \boldsymbol{\theta}\right)} p\left(Z_j \mid |\hat{\gamma}_{1,j}/\hat{s}_{X_1,j}| \geq t; \boldsymbol{\theta}\right) \\ =& \prod_{j=1}^{M_t} \frac{\mathcal{N}\left(\begin{pmatrix} \hat{\gamma}_{1,j} \\ \hat{\gamma}_{2,j} \\ \hat{\Gamma}_{2,j} \end{pmatrix} \Bigg| Z_j \mathbf{A}(\beta) \begin{pmatrix} \gamma_{1,j} \\ \gamma_{2,j} \\ \alpha_j \end{pmatrix}, \hat{\mathbf{\Omega}}_j + \hat{\mathbf{S}}_j \hat{\mathbf{C}} \hat{\mathbf{S}}_j\right) \mathcal{N}\left(\begin{pmatrix} \gamma_{1,j} \\ \gamma_{2,j} \\ \alpha_j \end{pmatrix} \Bigg| \mathbf{0}, \mathbf{\Sigma}\right)}{\left(2\Phi\left(-\frac{t\hat{s}_{X_1,j}}{\sqrt{l_{1j}\hat{\omega}_1^2 + \hat{c}_1 \hat{s}_{X_1,j}^2}}\right)\right)^{1-Z_j} \left(2\Phi\left(-\frac{t\hat{s}_{X_1,j}}{\sqrt{\sigma_{1,f}^2 + l_{1j}\hat{\omega}_1^2 + \hat{c}_1 \hat{s}_{X_1,j}^2}}\right)\right)^{Z_j}} \pi_t^{Z_j} (1-\pi_t)^{1-Z_j}. \end{aligned}$$

The complete data log-likelihood is:

$$
\begin{aligned}
&\mathcal{L}_c(\boldsymbol{\theta}) \\
=&\sum_{j=1}^{M_t} \log \mathcal{N}\left(\left(\begin{matrix}\hat{\gamma}_{1,j}\\ \hat{\gamma}_{2,j}\\ \hat{\Gamma}_{2,j}\end{matrix}\right) \,\Big|\, Z_j\mathbf{A}(\beta)\left(\begin{matrix}\gamma_{1,j}\\ \gamma_{2,j}\\ \alpha_j\end{matrix}\right), \hat{\boldsymbol{\Omega}}_j + \hat{\mathbf{S}}_j\hat{\mathbf{C}}\hat{\mathbf{S}}_j\right) + \\
&\sum_{j=1}^{M_t} \log \mathcal{N}\left(\left(\begin{matrix}\gamma_{1,j}\\ \gamma_{2,j}\\ \alpha_j\end{matrix}\right) \,\Big|\, \mathbf{0}, \boldsymbol{\Sigma}\right) + Z_j \log \pi_t + (1-Z_j)\log(1-\pi_t) - \\
&\sum_{j=1}^{M_t} Z_j \log\left(2\Phi\left(-\frac{t\hat{s}_{X_1,j}}{\sqrt{\sigma_{1,f}^2 + l_{1j}\hat{\omega}_1^2 + \hat{c}_1\hat{s}_{X_1,j}^2}}\right)\right) - (1-Z_j)\log\left(2\Phi\left(-\frac{t\hat{s}_{X_1,j}}{\sqrt{l_{1j}\hat{\omega}_1^2 + \hat{c}_1\hat{s}_{X_1,j}^2}}\right)\right) \\
=&\sum_{j=1}^{M_t} -\frac{1}{2}\log\det(\hat{\mathbf{S}}_j\hat{\mathbf{C}}\hat{\mathbf{S}}_j + \hat{\boldsymbol{\Omega}}_j) - \\
&\sum_{j=1}^{M_t} \frac{1}{2}\left\{\left(\begin{matrix}\hat{\gamma}_{1,j}\\ \hat{\gamma}_{2,j}\\ \hat{\Gamma}_{2,j}\end{matrix}\right) - Z_j\mathbf{A}(\beta)\left(\begin{matrix}\gamma_{1,j}\\ \gamma_{2,j}\\ \alpha_j\end{matrix}\right)\right\}^T (\hat{\mathbf{S}}_j\hat{\mathbf{C}}\hat{\mathbf{S}}_j + \hat{\boldsymbol{\Omega}}_j)^{-1}\left\{\left(\begin{matrix}\hat{\gamma}_{1,j}\\ \hat{\gamma}_{2,j}\\ \hat{\Gamma}_{2,j}\end{matrix}\right) - Z_j\mathbf{A}(\beta)\left(\begin{matrix}\gamma_{1,j}\\ \gamma_{2,j}\\ \alpha_j\end{matrix}\right)\right\} + \\
&\sum_{j=1}^{M_t} -\frac{1}{2}\log\det(\boldsymbol{\Sigma}) - \frac{1}{2}\left(\begin{matrix}\gamma_{1,j}\\ \gamma_{2,j}\\ \alpha_j\end{matrix}\right)^T \boldsymbol{\Sigma}^{-1}\left(\begin{matrix}\gamma_{1,j}\\ \gamma_{2,j}\\ \alpha_j\end{matrix}\right) + Z_j\log\pi_t + (1-Z_j)\log(1-\pi_t) - \\
&\sum_{j=1}^{M_t} Z_j \log\left(2\Phi\left(-\frac{t\hat{s}_{X_1,j}}{\sqrt{\sigma_{1,f}^2 + l_{1j}\hat{\omega}_1^2 + \hat{c}_1\hat{s}_{X_1,j}^2}}\right)\right) - (1-Z_j)\log\left(2\Phi\left(-\frac{t\hat{s}_{X_1,j}}{\sqrt{l_{1j}\hat{\omega}_1^2 + \hat{c}_1\hat{s}_{X_1,j}^2}}\right)\right) + \\
&\text{constant.}
\end{aligned}
$$

Let $q(\boldsymbol{\gamma}_1, \boldsymbol{\gamma}_2, \boldsymbol{\alpha}, \mathbf{Z})$ be a variational distribution. The logarithm of the marginal likelihood can be written as

$$
\begin{aligned}
&\log p(\hat{\boldsymbol{\gamma}}_1, \hat{\boldsymbol{\gamma}}_2, \hat{\boldsymbol{\Gamma}} \mid \boldsymbol{\theta}, t) \\
=&\mathbb{E}_{q(\boldsymbol{\gamma}_1,\boldsymbol{\gamma}_2,\boldsymbol{\alpha},\mathbf{Z})}(\log p(\hat{\boldsymbol{\gamma}}_1, \hat{\boldsymbol{\gamma}}_2, \hat{\boldsymbol{\Gamma}} \mid \boldsymbol{\theta}, t)) \\
=&\mathbb{E}_{q(\boldsymbol{\gamma}_1,\boldsymbol{\gamma}_2,\boldsymbol{\alpha},\mathbf{Z})}\left(\log\frac{p(\hat{\boldsymbol{\gamma}}_1, \hat{\boldsymbol{\gamma}}_2, \hat{\boldsymbol{\Gamma}}, \boldsymbol{\gamma}_1, \boldsymbol{\gamma}_2, \boldsymbol{\alpha}, \mathbf{Z} \mid \boldsymbol{\theta}, t)}{p(\boldsymbol{\gamma}_1, \boldsymbol{\gamma}_2, \boldsymbol{\alpha}, \mathbf{Z} \mid \hat{\boldsymbol{\gamma}}_1, \hat{\boldsymbol{\gamma}}_2, \hat{\boldsymbol{\Gamma}}, \boldsymbol{\theta}, t)}\right) \\
=&\mathbb{E}_{q(\boldsymbol{\gamma}_1,\boldsymbol{\gamma}_2,\boldsymbol{\alpha},\mathbf{Z})}\left(\log\frac{p(\hat{\boldsymbol{\gamma}}_1, \hat{\boldsymbol{\gamma}}_2, \hat{\boldsymbol{\Gamma}}, \boldsymbol{\gamma}_1, \boldsymbol{\gamma}_2, \boldsymbol{\alpha}, \mathbf{Z} \mid \boldsymbol{\theta}, t)}{q(\boldsymbol{\gamma}_1, \boldsymbol{\gamma}_2, \boldsymbol{\alpha}, \mathbf{Z})} - \log\frac{p(\boldsymbol{\gamma}_1, \boldsymbol{\gamma}_2, \boldsymbol{\alpha}, \mathbf{Z} \mid \hat{\boldsymbol{\gamma}}_1, \hat{\boldsymbol{\gamma}}_2, \hat{\boldsymbol{\Gamma}}, \boldsymbol{\theta}, t)}{q(\boldsymbol{\gamma}_1, \boldsymbol{\gamma}_2, \boldsymbol{\alpha}, \mathbf{Z})}\right) \\
=&\mathcal{L}(q; \boldsymbol{\theta}, t) + \mathrm{D}_{\mathrm{KL}}\left(q(\boldsymbol{\gamma}_1, \boldsymbol{\gamma}_2, \boldsymbol{\alpha}, \mathbf{Z}) \parallel p(\boldsymbol{\gamma}_1, \boldsymbol{\gamma}_2, \boldsymbol{\alpha}, \mathbf{Z} \mid \hat{\boldsymbol{\gamma}}_1, \hat{\boldsymbol{\gamma}}_2, \hat{\boldsymbol{\Gamma}}, \boldsymbol{\theta}, t)\right),
\end{aligned}
$$

where

$$
\mathcal{L}(q; \boldsymbol{\theta}, t) = \mathbb{E}_{q(\boldsymbol{\gamma}_1,\boldsymbol{\gamma}_2,\boldsymbol{\alpha},\mathbf{Z})}\left(\log\frac{p(\hat{\boldsymbol{\gamma}}_1, \hat{\boldsymbol{\gamma}}_2, \hat{\boldsymbol{\Gamma}}, \boldsymbol{\gamma}_1, \boldsymbol{\gamma}_2, \boldsymbol{\alpha}, \mathbf{Z}|\boldsymbol{\theta}, t)}{q(\boldsymbol{\gamma}_1, \boldsymbol{\gamma}_2, \boldsymbol{\alpha}, \mathbf{Z})}\right),
$$

$$
\mathrm{D}_{\mathrm{KL}}\left(q(\boldsymbol{\gamma}_1, \boldsymbol{\gamma}_2, \boldsymbol{\alpha}, \mathbf{Z}) \parallel p(\boldsymbol{\gamma}_1, \boldsymbol{\gamma}_2, \boldsymbol{\alpha}, \mathbf{Z} \mid \hat{\boldsymbol{\gamma}}_1, \hat{\boldsymbol{\gamma}}_2, \hat{\boldsymbol{\Gamma}}, \boldsymbol{\theta}, t)\right) = -\,\mathbb{E}_{q(\boldsymbol{\gamma}_1,\boldsymbol{\gamma}_2,\boldsymbol{\alpha},\mathbf{Z})}\left(\log\frac{p(\boldsymbol{\gamma}_1, \boldsymbol{\gamma}_2, \boldsymbol{\alpha}, \mathbf{Z} \mid \hat{\boldsymbol{\gamma}}_1, \hat{\boldsymbol{\gamma}}_2, \hat{\boldsymbol{\Gamma}}, \boldsymbol{\theta}, t)}{q(\boldsymbol{\gamma}_1, \boldsymbol{\gamma}_2, \boldsymbol{\alpha}, \mathbf{Z})}\right).
$$

Since the Kullback-Leibler (KL) divergence $\mathrm{D}_{\mathrm{KL}}\left(q(\boldsymbol{\gamma}_1, \boldsymbol{\gamma}_2, \boldsymbol{\alpha}, \mathbf{Z}) \parallel p(\boldsymbol{\gamma}_1, \boldsymbol{\gamma}_2, \boldsymbol{\alpha}, \mathbf{Z} \mid \hat{\gamma}_1, \hat{\gamma}_2, \hat{\boldsymbol{\Gamma}}, \boldsymbol{\theta}, t)\right)$ is non-negative, $\mathcal{L}(q; \boldsymbol{\theta}, t)$ is the evidence lower bound (ELBO) of the marginal log-likelihood $\log p(\hat{\boldsymbol{\gamma}}_1, \hat{\boldsymbol{\gamma}}_2, \hat{\boldsymbol{\Gamma}} | \boldsymbol{\theta}, t)$. Thus, maximization of $\mathcal{L}(q; \boldsymbol{\theta}, t)$ w.r.t. variational distribution $q$ and parameter $\boldsymbol{\theta}$ follows the EM framework: in the E-step, variational distribution $q$ is updated to approximate the true posterior; in the M-step, parameters in $\boldsymbol{\theta}$ are optimized to increase the ELBO.

**E-step.** To make it feasible for evaluation of the lower bound $\mathcal{L}(q; \boldsymbol{\theta}, t)$, we adopt the mean-field assumption that the variational distribution $q(\boldsymbol{\gamma}_1, \boldsymbol{\gamma}_2, \boldsymbol{\alpha}, \mathbf{Z})$ can be factorized as:

$$q(\boldsymbol{\gamma}_1, \boldsymbol{\gamma}_2, \boldsymbol{\alpha}, \mathbf{Z}) = \prod_{j=1}^{M_t} q(\gamma_{1,j}, \gamma_{2,j}, \alpha_j, Z_j) = \prod_{j=1}^{M_t} q(\gamma_{1,j}, \gamma_{2,j}, \alpha_j \mid Z_j) q(Z_j). \tag{S5}$$

Noting that $Z_j$ is a binary variable, we define

$$q(Z_j) = \eta_j^{Z_j} (1 - \eta_j)^{(1 - Z_j)}, \quad \text{where } \eta_j = q(Z_j = 1). \tag{S6}$$

Based on the mean-field approximation, we can derive the optimal solutions for the $q$ distribution in Eq. (S5) at each step. We first obtain the optimal solution for $q(\gamma_{1,j}, \gamma_{2,j}, \alpha_j \mid Z_j)$, for $j = 1, \ldots, M_t$. Given $Z_j = 1$, we have

$$\log q(\gamma_{1,j}, \gamma_{2,j}, \alpha_j \mid Z_j = 1) = \mathbb{E}_{q_{-j}} \left( \log p(\hat{\boldsymbol{\gamma}}_1, \hat{\boldsymbol{\gamma}}_2, \hat{\boldsymbol{\Gamma}}, \boldsymbol{\gamma}_1, \boldsymbol{\gamma}_2, \boldsymbol{\alpha}, \mathbf{Z} \mid \boldsymbol{\theta}, t) \right) + \text{constant},$$

where $\mathbb{E}_{q_{-j}}$ denotes the expectation w.r.t. the $q$ distribution over $(\boldsymbol{\gamma}_1, \boldsymbol{\gamma}_2, \boldsymbol{\alpha})$ except $(\gamma_{1,j}, \gamma_{2,j}, \alpha_j)$, conditioning on $Z_j = 1$. Thus, we have

$$\begin{aligned} &\log q(\gamma_{1,j}, \gamma_{2,j}, \alpha_j \mid Z_j = 1) \\ &= -\frac{1}{2} \left\{ \begin{pmatrix} \hat{\gamma}_{1,j} \\ \hat{\gamma}_{2,j} \\ \hat{\Gamma}_{2,j} \end{pmatrix} - \mathbf{A}(\beta) \begin{pmatrix} \gamma_{1,j} \\ \gamma_{2,j} \\ \alpha_j \end{pmatrix} \right\}^T (\hat{\mathbf{S}}_j \hat{\mathbf{C}} \hat{\mathbf{S}}_j + \hat{\boldsymbol{\Omega}}_j)^{-1} \left\{ \begin{pmatrix} \hat{\gamma}_{1,j} \\ \hat{\gamma}_{2,j} \\ \hat{\Gamma}_{2,j} \end{pmatrix} - \mathbf{A}(\beta) \begin{pmatrix} \gamma_{1,j} \\ \gamma_{2,j} \\ \alpha_j \end{pmatrix} \right\} \\ &\quad - \frac{1}{2} \begin{pmatrix} \gamma_{1,j} \\ \gamma_{2,j} \\ \alpha_j \end{pmatrix}^T \boldsymbol{\Sigma}^{-1} \begin{pmatrix} \gamma_{1,j} \\ \gamma_{2,j} \\ \alpha_j \end{pmatrix} + \text{constant}. \end{aligned}$$

We observe that the right hand side of the above expression is a quadratic function of $(\gamma_{1,j}, \gamma_{2,j}, \alpha_j)$, and we can identify $q(\gamma_{1,j}, \gamma_{2,j}, \alpha_j \mid Z_j = 1)$ as a Gaussian distribution:

$$q(\gamma_{1,j}, \gamma_{2,j}, \alpha_j \mid Z_j = 1) = \mathcal{N}\left( \begin{pmatrix} \gamma_{1,j} \\ \gamma_{2,j} \\ \alpha_j \end{pmatrix} \middle| \boldsymbol{\mu}_j, \boldsymbol{\Lambda}_j^{-1} \right), \tag{S7}$$

where

$$\begin{aligned} \boldsymbol{\Lambda}_j &= \mathbf{A}(\beta)^T (\hat{\mathbf{S}}_j \hat{\mathbf{C}} \hat{\mathbf{S}}_j + \hat{\boldsymbol{\Omega}}_j)^{-1} \mathbf{A}(\beta) + \boldsymbol{\Sigma}^{-1}, \\ \boldsymbol{\mu}_j &= \boldsymbol{\Lambda}_j^{-1} \mathbf{A}(\beta)^T (\hat{\mathbf{S}}_j \hat{\mathbf{C}} \hat{\mathbf{S}}_j + \hat{\boldsymbol{\Omega}}_j)^{-1} \begin{pmatrix} \hat{\gamma}_{1,j} \\ \hat{\gamma}_{2,j} \\ \hat{\Gamma}_{2,j} \end{pmatrix}. \end{aligned}$$

Similarly, the optimal solution for $q(\gamma_{1,j}, \gamma_{2,j}, \alpha_j \mid Z_j = 0)$ is given by

$$\log q(\gamma_{1,j}, \gamma_{2,j}, \alpha_j \mid Z_j = 0) = -\frac{1}{2} \begin{pmatrix} \gamma_{1,j} \\ \gamma_{2,j} \\ \alpha_j \end{pmatrix}^T \boldsymbol{\Sigma}^{-1} \begin{pmatrix} \gamma_{1,j} \\ \gamma_{2,j} \\ \alpha_j \end{pmatrix} + \text{constant}.$$

Thus, we have

$$q(\gamma_{1,j}, \gamma_{2,j}, \alpha_j \mid Z_j = 0) = \mathcal{N}\left( \begin{pmatrix} \gamma_{1,j} \\ \gamma_{2,j} \\ \alpha_j \end{pmatrix} \middle| \, \mathbf{0}, \boldsymbol{\Sigma} \right). \tag{S8}$$

Combining Eqs. (S5), (S6), (S7), and (S8), we have

$$q(\gamma_{1,j}, \gamma_{2,j}, \alpha_j, Z_j) = \left[ \eta_j \mathcal{N}\left( \begin{pmatrix} \gamma_{1,j} \\ \gamma_{2,j} \\ \alpha_j \end{pmatrix} \middle| \, \boldsymbol{\mu}_j, \boldsymbol{\Lambda}_j^{-1} \right) \right]^{Z_j} \left[ (1 - \eta_j) \mathcal{N}\left( \begin{pmatrix} \gamma_{1,j} \\ \gamma_{2,j} \\ \alpha_j \end{pmatrix} \middle| \, \mathbf{0}, \boldsymbol{\Sigma} \right) \right]^{1-Z_j}.$$

Once the variational distribution $q(\gamma_{1,j}, \gamma_{2,j}, \alpha_j, Z_j)$ is obtained, we can evaluate the ELBO:

$$\begin{aligned} \mathcal{L}(q; \boldsymbol{\theta}, t) =& \mathbb{E}_{q(\boldsymbol{\gamma}_1, \boldsymbol{\gamma}_2, \boldsymbol{\alpha}, \mathbf{Z})} \left( \log \frac{p(\hat{\boldsymbol{\gamma}}_1, \hat{\boldsymbol{\gamma}}_2, \hat{\boldsymbol{\Gamma}}, \boldsymbol{\gamma}_1, \boldsymbol{\gamma}_2, \boldsymbol{\alpha}, \mathbf{Z} \mid \boldsymbol{\theta}, t)}{q(\boldsymbol{\gamma}_1, \boldsymbol{\gamma}_2, \boldsymbol{\alpha}, \mathbf{Z})} \right) \\ =& \mathbb{E}_q \log p(\hat{\boldsymbol{\gamma}}_1, \hat{\boldsymbol{\gamma}}_2, \hat{\boldsymbol{\Gamma}}, \boldsymbol{\gamma}_1, \boldsymbol{\gamma}_2, \boldsymbol{\alpha}, \mathbf{Z} \mid \boldsymbol{\theta}, t) - \mathbb{E}_q \log q(\boldsymbol{\gamma}_1, \boldsymbol{\gamma}_2, \boldsymbol{\alpha}, \mathbf{Z}), \end{aligned}$$

where

$$\begin{aligned} & \mathbb{E}_q \log p(\hat{\boldsymbol{\gamma}}_1, \hat{\boldsymbol{\gamma}}_2, \hat{\boldsymbol{\Gamma}}, \boldsymbol{\gamma}_1, \boldsymbol{\gamma}_2, \boldsymbol{\alpha}, \mathbf{Z} \mid \boldsymbol{\theta}, t) \\ =& \sum_{j=1}^{M_t} \eta_j \widetilde{\boldsymbol{\mu}}_j^T \mathbf{A}(\beta)^T (\hat{\mathbf{S}}_j \hat{\mathbf{C}} \hat{\mathbf{S}}_j + \hat{\boldsymbol{\Omega}}_j)^{-1} \begin{pmatrix} \hat{\gamma}_{1,j} \\ \hat{\gamma}_{2,j} \\ \hat{\Gamma}_{2,j} \end{pmatrix} - \\ & \sum_{j=1}^{M_t} \frac{1}{2} \eta_j \operatorname{Tr} \left[ \mathbf{A}(\beta)^T (\hat{\mathbf{S}}_j \hat{\mathbf{C}} \hat{\mathbf{S}}_j + \hat{\boldsymbol{\Omega}}_j)^{-1} \mathbf{A}(\beta) (\widetilde{\boldsymbol{\Lambda}}_j^{-1} + \widetilde{\boldsymbol{\mu}}_j \widetilde{\boldsymbol{\mu}}_j^T) \right] + \\ & \sum_{j=1}^{M_t} -\frac{1}{2} \log \det(\boldsymbol{\Sigma}) - \frac{1}{2} \eta_j \widetilde{\boldsymbol{\mu}}_j^T \boldsymbol{\Sigma}^{-1} \widetilde{\boldsymbol{\mu}}_j - \frac{1}{2} \operatorname{Tr} \left[ \eta_j \boldsymbol{\Sigma}^{-1} \widetilde{\boldsymbol{\Lambda}}_j^{-1} \right] - \frac{1}{2} (1 - \eta_j) \operatorname{Tr} \left[ \boldsymbol{\Sigma}^{-1} \widetilde{\boldsymbol{\Sigma}} \right] + \\ & \sum_{j=1}^{M_t} \eta_j \log \pi_t + (1 - \eta_j) \log(1 - \pi_t) - \\ & \sum_{j=1}^{M_t} \eta_j \log \left( 2\Phi \left( - \frac{t \hat{s}_{X_1,j}}{\sqrt{\sigma_{1,f}^2 + l_{1j} \hat{\omega}_1^2 + \hat{c}_1 \hat{s}_{X_1,j}^2}} \right) \right) - (1 - \eta_j) \log \left( 2\Phi \left( - \frac{t \hat{s}_{X_1,j}}{\sqrt{l_{1j} \hat{\omega}_1^2 + \hat{c}_1 \hat{s}_{X_1,j}^2}} \right) \right) + \text{constant}, \end{aligned}$$

and

$$
\begin{aligned}
&-\mathbb{E}_q \log q(\boldsymbol{\gamma}_1, \boldsymbol{\gamma}_2, \boldsymbol{\alpha}, \mathbf{Z}) \\
=&\sum_{j=1}^{M_t} \frac{1}{2}\eta_j \log\det(\widetilde{\boldsymbol{\Lambda}}_j^{-1}) + \frac{1}{2}(1-\eta_j)\log\det(\widetilde{\boldsymbol{\Sigma}}) - \eta_j \log\eta_j - (1-\eta_j)\log(1-\eta_j) + \frac{1}{2}\eta_j \operatorname{Tr}\left[\boldsymbol{\Lambda}_j \widetilde{\boldsymbol{\Lambda}}_j^{-1}\right] + \\
&\sum_{j=1}^{M_t} \frac{1}{2}(1-\eta_j)\operatorname{Tr}\left[\widetilde{\boldsymbol{\Sigma}}^{-1}\widetilde{\boldsymbol{\Sigma}}\right] + \text{constant} \\
=&\sum_{j=1}^{M_t} \frac{1}{2}\eta_j \log\det(\widetilde{\boldsymbol{\Lambda}}_j^{-1}) + \frac{1}{2}(1-\eta_j)\log\det(\widetilde{\boldsymbol{\Sigma}}) - \eta_j \log\eta_j - (1-\eta_j)\log(1-\eta_j) + \text{constant}.
\end{aligned}
$$

Here, $\widetilde{\boldsymbol{\mu}}_j$, $\widetilde{\boldsymbol{\Lambda}}_j$, and $\widetilde{\boldsymbol{\Sigma}}$ denote the corresponding statistics $\boldsymbol{\mu}_j$, $\boldsymbol{\Lambda}_j$, and $\boldsymbol{\Sigma}$ in Eqs. (S7) and (S8) derived from expectations with respect to the variational posterior distribution $q$.

By maximizing $\mathcal{L}(q;\boldsymbol{\theta},t)$ w.r.t. $\eta_j$, where $\widetilde{\boldsymbol{\mu}}_j = \boldsymbol{\mu}_j$, $\widetilde{\boldsymbol{\Lambda}}_j = \boldsymbol{\Lambda}_j$ and $\widetilde{\boldsymbol{\Sigma}} = \boldsymbol{\Sigma}$ , we obtain

$$
\eta_j = \frac{1}{1+\exp(-\mathbf{b}_j)},
$$

where

$$
\mathbf{b}_j = \frac{1}{2}\boldsymbol{\mu}_j^T \boldsymbol{\Lambda}_j \boldsymbol{\mu}_j + \log\frac{\pi_t}{1-\pi_t} + \frac{1}{2}\log\frac{\det(\boldsymbol{\Lambda}_j^{-1})}{\det(\boldsymbol{\Sigma})} - \log\frac{\Phi\left(-\frac{t\hat{s}_{X_1,j}}{\sqrt{\sigma_{1,f}^2 + l_{1j}\hat{\omega}_1^2 + \hat{c}_1\hat{s}_{X_1,j}^2}}\right)}{\Phi\left(-\frac{t\hat{s}_{X_1,j}}{\sqrt{l_{1j}\hat{\omega}_1^2 + \hat{c}_1\hat{s}_{X_1,j}^2}}\right)}.
$$

**M-step.** We derive the updating equations for parameters $\beta$, $\pi_t$, $\tau^2$, and $\boldsymbol{\Sigma}_e$. Here, $\widetilde{\boldsymbol{\mu}}_j$, $\widetilde{\boldsymbol{\Lambda}}_j$, and $\widetilde{\boldsymbol{\Sigma}}$ are computed using the parameter estimates from the previous EM iteration. We first derive the updating equation for $\beta$. The terms in $\mathcal{L}(q;\boldsymbol{\theta},t)$ involving $\beta$ are

$$
\begin{aligned}
\mathcal{L}(\beta) =& \sum_{j=1}^{M_t} \eta_j \widetilde{\boldsymbol{\mu}}_j^T \mathbf{A}(\beta)^T (\hat{\mathbf{S}}_j \hat{\mathbf{C}} \hat{\mathbf{S}}_j + \hat{\boldsymbol{\Omega}}_j)^{-1} \begin{pmatrix} \hat{\gamma}_{1,j} \\ \hat{\gamma}_{2,j} \\ \hat{\Gamma}_{2,j} \end{pmatrix} - \\
& \sum_{j=1}^{M_t} \frac{1}{2}\eta_j \operatorname{Tr}\left[\mathbf{A}(\beta)^T (\hat{\mathbf{S}}_j \hat{\mathbf{C}} \hat{\mathbf{S}}_j + \hat{\boldsymbol{\Omega}}_j)^{-1} \mathbf{A}(\beta)(\widetilde{\boldsymbol{\Lambda}}_j^{-1} + \widetilde{\boldsymbol{\mu}}_j \widetilde{\boldsymbol{\mu}}_j^T)\right].
\end{aligned}
$$

Here we write $\boldsymbol{A}(\beta) = \begin{pmatrix} 1 & 0 & 0 \\ 0 & 1 & 0 \\ 0 & \beta & 1 \end{pmatrix}$, as $\boldsymbol{A}(\beta) = \mathbf{I}_3 + \beta\mathbf{V}_1$, where $\mathbf{I}_3 = \begin{pmatrix} 1 & 0 & 0 \\ 0 & 1 & 0 \\ 0 & 0 & 1 \end{pmatrix}$, and $\mathbf{V}_1 = \begin{pmatrix} 0 & 0 & 0 \\ 0 & 0 & 0 \\ 0 & 1 & 0 \end{pmatrix}$. Taking the derivative of $\mathcal{L}(\beta)$ w.r.t. $\beta$ and setting it to zero, the updating equation for $\beta$ is given as

$$
\beta = \frac{\sum_{j=1}^{M_t} \eta_j \widetilde{\boldsymbol{\mu}}_j^T \mathbf{V}_1^T (\hat{\mathbf{S}}_j \hat{\mathbf{C}} \hat{\mathbf{S}}_j + \hat{\boldsymbol{\Omega}}_j)^{-1} \begin{pmatrix} \hat{\gamma}_{1,j} \\ \hat{\gamma}_{2,j} \\ \hat{\Gamma}_{2,j} \end{pmatrix} - \eta_j \operatorname{Tr}(\mathbf{V}_1^T (\hat{\mathbf{S}}_j \hat{\mathbf{C}} \hat{\mathbf{S}}_j + \hat{\boldsymbol{\Omega}}_j)^{-1} (\widetilde{\boldsymbol{\Lambda}}_j^{-1} + \widetilde{\boldsymbol{\mu}}_j \widetilde{\boldsymbol{\mu}}_j^T))}{\sum_{j=1}^{M_t} \eta_j \operatorname{Tr}\left[\mathbf{V}_1^T (\hat{\mathbf{S}}_j \hat{\mathbf{C}} \hat{\mathbf{S}}_j + \hat{\boldsymbol{\Omega}}_j)^{-1} \mathbf{V}_1 (\widetilde{\boldsymbol{\Lambda}}_j^{-1} + \widetilde{\boldsymbol{\mu}}_j \widetilde{\boldsymbol{\mu}}_j^T)\right]}. \tag{S9}
$$

We next derive the updating equation for $\pi_t$. The terms in $\mathcal{L}(q;\boldsymbol{\theta},t)$ involving $\pi_t$ are

$$\mathcal{L}(\pi_t) = \sum_{j=1}^{M_t} \eta_j \log \pi_t + (1-\eta_j)\log(1-\pi_t).$$

By setting the derivative of $\mathcal{L}(\pi_t)$ w.r.t. $\pi_t$ to zero, we obtain

$$\pi_t = \frac{\sum_{j=1}^{M_t} \eta_j}{M_t}. \tag{S10}$$

We then derive the updating equation for $\tau^2$. Denote $\widetilde{\boldsymbol{\mu}}_j := (\mu_{\gamma_{1,j}}, \mu_{\gamma_{2,j}}, \mu_{\alpha_j})^T$ and the diagonal elements in $\widetilde{\boldsymbol{\Lambda}}_j^{-1}$ by $(\sigma^2_{\gamma_{1,j}}, \sigma^2_{\gamma_{2,j}}, \sigma^2_{\alpha_j})$. The terms in $\mathcal{L}(q;\boldsymbol{\theta},t)$ involving $\tau^2$ are given as

$$\mathcal{L}(\tau^2) = -\frac{1}{2}\sum_{j=1}^{M_t} \log\tau^2 - \frac{1}{2}\sum_{j=1}^{M_t} \eta_j \frac{\mu^2_{\alpha_j} + \sigma^2_{\alpha_j}}{\tau^2} - \frac{1}{2}\sum_{j=1}^{M_t}(1-\eta_j)\frac{\widetilde{\tau}^2}{\tau^2},$$

where $\widetilde{\tau}^2$ denotes the estimate of $\tau^2$ obtained in the previous EM iteration. Therefore, we obtain the updating equation for $\tau^2$ as

$$\tau^2 = \frac{\sum_{j=1}^{M_t} \eta_j(\mu^2_{\alpha_j} + \sigma^2_{\alpha_j}) + (1-\eta_j)\widetilde{\tau}^2}{M_t}.$$

Finally, we derive the update for $\boldsymbol{\Sigma}_e$. The terms in $\mathcal{L}(q;\boldsymbol{\theta},t)$ involving $\boldsymbol{\Sigma}_e$ are

$$\begin{aligned}\mathcal{L}(\boldsymbol{\Sigma}_e) = &\sum_{j=1}^{M_t} -\frac{1}{2}\log|\boldsymbol{\Sigma}_e| - \frac{1}{2}\eta_j \widetilde{\boldsymbol{\mu}}_{ej}^T \boldsymbol{\Sigma}_e^{-1} \widetilde{\boldsymbol{\mu}}_{ej} - \frac{1}{2}\operatorname{Tr}\left[\boldsymbol{\Sigma}_e^{-1}\left(\eta_j \widetilde{\boldsymbol{\Lambda}}_{ej}^{-1} + (1-\eta_j)\widetilde{\boldsymbol{\Sigma}}_e\right)\right] - \\ &\sum_{j=1}^{M_t} \eta_j \log\left(2\Phi\left(-\frac{t\hat{s}_{X_1,j}}{\sqrt{\sigma^2_{1,f} + l_{1j}\hat{\omega}_1^2 + \hat{c}_1\hat{s}^2_{X_1,j}}}\right)\right),\end{aligned}$$

where $\widetilde{\boldsymbol{\mu}}_{ej} := (\mu_{\gamma_{1,j}}, \mu_{\gamma_{2,j}})^T$, $\widetilde{\boldsymbol{\Lambda}}_{ej}^{-1}$ denotes the top-left $2\times 2$ submatrix of $\widetilde{\boldsymbol{\Lambda}}_j^{-1}$, and $\widetilde{\boldsymbol{\Sigma}}_e$ represents the value of $\boldsymbol{\Sigma}_e$ from the previous iteration.

If $t = 0$, we directly set the derivative of $\mathcal{L}(\boldsymbol{\Sigma}_e)$ w.r.t. $\boldsymbol{\Sigma}_e$ to zero and obtain the update for $\boldsymbol{\Sigma}_e$:

$$\boldsymbol{\Sigma}_e = \frac{\sum_{j=1}^{M_t} \eta_j(\widetilde{\boldsymbol{\mu}}_{ej}\widetilde{\boldsymbol{\mu}}_{ej}^T + \widetilde{\boldsymbol{\Lambda}}_{ej}^{-1}) + (1-\eta_j)\widetilde{\boldsymbol{\Sigma}}_e}{M_t}.$$

If $t \neq 0$, directly taking a derivative to obtain the optimum solution is difficult. To fix this, we first obtain a bound of $\mathcal{L}(\boldsymbol{\Sigma}_e)$, which is given by,

$$\begin{aligned}\mathcal{L}(\boldsymbol{\Sigma}_e) \geq &\sum_{j=1}^{M_t} -\frac{1}{2}\log\det(\boldsymbol{\Sigma}_e^{(t)}) - \frac{1}{2}\operatorname{Tr}\left(\left(\boldsymbol{\Sigma}_e^{(t)}\right)^{-1}(\boldsymbol{\Sigma}_e - \boldsymbol{\Sigma}_e^{(t)})\right) - \\ &\sum_{j=1}^{M_t} \frac{1}{2}\eta_j \widetilde{\boldsymbol{\mu}}_{ej}^T \boldsymbol{\Sigma}_e^{-1} \widetilde{\boldsymbol{\mu}}_{ej} - \frac{1}{2}\operatorname{Tr}\left[\boldsymbol{\Sigma}_e^{-1}\left(\eta_j \widetilde{\boldsymbol{\Lambda}}_{ej}^{-1} + (1-\eta_j)\widetilde{\boldsymbol{\Sigma}}_e\right)\right] - \\ &\sum_{j=1}^{M_t} \frac{1}{2}\eta_j t\hat{s}_{X_1,j} \frac{\phi\left(-\frac{t\hat{s}_{X_1,j}}{\sqrt{\sigma^{2\ (t)}_{1,f} + l_{1j}\hat{\omega}_1^2 + \hat{c}_1\hat{s}^2_{X_1,j}}}\right)}{\Phi\left(-\frac{t\hat{s}_{X_1,j}}{\sqrt{\sigma^{2\ (t)}_{1,f} + l_{1j}\hat{\omega}_1^2 + \hat{c}_1\hat{s}^2_{X_1,j}}}\right)} (\sigma^{2\ (t)}_{1,f} + l_{1j}\hat{\omega}_1^2 + \hat{c}_1\hat{s}^2_{X_1,j})^{-3/2}\mathbf{e}_1^T(\boldsymbol{\Sigma}_e - \boldsymbol{\Sigma}_e^{(t)})\mathbf{e}_1,\end{aligned}$$

where $\phi(\cdot)$ denotes the probability density function (PDF) of the standard normal distribution; $\mathbf{e}_1^T = \begin{pmatrix}1 & 0\end{pmatrix}$; ${\sigma^2_{1,f}}^{(t)}$ corresponds to the first diagonal element of $\mathbf{\Sigma}_e^{(t)}$; and $\mathbf{\Sigma}_e^{(t)}$ is treated as a fixed term in this step. Here we set $\mathbf{\Sigma}_e^{(t)} = \widetilde{\mathbf{\Sigma}}_e$.

Instead of maximizing $\mathcal{L}(\mathbf{\Sigma}_e)$, we maximize its lower bound and we have

$$\sum_{j=1}^{M_t} -\frac{1}{2}\widetilde{\mathbf{\Sigma}}_e^{-1} + \sum_{j=1}^{M_t} \frac{1}{2}\mathbf{\Sigma}_e^{-1}\left(\eta_j\left(\widetilde{\boldsymbol{\mu}}_{ej}\widetilde{\boldsymbol{\mu}}_{ej}^T + \widetilde{\mathbf{\Lambda}}_{ej}^{-1}\right) + (1-\eta_j)\widetilde{\mathbf{\Sigma}}_e\right)\mathbf{\Sigma}_e^{-1} -$$

$$\sum_{j=1}^{M_t} \frac{1}{2}\eta_j t\hat{s}_{X_1,j} \frac{\phi\left(-\frac{t\hat{s}_{X_1,j}}{\sqrt{\widetilde{\sigma}^2_{1,f}+l_{1j}\hat{\omega}_1^2+\hat{c}_1\hat{s}^2_{X_1,j}}}\right)}{\Phi\left(-\frac{t\hat{s}_{X_1,j}}{\sqrt{\widetilde{\sigma}^2_{1,f}+l_{1j}\hat{\omega}_1^2+\hat{c}_1\hat{s}^2_{X_1,j}}}\right)} (\widetilde{\sigma}^2_{1,f} + l_{1j}\hat{\omega}_1^2 + \hat{c}_1\hat{s}^2_{X_1,j})^{-3/2}\mathbf{e}_1\mathbf{e}_1^T = 0,$$

where $\widetilde{\sigma}^2_{1,f}$ denotes the $(1,1)$ element of $\widetilde{\mathbf{\Sigma}}_e$.

Let $\mathbf{L}$ be the Cholesky factor of $\mathbf{B} = \sum_{j=1}^{M_t} \widetilde{\mathbf{\Sigma}}_e^{-1} + \eta_j t\hat{s}_{X_1,j} \frac{\phi\left(-\frac{t\hat{s}_{X_1,j}}{\sqrt{\widetilde{\sigma}^2_{1,f}+l_{1j}\hat{\omega}_1^2+\hat{c}_1\hat{s}^2_{X_1,j}}}\right)}{\Phi\left(-\frac{t\hat{s}_{X_1,j}}{\sqrt{\widetilde{\sigma}^2_{1,f}+l_{1j}\hat{\omega}_1^2+\hat{c}_1\hat{s}^2_{X_1,j}}}\right)} (\widetilde{\sigma}^2_{1,f} + l_{1j}\hat{\omega}_1^2 + \hat{c}_1\hat{s}^2_{X_1,j})^{-3/2}\mathbf{e}_1\mathbf{e}_1^T$ such that $\mathbf{B} = \mathbf{L}\mathbf{L}^T$. The updating equation for $\mathbf{\Sigma}_e$ is

$$\mathbf{\Sigma}_e = \mathbf{L}^{-T}\left(\mathbf{L}^T \sum_{j=1}^{M_t}\left(\eta_j\left(\widetilde{\boldsymbol{\mu}}_{ej}\widetilde{\boldsymbol{\mu}}_{ej}^T + \widetilde{\mathbf{\Lambda}}_{ej}^{-1}\right) + (1-\eta_j)\widetilde{\mathbf{\Sigma}}_e\right)\mathbf{L}\right)^{1/2}\mathbf{L}^{-1}. \tag{S11}$$

#### 1.4 Adjustment of selection bias

We have accounted for selection bias due to $P$ value thresholding in the variational EM algorithm by evaluating conditional probabilities given $|\hat{\gamma}_{1,j}/\hat{s}_{X_1,j}| \geq t$. However, LD clumping can introduce additional bias, as it preferentially retains the most significant SNPs within each genomic region. Consequently, the $Z$-score boundary $t$ used in the conditional likelihood should be moderately increased to account for this additional selection effect. Moreover, when LD clumping is performed based on the target population's $P$ values, the corresponding threshold adjustment in the auxiliary population should reflect dataset-specific factors, including the sample size ratio and the genetic correlation between the two populations. More specifically, we modify the threshold as follows:

$$P \text{ value threshold} \leftarrow 2 \times \left[1 - \Phi\left(\left|\Phi^{-1}\left(1 - \frac{\text{IV threshold}}{2}\right)\right| + C\sqrt{\frac{N_1}{N_2}}\right)\right], \tag{S12}$$

where $\Phi(\cdot)$ is the CDF of the standard normal distribution, and $C$ represents a constant, while $N_1$ and $N_2$ correspond to the sample sizes of the auxiliary large-sample population and target population, respectively. We set $C = 0.13$ based on empirical calibration. We distinguish two thresholds: (i) the IV selection threshold in the right-hand side of Eq. (S12), the $P$ value cutoff used to select SNPs as instruments, and (ii) the conditioning threshold in the left-hand side of Eq. (S12), the $P$ value corresponding to the $Z$-score boundary $t$ at which the conditional likelihood is evaluated to account for selection bias. Equivalently,

$$Z\text{-score}\,(P \text{ value threshold}) \leftarrow Z\text{-score}\,(\text{IV threshold}) + C\sqrt{\frac{N_1}{N_2}}. \tag{S13}$$

To understand how a shift in the auxiliary population's selection threshold propagates to the target population's $Z$-Scores—and thereby to justify the $\sqrt{N_1/N_2}$ factor—we provide a simplified analysis of the conditional expectation $\mathbb{E}[z_{2j} \mid z_{1j} \geq t]$ and its sensitivity to the threshold $t$. This analysis reveals the rate at which tightening the auxiliary threshold induces additional selection on the target population, which in turn motivates the threshold adjustment.

For

$$z_{1j} = \frac{\hat{\gamma}_{1,j}}{\hat{s}_{X_1,j}},\ z_{2j} = \frac{\hat{\gamma}_{2,j}}{\hat{s}_{X_2,j}},$$

assume

$$\begin{pmatrix} \hat{\gamma}_{1,j} \\ \hat{\gamma}_{2,j} \end{pmatrix} \sim \mathcal{N}(\mathbf{0}, \mathbf{\Sigma}), \text{with } \mathbf{\Sigma} = \begin{pmatrix} \sigma_1^2 & \rho\sigma_1\sigma_2 \\ \rho\sigma_1\sigma_2 & \sigma_2^2 \end{pmatrix},$$

where $z_{1j}$ and $z_{2j}$ denote the $Z$-scores of exposure from the auxiliary population and target population, respectively, and $\hat{\gamma}_{1,j}$ and $\hat{\gamma}_{2,j}$ represent the observed effect sizes. $\hat{s}_{X_1,j}$ and $\hat{s}_{X_2,j}$ are the corresponding standard errors, with $\hat{s}_{X_1,j} \approx \frac{1}{\sqrt{N_{1j}}}, \hat{s}_{X_2,j} \approx \frac{1}{\sqrt{N_{2j}}}$, where $N_{1j}$ and $N_{2j}$ are sample sizes for SNP $j$ in two populations. For simplicity, we assume constant sample sizes for all SNPs: $N_{1j} = N_1, N_{2j} = N_2$. Then

$$\begin{pmatrix} z_{1j} \\ z_{2j} \end{pmatrix} \sim \mathcal{N}(\mathbf{0}, \mathbf{\Sigma'}), \text{with } \mathbf{\Sigma'} = \begin{pmatrix} N_1\sigma_1^2 & \sqrt{N_1N_2}\cdot\rho\sigma_1\sigma_2 \\ \sqrt{N_1N_2}\cdot\rho\sigma_1\sigma_2 & N_2\sigma_2^2 \end{pmatrix} := \begin{pmatrix} {\sigma'_1}^2 & \rho\sigma'_1\sigma'_2 \\ \rho\sigma'_1\sigma'_2 & {\sigma'_2}^2 \end{pmatrix}. \tag{S14}$$

We aim to show that $\frac{d\mathbb{E}[z_{2j}|z_{1j}\geq t]}{dt} \sim O\left(\rho\sqrt{\frac{N_2}{N_1}}\right)$ as $t \to \infty$. Note that in practice, IV selection uses $|z_{1j}| \geq t$, a two-sided condition. However, since the joint distribution of $(z_{1j}, z_{2j})$ is symmetric about the origin, it suffices to analyze the one-sided case $z_{1j} \geq t$ (restricting to positive values) and the conclusion extends directly to the two-sided setting by symmetry.

For $t > 0$, define $f(t) := \mathbb{E}\left[z_{2j} \mid z_{1j} \geq t\right]$. From Eq. (S14), $z_{2j}$ can be decomposed into: $z_{2j} = \rho\frac{\sigma'_2}{\sigma'_1}z_{1j} + \epsilon$, where $\epsilon \sim N\left(0, (1-\rho^2)\,{\sigma'_2}^2\right)$ and is independent of $z_{1j}$. Therefore,

$$\begin{aligned} f(t) = \mathbb{E}\left[\rho\frac{\sigma'_2}{\sigma'_1}z_{1j} + \epsilon\middle| z_{1j} \geq t\right] &= \rho\frac{\sigma'_2}{\sigma'_1}\,\mathbb{E}\left[z_{1j} \mid z_{1j} \geq t\right] + \mathbb{E}\left[\epsilon \mid z_{1j} \geq t\right] \\ \overset{\text{by independence}}{=\!=\!=\!=} \rho\frac{\sigma'_2}{\sigma'_1}\,\mathbb{E}\left[z_{1j} \mid z_{1j} \geq t\right] + \mathbb{E}\left[\epsilon\right] &= \rho\frac{\sigma'_2}{\sigma'_1}\,\mathbb{E}\left[z_{1j} \mid z_{1j} \geq t\right], \end{aligned}$$

where

$$\mathbb{E}\left[z_{1j} \mid z_{1j} \geq t\right] = \sigma_1' \, \mathbb{E}\left[\frac{z_{1j}}{\sigma_1'} \mid \frac{z_{1j}}{\sigma_1'} \geq \frac{t}{\sigma_1'}\right] \xlongequal{X:=\frac{z_{1j}}{\sigma_1'}\sim N(0,1)} \sigma_1' \, \mathbb{E}\left[X \mid X \geq \frac{t}{\sigma_1'}\right] = \sigma_1' \frac{\mathbb{E}\left[X 1_{\{X \geq \frac{t}{\sigma_1'}\}}\right]}{p(X \geq \frac{t}{\sigma_1'})}$$

$$= \sigma_1' \frac{\int_{\frac{t}{\sigma_1'}}^{\infty} x \cdot \frac{1}{\sqrt{2\pi}} \exp\left(-\frac{x^2}{2}\right) dx}{1 - \Phi(\frac{t}{\sigma_1'})} = \sigma_1' \frac{-\frac{1}{\sqrt{2\pi}} \exp\left(-\frac{x^2}{2}\right)\Big|_{x=\frac{t}{\sigma_1'}}^{\infty}}{1 - \Phi(\frac{t}{\sigma_1'})} = \sigma_1' \frac{\phi(\frac{t}{\sigma_1'})}{1 - \Phi(\frac{t}{\sigma_1'})},$$

with $\phi(\cdot)$ and $\Phi(\cdot)$ being the PDF and CDF of $N(0,1)$, respectively.

Define $\lambda(u) := \frac{\phi(u)}{1-\Phi(u)}$, which is known as the Inverse Mills Ratio (IMR). Then

$$\lambda'(u) = \frac{\phi'(u)\left(1 - \Phi(u)\right) + \phi^2(u)}{\left(1 - \Phi(u)\right)^2} = \frac{-u\phi(u)\left(1 - \Phi(u)\right) + \phi^2(u)}{\left(1 - \Phi(u)\right)^2}$$

$$= -u \frac{\phi(u)}{1 - \Phi(u)} + \left(\frac{\phi(u)}{1 - \Phi(u)}\right)^2 = -u\lambda(u) + \lambda^2(u) = \lambda(u)\left(\lambda(u) - u\right).$$

Thereby,

$$\frac{df(t)}{dt} = \frac{d\left(\rho \frac{\sigma_2'}{\sigma_1'} \cdot \sigma_1' \lambda(\frac{t}{\sigma_1'})\right)}{dt} = \rho\sigma_2' \frac{d\lambda(\frac{t}{\sigma_1'})}{dt} = \rho \frac{\sigma_2'}{\sigma_1'} \lambda'(\frac{t}{\sigma_1'}) = \rho \frac{\sigma_2'}{\sigma_1'} \lambda(\frac{t}{\sigma_1'}) \left(\lambda(\frac{t}{\sigma_1'}) - \frac{t}{\sigma_1'}\right). \tag{S15}$$

When $u$ tends to infinity, the asymptotic expansion of Mills ratio $\frac{1}{\lambda(u)}$ is (see [3])

$$\frac{1}{\lambda(u)} \sim \frac{1}{u} - \frac{1}{u^3} + \frac{1 \cdot 3}{u^5} + \cdots, u \to \infty.$$

Then

$$\lambda(u) \sim \frac{1}{\frac{1}{u} - \frac{1}{u^3} + \frac{1\cdot 3}{u^5} + o(\frac{1}{u^5})} = u + \frac{1}{u} - \frac{2}{u^3} + o(\frac{1}{u^3}), u \to \infty,$$

which leads to

$$\lambda(u)\left(\lambda(u) - u\right) \sim \left(u + \frac{1}{u} + O(\frac{1}{u^3})\right)\left(\frac{1}{u} + O(\frac{1}{u^3})\right) = 1 + O(\frac{1}{u^2}) \to 1, \text{ when } u \to \infty.$$

Substituting these results into Eq. (S15), we can derive

$$\frac{d\,\mathbb{E}\left[z_{2j} \mid z_{1j} \geq t\right]}{dt} = \frac{df(t)}{dt} \to \rho \frac{\sigma_2'}{\sigma_1'} = \rho \frac{\sigma_2}{\sigma_1} \sqrt{\frac{N_2}{N_1}}, \text{ when } t \to \infty. \tag{S16}$$

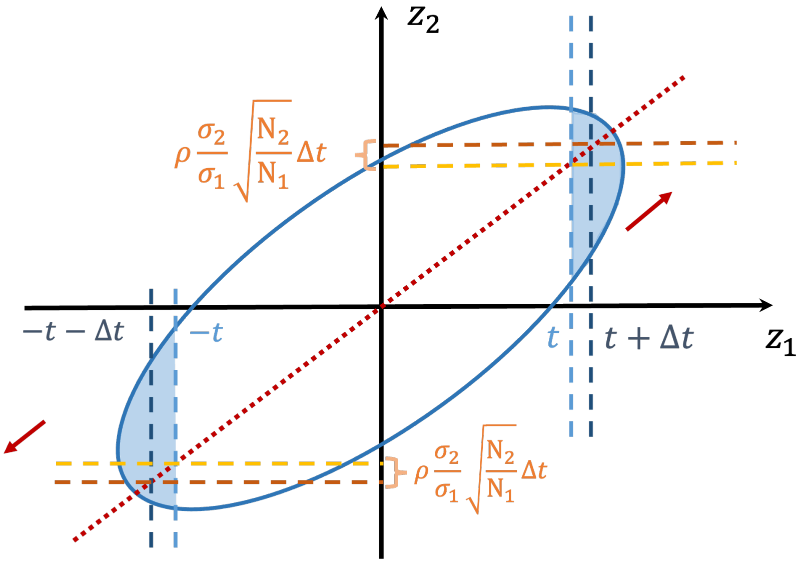

Figure S1: **Illustration of $Z$-score shift ratio.** The $Z$-scores of the two populations follow a joint Gaussian distribution, where the probability density function is represented by the blue ellipse. When the threshold for the auxiliary population $z_1$ increases from $t$ to $t + \Delta t$, the corresponding magnitude for the target population $z_2$ increases by $\rho \frac{\sigma_2}{\sigma_1} \sqrt{\frac{N_2}{N_1}}$. The factor is determined by the shape of the PDF contour, which depends on the covariance matrix and thus, genetic correlation and sample size ratio between two populations.

The result above shows that increasing the auxiliary population's selection threshold by $\Delta t$ shifts the expected target population $Z$-score by approximately $\rho \frac{\sigma_2}{\sigma_1} \sqrt{\frac{N_2}{N_1}} \cdot \Delta t$. However, since LD clumping is performed using the target population's $P$ values, the relevant question is the inverse: a selection effect of magnitude $\delta$ in the target population's $Z$-score scale corresponds to an auxiliary-population threshold shift of $\delta \cdot \frac{\sigma_1}{\rho \sigma_2} \sqrt{\frac{N_1}{N_2}}$. In other words, to equivalently capture the additional selection bias introduced by LD clumping in the target population, the conditioning threshold in the auxiliary population should be shifted by a factor proportional to $\sqrt{N_1/N_2}$. Since the quantities $\rho$, $\sigma_1$, $\sigma_2$, and the magnitude of the shift are not directly observable, we absorb their combined effect into a single empirical constant $C$. The constant $C = 0.13$ was calibrated by selecting the value that yields well-controlled type I error rates across a range of experiments. This leads to the threshold adjustment $C\sqrt{\frac{N_1}{N_2}}$ in Eqs. (S12) and (S13). Fig. S1 provides an intuitive illustration of this relationship.

### 2 Supplementary notes

#### 2.1 Simulation design

Our simulation is designed to evaluate XMR under realistic and challenging conditions, including model misspecification (only a subset of SNPs exhibit correlated pleiotropy), population-specific IV validity (a SNP may be valid in only one population), and varying levels of genetic correlation between populations. We simulate genome-wide summary statistics for $m$ = 50,000 SNPs across an auxiliary population ($N_1$ = 80,000) and a target population ($N_2$ = 15,000), then apply IV selection to obtain candidate instruments. Below, we describe the data-generating process in detail.

We simulated effects $(\hat{\gamma}_{1,j}, \hat{\gamma}_{2,j}, \hat{\Gamma}_{2,j})$ of SNP $j$ based on the following relationship:

$$\begin{pmatrix} \hat{\gamma}_{1,j} \\ \hat{\gamma}_{2,j} \\ \hat{\Gamma}_{2,j} \end{pmatrix} = \begin{pmatrix} Z_{1,j}\gamma_{1,j} \\ Z_{2,j}\gamma_{2,j} \\ \beta Z_{2,j}\gamma_{2,j} + \alpha_j \end{pmatrix} + I_j \cdot \begin{pmatrix} u_{1,j} \\ u_{2,j} \\ v_{2,j} \end{pmatrix} + \begin{pmatrix} \epsilon_{1,j} \\ \epsilon_{2,j} \\ \xi_{2,j} \end{pmatrix}, \tag{S17}$$

where $\beta$ is the target causal effect, $(\gamma_{1,j}, \gamma_{2,j}, \alpha_j)$ represent true effects of SNP $j$ on exposure $X_1$ and $X_2$ and its direct effect on outcome $Y_2$, $(u_{1,j}, u_{2,j}, v_{2,j})$ denote correlated pleiotropy, and $(\epsilon_{1,j}, \epsilon_{2,j}, \xi_{2,j})$ are sample structure terms, following definitions in the main text. With $m$ = 50,000 SNPs, the standard errors are set as $\hat{s}_{X_1,j} = \frac{1}{\sqrt{N_1}} = \frac{1}{\sqrt{80000}}$, $\hat{s}_{X_2,j} = \frac{1}{\sqrt{N_2}} = \frac{1}{\sqrt{15000}}$ and $\hat{s}_{Y_2,j} = \frac{1}{\sqrt{N_2}} = \frac{1}{\sqrt{15000}}$, reflecting the different precision levels of the two populations.

The sample structure terms are modeled as:

$$\begin{pmatrix} \epsilon_{1,j} \\ \epsilon_{2,j} \\ \xi_{2,j} \end{pmatrix} \sim \mathcal{N}\left(\mathbf{0}, \hat{\boldsymbol{S}}_j \boldsymbol{C} \hat{\boldsymbol{S}}_j\right), \quad \text{with } \hat{\boldsymbol{S}}_j = \begin{pmatrix} \hat{s}_{X_1,j} & 0 & 0 \\ 0 & \hat{s}_{X_2,j} & 0 \\ 0 & 0 & \hat{s}_{Y_2,j} \end{pmatrix}, \boldsymbol{C} = \begin{pmatrix} c_1 & c_{12} & c_{1y} \\ c_{12} & c_2 & c_{2y} \\ c_{1y} & c_{2y} & c_y \end{pmatrix}, \tag{S18}$$

where $c_1 = 1.25$, $c_2 = 1.05$, and $c_y = 1.02$, consistent with greater population stratification expected in larger cohorts. Assuming greater sample overlap within the same population than across populations, we set $c_{2y} = 0.15$, $c_{12} = 0.03$, and $c_{1y} = 0.015$.

The pleiotropic effects $(u_{1,j}, u_{2,j}, v_{2,j})$ are sampled from:

$$\begin{pmatrix} u_{1,j} \\ u_{2,j} \\ v_{2,j} \end{pmatrix} \sim \mathcal{N}(\mathbf{0}, \boldsymbol{\Omega}_j), \quad \text{where } \boldsymbol{\Omega}_j = \begin{pmatrix} \omega_1^2 & \omega_{12} & \omega_{1y} \\ \omega_{12} & \omega_2^2 & \omega_{2y} \\ \omega_{1y} & \omega_{2y} & \omega_y^2 \end{pmatrix}. \tag{S19}$$

Here we set all LD scores to 1 for simplicity. The binary indicator $I_j$ controls whether correlated pleiotropy is present for SNP $j$: we set $I_j = 1$ for $m_1$ = 30,000 out of the 50,000

SNPs and $I_j = 0$ for the remaining 20,000. This partial-pleiotropy design introduces model misspecification relative to XMR's assumption that all SNPs share the same pleiotropic covariance structure, thereby testing its robustness. Assuming a total heritability $h^2 = 0.5$, we set $\omega_1^2 = \omega_2^2 = \omega_y^2 = \frac{h^2}{m_1} = \frac{0.5}{30000}$, and $(\omega_{12}, \omega_{1y}, \omega_{2y}) = \frac{h^2}{m_1} \times (0.5, 0.2, 0.3)$, implying a stronger genetic correlation between the same trait across populations than between different traits. Since we assume all LD scores are equal to 1, the actual $\boldsymbol{C}$ and $\boldsymbol{\Omega}_j$ are directly used as inputs for XMR.

The effects $(\gamma_{1,j}, \gamma_{2,j}, \alpha_j)$ are modeled as:

$$\begin{pmatrix} \gamma_{1,j} \\ \gamma_{2,j} \\ \alpha_j \end{pmatrix} \sim \mathcal{N}(\mathbf{0}, \mathbf{\Sigma}), \quad \text{with } \mathbf{\Sigma} = \begin{pmatrix} \sigma_{1,f}^2 & \rho\sigma_{1,f}\sigma_{2,f} & 0 \\ \rho\sigma_{1,f}\sigma_{2,f} & \sigma_{2,f}^2 & 0 \\ 0 & 0 & \tau^2 \end{pmatrix}, \tag{S20}$$

where $(\sigma_{1,f}^2, \sigma_{2,f}^2, \tau^2) = \frac{h^2}{m_1} \times (10, 10, 1)$. The ratios $\sigma_{1,f}^2 : \omega_1^2 = \sigma_{2,f}^2 : \omega_2^2 = 10$ ensure strong instrument strength, while $\tau^2 : \omega_y^2 = 1$ ensures that the magnitude of the direct effect $\alpha_j$ is comparable to that of the polygenic effects $v_{2,j}$. The genetic correlation between populations varies among $\rho \in \{0, 0.3, 0.7\}$, providing a comprehensive evaluation across scenarios ranging from no cross-population correlation to strong correlation.

The binary variables $Z_{1,j}$ and $Z_{2,j}$ control whether SNP $j$ has a true causal effect on the exposure in the auxiliary and target populations, respectively. For the 20,000 SNPs without correlated pleiotropy ($I_j = 0$), we set $Z_{1,j} = Z_{2,j} = 0$, meaning these SNPs have no true exposure effects and serve as null instruments. Among the 30,000 SNPs with correlated pleiotropic effects ($I_j = 1$), we randomly select 5% (1,500 SNPs) to be valid instruments in at least one population. Unlike XMR's assumption of consistent IV validity across populations, our simulation allows a SNP to be valid in only one of the two populations, introducing a form of model misspecification. Specifically, of the 1,500 valid SNPs, 250 have $(Z_{1,j} = 0, Z_{2,j} = 1)$, 150 have $(Z_{1,j} = 1, Z_{2,j} = 0)$, and the remaining 1,100 have $Z_{1,j} = Z_{2,j} = 1$. The remaining 28,500 SNPs with $I_j = 1$ have $Z_{1,j} = Z_{2,j} = 0$ and contribute only through their pleiotropic effects.

The causal effect $\beta$ takes values in $\{0, 0.05, 0.1, 0.15, 0.2, 0.25, 0.3\}$, allowing us to evaluate the type I error rate under the null ($\beta = 0$) and statistical power under the alternatives ($\beta \neq 0$). After generating $(\hat{\gamma}_{1,j}, \hat{\gamma}_{2,j}, \hat{\Gamma}_{2,j})$ from the above model, we compute $Z$-scores as $(\frac{\hat{\gamma}_{1,j}}{\hat{s}_{X_1,j}}, \frac{\hat{\gamma}_{2,j}}{\hat{s}_{X_2,j}}, \frac{\hat{\Gamma}_{2,j}}{\hat{s}_{Y_2,j}})$ and derive the corresponding $P$ values. Summary statistics from all three samples are used as inputs for XMR, while two-sample MR methods use only those from the target population.

#### 2.2 Simulation design for TEMR

TEMR requires auxiliary-population summary statistics for the outcome trait in addition to the exposure, i.e., it uses four sets of summary statistics rather than three. To accommodate this, we extend the three-variable model in section 2.1 by appending a fourth component corresponding to the auxiliary outcome $Y_1$. All parameters from section 2.1 are retained unchanged; below we describe only the new elements introduced for the auxiliary outcome.

The observed effects $(\hat{\gamma}_{1,j}, \hat{\gamma}_{2,j}, \hat{\Gamma}_{2,j}, \hat{\Gamma}_{1,j})$ of SNP $j$ are generated as:

$$\begin{pmatrix} \hat{\gamma}_{1,j} \\ \hat{\gamma}_{2,j} \\ \hat{\Gamma}_{2,j} \\ \hat{\Gamma}_{1,j} \end{pmatrix} = \begin{pmatrix} Z_{1,j}\gamma_{1,j} \\ Z_{2,j}\gamma_{2,j} \\ \beta Z_{2,j}\gamma_{2,j} + \alpha_j \\ \beta_1 Z_{1,j}\gamma_{1,j} + \alpha_{1,j} \end{pmatrix} + I_j \cdot \begin{pmatrix} u_{1,j} \\ u_{2,j} \\ v_{2,j} \\ v_{1,j} \end{pmatrix} + \begin{pmatrix} \epsilon_{1,j} \\ \epsilon_{2,j} \\ \xi_{2,j} \\ \xi_{1,j} \end{pmatrix}, \tag{S21}$$

where $\hat{\Gamma}_{1,j}$ is the observed effect of SNP $j$ on the auxiliary-population outcome $Y_1$, and $\beta_1$ is the causal effect of $X_1$ on $Y_1$ in the auxiliary population. The term $\alpha_{1,j}$ represents the direct effect of SNP $j$ on $Y_1$ that is not mediated through the exposure, while $v_{1,j}$ and $\xi_{1,j}$ denote the corresponding correlated pleiotropy and sample structure terms for $Y_1$, respectively. The first three components are identical to Eq. (S17).

The sample structure terms are extended to four dimensions:

$$\begin{pmatrix} \epsilon_{1,j} \\ \epsilon_{2,j} \\ \xi_{2,j} \\ \xi_{1,j} \end{pmatrix} \sim \mathcal{N}\left(\mathbf{0}, \hat{\boldsymbol{S}}_j \boldsymbol{C} \hat{\boldsymbol{S}}_j\right), \quad \text{with } \hat{\boldsymbol{S}}_j = \begin{pmatrix} \hat{s}_{X_1,j} & 0 & 0 & 0 \\ 0 & \hat{s}_{X_2,j} & 0 & 0 \\ 0 & 0 & \hat{s}_{Y_2,j} & 0 \\ 0 & 0 & 0 & \hat{s}_{Y_1,j} \end{pmatrix}, \boldsymbol{C} = \begin{pmatrix} c_1 & c_{12} & c_{1y} & c_{41} \\ c_{12} & c_2 & c_{2y} & c_{42} \\ c_{1y} & c_{2y} & c_y & c_{4y} \\ c_{41} & c_{42} & c_{4y} & c_4 \end{pmatrix}, \tag{S22}$$

where the upper-left $3 \times 3$ block of $\boldsymbol{C}$ is identical to Eq. (S18). The standard error for the auxiliary outcome is $\hat{s}_{Y_1,j} = \frac{1}{\sqrt{N_1}} = \frac{1}{\sqrt{80000}}$, matching the auxiliary exposure precision. For the new entries, we set $c_4 = 1.1$, $c_{41} = 0.1$, $c_{42} = 0.01$, and $c_{4y} = 0.008$, where the subscript 4 refers to the fourth component ($Y_1$). The value $c_{41} = 0.1$ reflects substantial sample overlap between $X_1$ and $Y_1$ within the auxiliary population, whereas $c_{42}$ and $c_{4y}$ are much smaller, reflecting minimal overlap between the auxiliary outcome and the target-population samples.

The pleiotropic effects are extended analogously:

$$\begin{pmatrix} u_{1,j} \\ u_{2,j} \\ v_{2,j} \\ v_{1,j} \end{pmatrix} \sim \mathcal{N}\left(\mathbf{0}, \boldsymbol{\Omega}_j\right), \quad \text{where } \boldsymbol{\Omega}_j = \begin{pmatrix} \omega_1^2 & \omega_{12} & \omega_{1y} & \omega_{41} \\ \omega_{12} & \omega_2^2 & \omega_{2y} & \omega_{42} \\ \omega_{1y} & \omega_{2y} & \omega_y^2 & \omega_{4y} \\ \omega_{41} & \omega_{42} & \omega_{4y} & \omega_4^2 \end{pmatrix}, \tag{S23}$$

with $(\omega_{41}, \omega_{42}, \omega_{4y}, \omega_4^2) = \frac{h^2}{m_1} \times (0.3, 0.1, 0.5, 1)$. The upper-left $3 \times 3$ block is identical to Eq. (S19). The choice $\omega_4^2 = \frac{h^2}{m_1}$ gives the auxiliary outcome the same per-SNP pleiotropic variance as the other components. The relatively large value of $\omega_{4y} = 0.5 \cdot \frac{h^2}{m_1}$ reflects strong pleiotropic correlation between the same outcome trait measured in the two populations, while $\omega_{42} = 0.1 \cdot \frac{h^2}{m_1}$ implies weaker pleiotropic correlation between the auxiliary outcome and the target exposure.

The effects $(\gamma_{1,j}, \gamma_{2,j}, \alpha_j, \alpha_{1,j})$ are modeled as:

$$\begin{pmatrix} \gamma_{1,j} \\ \gamma_{2,j} \\ \alpha_j \\ \alpha_{1,j} \end{pmatrix} \sim \mathcal{N}(\mathbf{0}, \mathbf{\Sigma}), \quad \text{with } \mathbf{\Sigma} = \begin{pmatrix} \sigma_{1,f}^2 & \rho\sigma_{1,f}\sigma_{2,f} & 0 & 0 \\ \rho\sigma_{1,f}\sigma_{2,f} & \sigma_{2,f}^2 & 0 & 0 \\ 0 & 0 & \tau^2 & 0 \\ 0 & 0 & 0 & \tau_1^2 \end{pmatrix}, \tag{S24}$$

where $\tau_1^2 = \frac{h^2}{m_1}$, so that $\tau_1^2 : \omega_4^2 = 1$, mirroring the ratio $\tau^2 : \omega_y^2 = 1$ used for the target population. The block-diagonal structure of $\mathbf{\Sigma}$ encodes the InSIDE condition for both populations: the direct effects $\alpha_j$ and $\alpha_{1,j}$ are assumed independent of the exposure effects $(\gamma_{1,j}, \gamma_{2,j})$. The IV validity indicators $Z_{1,j}$ and $Z_{2,j}$ follow the same configuration as in section 2.1.

We set $\beta_1 = \beta/2$, so that a causal effect is present in the auxiliary population whenever it is present in the target population (and absent in both under the null). This constitutes a favorable experimental setting for TEMR, since it ensures that the auxiliary outcome data are informative about the causal relationship of interest. In practice, the auxiliary-population causal effect may differ substantially from the target-population effect or may even be absent; such scenarios would be less advantageous for TEMR.

After generating $(\hat{\gamma}_{1,j}, \hat{\gamma}_{2,j}, \hat{\Gamma}_{2,j}, \hat{\Gamma}_{1,j})$ from the model above, we compute the corresponding $Z$-scores and $P$ values as described in section 2.1. All four sets of summary statistics are used as inputs for TEMR.

#### 2.3 Real data analysis

##### 2.3.1 GWAS summary datasets formatting and pre-processing.

###### 2.3.1.1 Step 1: quality control

Prior to IV selection and MR estimation, we applied a series of standard quality control (QC) filters to each GWAS summary-statistics dataset to ensure that only high-quality, unambiguous SNPs were retained.

First, we excluded any SNP lacking essential information, namely the rs identifier, effect and non-effect alleles, estimated effect size, standard error, sample size, and association $P$ value. Second, SNPs appearing more than once (i.e., sharing the same rs identifier) were discarded to prevent ambiguity. Third, we excluded all SNPs located within the Major Histocompatibility Complex region on chromosome 6 (26–34 Mb) owing to its highly complex LD structure. Fourth, we restricted the analysis to unambiguous SNPs whose allele pairs are A/G, A/C, T/G, or T/C, thereby excluding variants that cannot be reliably aligned across datasets. Fifth, SNPs with a minor allele frequency (MAF) below a certain threshold (e.g., 0.01 in our analysis) were filtered out, since rare variants are more susceptible to genotyping and imputation errors. Sixth, when imputation quality scores were available, we removed SNPs with an information score below 0.9, as these are indicative of poor imputation accuracy. Finally, to mitigate the influence of outlying association signals, we removed SNPs whose $\chi^2$ statistic exceeded $\max\{80,\ N/1000\}$, where $N$ denotes the GWAS sample size.

In addition, following a strategy similar to that employed in LDSC [1], we further confined the SNP set to those present in the HapMap 3 reference panel. This panel provides a curated collection of common, well-imputed variants and serves as a safeguard when MAF or imputation quality information is unavailable in certain datasets.

After applying these filters, each GWAS dataset was reformatted to retain only the rs identifier, effect allele, non-effect allele, effect size, standard error, and $P$ value for the SNPs that passed all QC criteria. Throughout our analyses, we assumed that both the phenotype and genotypes in each GWAS had been standardized to zero mean and unit variance, so that effect sizes and standard errors could be equivalently derived from $Z$-scores and sample sizes when necessary.

We note that this genome-wide QC procedure is also a prerequisite for methods such as XMR, MRAPSS, and CAUSE, which leverage genome-wide summary statistics to estimate background parameters (e.g., the sample structure matrix $\boldsymbol{C}$ and the pleiotropic covariance $\boldsymbol{\Omega}$) in their respective models.

###### 2.3.1.2 Step 2: SNP effect alignment

A valid MR analysis requires that the SNP-exposure and SNP-outcome effect estimates refer to the same allele. To this end, we carried out a two-stage harmonization procedure for each exposure–outcome pair. In the first stage, we compared the allele coding between the exposure and outcome GWAS and, where necessary, modified the alleles in one dataset to match the strand orientation of the other dataset. In the second stage, we verified whether the designated effect allele was consistent across the two datasets; when it was not, we reversed the sign of the effect estimate in one dataset so that all effects were expressed with respect to the same reference allele.

As a concrete example, consider a SNP recorded as A/G (effect/non-effect) in the exposure GWAS and C/T (effect/non-effect) in the outcome GWAS. Strand-flipping the outcome alleles yields G/A, revealing that the outcome effect allele (G) corresponds to the non-effect allele of the exposure; accordingly, the sign of the outcome effect estimate would be reversed. Because this alignment procedure is only reliable for non-palindromic SNPs with allele pairs A/G, A/C, T/G, or T/C, any SNP with an ambiguous (palindromic) allele configuration was excluded at this stage. This exclusion is consistent with the allele-type filter already applied during quality control (Step 1), ensuring that no ambiguous SNPs enter the analysis. For XMR and TEMR, which require three or four GWAS datasets simultaneously, we generalized this step by designating one dataset as the anchor and harmonizing all remaining datasets to it.

###### 2.3.1.3 Step 3: IV selection and LD clumping

Using the harmonized summary datasets, we proceeded to select instrumental variables for each exposure–outcome pair. SNPs were deemed candidate instruments if their association with the exposure reached a pre-specified $P$ value threshold; the specific threshold adopted for each method is described in section 2.3.2. We then applied LD clumping via PLINK with an $r^2$ threshold of 0.001 within a 1 Mb window to obtain a set of approximately independent SNPs. Samples from the target population in the 1000 Genomes Project served as the LD reference panel for this clumping step.

For a reliable analysis, we imposed two minimum requirements on the number of IVs. First, each trait pair was required to have at least 4 independent IVs after clumping; pairs failing to meet this criterion were excluded from downstream analysis. Second, as a data-quality check, we verified the number of SNPs that reached genome-wide significance ($P \leq 5 \times 10^{-8}$) after LD clumping. If fewer than 15 genome-wide significant SNPs remained, the trait pair was deemed to have insufficient instrument strength and was excluded from further analysis.

##### 2.3.2 Parameter settings for XMR and compared methods.

Different MR methods have distinct requirements for IV selection thresholds and parameter estimation. Below we describe the settings used for each method in the real-data analyses.

For XMR, we first estimated $\boldsymbol{\Omega}$ and $\boldsymbol{C}$ using bivariate LDSC on genome-wide summary statistics. Trait pairs yielding invalid estimates (e.g., negative diagonal elements in $\boldsymbol{\Omega}$ or non-positive definite $\boldsymbol{C}$) were excluded from further analysis. To ensure numerical stability, if the off-diagonal elements of $\boldsymbol{\Omega}$ implied correlations outside the range $[-1, 1]$, they were truncated to $\pm 0.95$. IVs were selected based on a significance threshold of $P \leq 5 \times 10^{-5}$ in the exposure GWAS of the target population. These SNPs were further pruned for independence using PLINK clumping ($r^2$ threshold of 0.001, 1Mb window) before applying the XMR model.

For MRAPSS, the confounding factor estimates were derived from the corresponding elements of $\boldsymbol{\Omega}$ and $\boldsymbol{C}$ computed as described above. IV selection followed the method's default significance threshold ($P \leq 5 \times 10^{-5}$).

For TEMR, we adhered to its standard protocol for combining $P$ values, selecting IVs based on a combined $P$ value threshold of $5 \times 10^{-8}$.

For CAUSE, we observed that the recommended default threshold ($P \leq 1 \times 10^{-3}$) produced unstable estimates in scenarios with limited sample sizes. Consequently, we adopted a stricter threshold of $P \leq 5 \times 10^{-5}$, which yielded comparable but more robust performance. A detailed comparison of the two thresholds is provided in section 4.9.

For the remaining two-sample MR methods, we employed the genome-wide significance threshold ($P \leq 5 \times 10^{-8}$) for IV selection in datasets with sufficient sample sizes ($N \geq$ 20,000). When sample sizes were smaller, a relaxed threshold ($P \leq 5 \times 10^{-5}$) was applied to retain a sufficient number of IVs for causal inference.

##### 2.3.3 Hypothesis test for the difference of causal effect estimates.

To investigate potential heterogeneity in causal effects across ancestry groups (East Asians (EAS), Central/South Asians (CSA), and Africans (AFR)), we compared the causal effect estimate in each target population against the corresponding estimate in the European (EUR) population for every trait pair. Since XMR is designed for cross-population analysis and cannot be applied to a single-population dataset, we used MRAPSS—which shares a similar statistical framework—to obtain the reference causal effect estimate for the European population.

Specifically, for each trait pair we tested:

$$H_0 : \beta_{\text{tar}} - \beta_{\text{EUR}} = 0 \quad \text{vs.} \quad H_1 : \beta_{\text{tar}} - \beta_{\text{EUR}} \neq 0,$$

where $\beta_{\text{tar}}$ denotes the XMR estimate for the target population and $\beta_{\text{EUR}}$ denotes the MRAPSS estimate for the European population. Let $SE_{\text{tar}}$ and $SE_{\text{EUR}}$ denote the corresponding standard errors. Because the two estimates are derived from non-overlapping samples, they can be treated as independent, and the test statistic is:

$$Z = \frac{\beta_{\text{tar}} - \beta_{\text{EUR}}}{\sqrt{SE_{\text{tar}}^2 + SE_{\text{EUR}}^2}}.$$

Under the null hypothesis, $Z$ follows a standard normal distribution. We computed the two-sided $P$ value and declared significant heterogeneity when the $P$ value fell below 0.05.

##### 2.3.4 Meta-analysis for two EAS cohorts.

To obtain more robust causal effect estimates for the EAS population and to facilitate cross-ancestry comparisons, we performed a meta-analysis combining estimates from the two EAS cohorts (BBJ and TPMI). We adopted the framework proposed by Xiao et al. [4], which leverages the correlation structure between cohorts to improve estimation accuracy. We observed that raw estimates derived from BBJ were systematically larger than those from TPMI, likely attributable to environmental heterogeneity or measurement differences between the cohorts; the meta-analysis helps to mitigate such discrepancies. The resulting augmented estimates were used for all subsequent cross-ancestry comparisons.

Specifically, let $\hat{\beta}_{1j}$ and $\hat{\beta}_{2j}$ denote the MR estimates for trait pair $j$ in BBJ and TPMI, respectively, for $j = 1, \ldots, N$ (in our analysis, $N = 105$). We assume that each estimate can be decomposed into a latent true effect and independent estimation noise:

$$\begin{pmatrix} \hat{\beta}_{1j} \\ \hat{\beta}_{2j} \end{pmatrix} = \begin{pmatrix} \beta_{1j} \\ \beta_{2j} \end{pmatrix} + \begin{pmatrix} e_{1j} \\ e_{2j} \end{pmatrix}, \quad \text{where } \begin{pmatrix} \beta_{1j} \\ \beta_{2j} \end{pmatrix} := \boldsymbol{\beta}_j \sim \mathcal{N}(\mathbf{0}, \boldsymbol{B}), \quad \begin{pmatrix} e_{1j} \\ e_{2j} \end{pmatrix} \sim \mathcal{N}\left(\mathbf{0}, \hat{\boldsymbol{E}}_j\right), \quad \text{(S25)}$$

with covariance matrices defined as:

$$\boldsymbol{B} = \begin{pmatrix} b_{11} & b_{12} \\ b_{12} & b_{22} \end{pmatrix}, \quad \hat{\boldsymbol{E}}_j = \begin{pmatrix} \hat{s}_{1j}^2 & 0 \\ 0 & \hat{s}_{2j}^2 \end{pmatrix}. \quad \text{(S26)}$$

Here, $\hat{s}_{1j}^2$ and $\hat{s}_{2j}^2$ represent the squared standard errors of the estimates $\hat{\beta}_{1j}$ and $\hat{\beta}_{2j}$, respectively. The off-diagonal element $b_{12}$ captures the covariance between the true causal effects in the two cohorts, which is expected to be positive when both cohorts share the same underlying biology but may differ due to cohort-specific factors.

We estimate the covariance matrix of the true effects, $\boldsymbol{B}$, using the method of moments. Given that $\begin{pmatrix} \hat{\beta}_{1j} \\ \hat{\beta}_{2j} \end{pmatrix} := \hat{\boldsymbol{\beta}}_j \sim \mathcal{N}\left(\mathbf{0}, \boldsymbol{B} + \hat{\boldsymbol{E}}_j\right)$, we have:

$$
\begin{aligned}
\mathbb{E}\left[\hat{\boldsymbol{\beta}}_j \hat{\boldsymbol{\beta}}_j^T\right] = \boldsymbol{B} + \hat{\boldsymbol{E}}_j \Rightarrow \mathbb{E}\left[\hat{\boldsymbol{\beta}}_j \hat{\boldsymbol{\beta}}_j^T - \hat{\boldsymbol{E}}_j\right] = \boldsymbol{B} \\
\Rightarrow \mathbb{E}\left[\frac{1}{N}\sum_{j=1}^{N}\left(\hat{\boldsymbol{\beta}}_j \hat{\boldsymbol{\beta}}_j^T - \hat{\boldsymbol{E}}_j\right)\right] = \boldsymbol{B}.
\end{aligned}
\tag{S27}
$$

Consequently, $\boldsymbol{B}$ is estimated as $\hat{\boldsymbol{B}} = \frac{1}{N}\sum_{j=1}^{N}\left(\hat{\boldsymbol{\beta}}_j \hat{\boldsymbol{\beta}}_j^T - \hat{\boldsymbol{E}}_j\right)$.

Next, we apply the generalized method of moments (GMM) [4, 5, 6] to derive a meta estimator, denoted as $\{\hat{\boldsymbol{\beta}}_j^{meta}, \hat{\mathbf{s}}^{meta}\} = \{\hat{\beta}_{1j}^{meta}, \hat{\beta}_{2j}^{meta}, \hat{s}_{1j}^{meta}, \hat{s}_{2j}^{meta}\}_{j=1,\dots,N}$. The moment condition is established by linearly projecting the estimate from the auxiliary population (e.g., TPMI) onto the true marginal effect of the target population (e.g., BBJ):

$$
\mathbb{E}\Big[\hat{\beta}_{2j} - \beta_{1j}\gamma\Big] = 0, \tag{S28}
$$

where $\gamma$ is an unknown coefficient.

The GMM estimator for $\gamma$ is obtained by minimizing the following objective function, using an identity matrix as the weight matrix:

$$
\mathbb{E}\Big[(\hat{\beta}_{2j} - \beta_{1j}\gamma)^2\Big]. \tag{S29}
$$

Taking the derivative with respect to $\gamma$ and setting it to zero yields:

$$
\begin{aligned}
0 &= \frac{\partial}{\partial\gamma}\,\mathbb{E}\Big[(\hat{\beta}_{2j} - \beta_{1j}\gamma)^2\Big] \\
&= \mathbb{E}\left[\frac{\partial}{\partial\gamma}(\hat{\beta}_{2j} - \beta_{1j}\gamma)^2\right] \\
&= -2\,\mathbb{E}\Big[\beta_{1j}(\hat{\beta}_{2j} - \beta_{1j}\gamma)\Big] \\
&= -2\,\mathbb{E}[\beta_{1j}(\beta_{2j} + e_{2j} - \beta_{1j}\gamma)] \\
&= -2\,\mathbb{E}[\beta_{1j}\,\beta_{2j}] + 2\,\mathbb{E}\big[\beta_{1j}^2\,\gamma\big] \\
&= -2\,b_{12} + 2\,b_{11}\,\gamma.
\end{aligned}
\tag{S30}
$$

Solving this gives:

$$
\hat{\gamma} = \frac{b_{12}}{b_{11}}. \tag{S31}
$$

This implies that the projection of $\hat{\beta}_{2j}$ onto the true marginal effect $\beta_{1j}$ is $\frac{b_{12}}{b_{11}}\beta_{1j}$.

Extending this to the full vector of estimates, the moment condition becomes:

$$\mathbb{E}\left[\begin{pmatrix}\hat{\beta}_{1j}\\ \hat{\beta}_{2j}\end{pmatrix} - \frac{\boldsymbol{b}_{\cdot 1}}{b_{11}}\beta_{1j}\right] = 0, \quad \text{where } \boldsymbol{b}_{\cdot 1} = \begin{pmatrix}b_{11}\\ b_{12}\end{pmatrix}. \tag{S32}$$

Accordingly, the conditional mean is derived as:

$$\mathbb{E}\left[\begin{pmatrix}\hat{\beta}_{1j}\\ \hat{\beta}_{2j}\end{pmatrix} - \frac{\boldsymbol{b}_{\cdot 1}}{b_{11}}\beta_{1j}\middle|\beta_{1j}\right] = \mathbb{E}\left[\begin{pmatrix}\hat{\beta}_{1j}\\ \hat{\beta}_{2j}\end{pmatrix}\middle|\beta_{1j}\right] - \mathbb{E}\left[\frac{\boldsymbol{b}_{\cdot 1}}{b_{11}}\beta_{1j}\middle|\beta_{1j}\right] = 0,$$

$$\Rightarrow \mathbb{E}\left[\begin{pmatrix}\hat{\beta}_{1j}\\ \hat{\beta}_{2j}\end{pmatrix}\middle|\beta_{1j}\right] = \frac{\boldsymbol{b}_{\cdot 1}}{b_{11}}\beta_{1j}. \tag{S33}$$

Next, the conditional variance is given by:

$$\begin{aligned}\text{Var}\left[\begin{pmatrix}\hat{\beta}_{1j}\\ \hat{\beta}_{2j}\end{pmatrix}\middle|\beta_{1j}\right] &= \text{Var}\left[\begin{pmatrix}\beta_{1j}\\ \beta_{2j}\end{pmatrix}\middle|\beta_{1j}\right] + \text{Var}\left[\begin{pmatrix}e_{1j}\\ e_{2j}\end{pmatrix}\middle|\beta_{1j}\right]\\ &= \boldsymbol{B} - \frac{\boldsymbol{b}_{\cdot 1}\boldsymbol{b}_{\cdot 1}^T}{b_{11}} + \hat{\mathbf{E}}_j := \boldsymbol{\Lambda}_{1j}^{-1}.\end{aligned} \tag{S34}$$

The GMM estimator for $\beta_{1j}$ is then obtained by minimizing the quadratic form with the optimal weight matrix $\boldsymbol{\Lambda}_{1j}$:

$$\mathbf{m}(\beta)^T\boldsymbol{\Lambda}_{1j}\,\mathbf{m}(\beta) = \left[\begin{pmatrix}\hat{\beta}_{1j}\\ \hat{\beta}_{2j}\end{pmatrix} - \frac{\boldsymbol{b}_{\cdot 1}}{b_{11}}\beta_{1j}\right]^T\boldsymbol{\Lambda}_{1j}\left[\begin{pmatrix}\hat{\beta}_{1j}\\ \hat{\beta}_{2j}\end{pmatrix} - \frac{\boldsymbol{b}_{\cdot 1}}{b_{11}}\beta_{1j}\right], \tag{S35}$$

where $\mathbf{m}(\beta) := \begin{pmatrix}\hat{\beta}_{1j}\\ \hat{\beta}_{2j}\end{pmatrix} - \frac{\boldsymbol{b}_{\cdot 1}}{b_{11}}\beta$. Taking the derivative with respect to $\beta_{1j}$ and setting it to zero:

$$\begin{aligned}0 &= \frac{\partial}{\partial\beta_{1j}}\left[\mathbf{m}(\beta)^T\boldsymbol{\Lambda}_{1j}\,\mathbf{m}(\beta)\right]\\ &= -2\,\frac{\boldsymbol{b}_{\cdot 1}^T}{b_{11}}\boldsymbol{\Lambda}_{1j}\begin{pmatrix}\hat{\beta}_{1j}\\ \hat{\beta}_{2j}\end{pmatrix} + 2\,\frac{\boldsymbol{b}_{\cdot 1}^T}{b_{11}}\boldsymbol{\Lambda}_{1j}\frac{\boldsymbol{b}_{\cdot 1}}{b_{11}}\beta_{1j},\end{aligned}$$

Solving for $\beta_{1j}$ yields the estimator:

$$\hat{\beta}_{1j}^{meta} = \left(\frac{\boldsymbol{b}_{\cdot 1}^T}{b_{11}}\boldsymbol{\Lambda}_{1j}\frac{\boldsymbol{b}_{\cdot 1}}{b_{11}}\right)^{-1}\frac{\boldsymbol{b}_{\cdot 1}^T}{b_{11}}\boldsymbol{\Lambda}_{1j}\begin{pmatrix}\hat{\beta}_{1j}\\ \hat{\beta}_{2j}\end{pmatrix}. \tag{S36}$$

Based on standard GMM theory, the estimator follows the asymptotic distribution:

$$\hat{\beta}_{1j}^{meta} \mid \beta_{1j} \sim \mathcal{N}\left(\beta_{1j},\ \left[\frac{\partial\mathbf{m}^T}{\partial\beta}\boldsymbol{\Lambda}_{1j}\frac{\partial\mathbf{m}}{\partial\beta}\right]^{-1}\right), \quad \text{where } \frac{\partial\mathbf{m}^T}{\partial\beta} = \frac{\boldsymbol{b}_{\cdot 1}^T}{b_{11}}. \tag{S37}$$

Eq. (S36) shows that the meta estimator effectively borrows information from the auxiliary cohort (TPMI) to improve the estimation accuracy of the target cohort (BBJ). The augmented effect size and its standard error for trait pair $j$ in the BBJ cohort are:

$$\hat{\beta}_{1j}^{meta} = \left(\frac{\boldsymbol{b}_{\cdot 1}^T}{b_{11}} \boldsymbol{\Lambda}_{1j} \frac{\boldsymbol{b}_{\cdot 1}}{b_{11}}\right)^{-1} \frac{\boldsymbol{b}_{\cdot 1}^T}{b_{11}} \boldsymbol{\Lambda}_{1j} \begin{pmatrix} \hat{\beta}_{1j} \\ \hat{\beta}_{2j} \end{pmatrix}, \quad \hat{s}_{1j}^{meta} = \sqrt{\left[\frac{\boldsymbol{b}_{\cdot 1}^T}{b_{11}} \boldsymbol{\Lambda}_{1j} \frac{\boldsymbol{b}_{\cdot 1}}{b_{11}}\right]^{-1}}.$$

Similarly, the augmented estimates for the TPMI cohort are:

$$\hat{\beta}_{2j}^{meta} = \left(\frac{\boldsymbol{b}_{\cdot 2}^T}{b_{22}} \boldsymbol{\Lambda}_{2j} \frac{\boldsymbol{b}_{\cdot 2}}{b_{22}}\right)^{-1} \frac{\boldsymbol{b}_{\cdot 2}^T}{b_{22}} \boldsymbol{\Lambda}_{2j} \begin{pmatrix} \hat{\beta}_{1j} \\ \hat{\beta}_{2j} \end{pmatrix}, \quad \hat{s}_{2j}^{meta} = \sqrt{\left[\frac{\boldsymbol{b}_{\cdot 2}^T}{b_{22}} \boldsymbol{\Lambda}_{2j} \frac{\boldsymbol{b}_{\cdot 2}}{b_{22}}\right]^{-1}},$$

where $\boldsymbol{\Lambda}_{2j}^{-1} := \boldsymbol{B} - \frac{\boldsymbol{b}_{\cdot 2}\boldsymbol{b}_{\cdot 2}^T}{b_{22}} + \hat{\mathbf{E}}_j$ and $\boldsymbol{b}_{\cdot 2} = \begin{pmatrix} b_{12} \\ b_{22} \end{pmatrix}$. As a sanity check, when $b_{12} = 0$ (i.e., the true effects in the two cohorts are uncorrelated), the meta estimator reduces to the original single-cohort estimator for each trait pair, thereby avoiding the incorporation of uninformative noise from the other cohort (see Xiao et al. [4], Supplementary Methods, section 3.6).

Finally, when a given MR method produced a valid estimate in only one of the two cohorts for a particular trait pair (e.g., by MR-Lasso), we retained the original single-cohort estimate without performing the meta-analysis.

### 3 Supplementary tables

#### 3.1 Heterogeneous effects between EUR and EAS

Table S1 lists the trait pairs that achieved statistical significance (after Benjamini–Hochberg [BH] correction) in at least one of the European and East Asian populations and exhibited statistically significant heterogeneity in causal effect size between the two populations. For the EAS estimates, we used augmented effect sizes derived from the meta-analysis of the BBJ and TPMI cohorts (section 2.3.4), with BBJ serving as the reference dataset. Rows shaded in red denote trait pairs that were statistically significant in the BBJ cohort and further validated in the TPMI cohort, meaning that both the causal estimate and its heterogeneity relative to EUR remained significant in the independent replication sample.

| Exposure | Outcome | $\beta_{EAS}$ | $SE_{EAS}$ | $P_{EAS}$ | $\beta_{EUR}$ | $SE_{EUR}$ | $P_{EUR}$ | Category |
|---|---|---|---|---|---|---|---|---|
| Body mass index | Type 2 diabetes | 0.3138 | 0.0289 | 1.5123e-27 | 0.1975 | 0.0216 | 5.5873e-20 | Significant in both populations |
| Body mass index | Iron deficiency anemia | -0.0182 | 0.0153 | 0.2338 | 0.0541 | 0.0089 | 1.4053e-9 | Only significant in EUR |
| Body mass index | Asthma | 0.0468 | 0.0175 | 0.0073 | 0.1154 | 0.0116 | 3.8479e-23 | Significant in both populations |
| Body mass index | Chronic obstructive pulmonary disease | 0.0084 | 0.0172 | 0.6273 | 0.1169 | 0.0109 | 9.1244e-27 | Only significant in EUR |
| Body mass index | Cholelithiasis | 0.0729 | 0.0179 | 4.6511e-5 | 0.1256 | 0.0109 | 7.203e-31 | Significant in both populations |
| Body mass index | Rheumatoid arthritis | 0.0050 | 0.0164 | 0.7616 | 0.0648 | 0.0100 | 8.6772e-11 | Only significant in EUR |
| Hemoglobin | Iron deficiency anemia | -0.0961 | 0.0168 | 1.0423e-8 | -0.0504 | 0.0080 | 2.4617e-10 | Significant in both populations |
| Alanine aminotransferase | Chronic obstructive pulmonary disease | -0.0936 | 0.0244 | 0.0001 | 0.0169 | 0.0137 | 0.2189 | Only significant in EAS |
| Alanine aminotransferase | Cholelithiasis | 0.0387 | 0.0275 | 0.1599 | 0.1151 | 0.0252 | 4.7466e-6 | Only significant in EUR |
| Glucose | Type 2 diabetes | 0.9152 | 0.0877 | 1.7343e-25 | 0.6103 | 0.0618 | 5.1982e-23 | Significant in both populations |
| Glucose | Iron deficiency anemia | -0.0438 | 0.0229 | 0.0562 | 0.0372 | 0.0143 | 0.0091 | Only significant in EUR |
| Glucose | Epilepsy | -0.0713 | 0.0233 | 0.0022 | 0.0051 | 0.0145 | 0.7230 | Only significant in EAS |
| Glucose | Cataract | 0.1390 | 0.0315 | 1.0474e-5 | 0.0419 | 0.0193 | 0.0299 | Only significant in EAS |
| Glucose | Asthma | -0.1226 | 0.0285 | 1.6747e-5 | -0.0015 | 0.0197 | 0.9396 | Only significant in EAS |
| HbA1c | Type 2 diabetes | 0.7152 | 0.0447 | 1.6106e-57 | 0.3516 | 0.0273 | 6.3884e-38 | Significant in both populations |
| HbA1c | Iron deficiency anemia | -0.0171 | 0.0154 | 0.2657 | 0.0392 | 0.0090 | 1.3758e-5 | Only significant in EUR |
| HbA1c | Epilepsy | -0.0395 | 0.0151 | 0.0087 | -0.0014 | 0.0083 | 0.8620 | Only significant in EAS |
| HbA1c | Cataract | 0.1217 | 0.0200 | 1.1441e-9 | 0.0158 | 0.0099 | 0.1121 | Only significant in EAS |
| HbA1c | Asthma | -0.0519 | 0.0188 | 0.0058 | 0.0016 | 0.0125 | 0.8957 | Only significant in EAS |
| HbA1c | Chronic obstructive pulmonary disease | -0.0473 | 0.0175 | 0.0071 | 0.0120 | 0.0094 | 0.2019 | Only significant in EAS |
| LDL cholesterol | Myocardial infarction | 0.1939 | 0.0298 | 7.9152e-11 | 0.0882 | 0.0187 | 2.5823e-6 | Significant in both populations |
| Diastolic blood pressure | Breast cancer | -0.1222 | 0.0350 | 0.0005 | -0.0118 | 0.0176 | 0.5018 | Only significant in EAS |
| Diastolic blood pressure | Type 2 diabetes | -0.0762 | 0.0317 | 0.0162 | 0.0577 | 0.0167 | 0.0006 | Significant in both populations |
| Diastolic blood pressure | Rheumatoid arthritis | -0.0368 | 0.0242 | 0.1279 | 0.0347 | 0.0122 | 0.0046 | Only significant in EUR |

Table S1: **Heterogeneous trait pairs between EAS (BBJ) and EUR populations.** $\beta$, $SE$, and $P$ denote the estimated causal effect size, standard error, and $P$ value, respectively. Significance is determined after BH correction. $\beta_{EAS}$ values are the meta-analyzed effect estimates combining BBJ and TPMI, with BBJ as the reference cohort. Red-shaded rows indicate pairs that are significant in the primary EAS cohort (BBJ) and independently validated in the auxiliary cohort (TPMI), with both the causal effect and the effect heterogeneity relative to EUR reaching statistical significance. Abbreviations: LDL cholesterol, low-density lipoprotein cholesterol; HbA1c, glycated hemoglobin.

#### 3.2 Heterogeneous effects between EUR and CSA

Table S2 lists the trait pairs that achieved statistical significance in at least one of the EUR and Central/South Asian populations (after BH correction) and exhibited statistically significant heterogeneity in causal effect size between the two groups.

| Exposure | Outcome | $\beta_{CSA}$ | $SE_{CSA}$ | $P_{CSA}$ | $\beta_{EUR}$ | $SE_{EUR}$ | $P_{EUR}$ | Category |
|---|---|---|---|---|---|---|---|---|
| Hypertension | SHBG | 0.0022 | 0.0225 | 0.9219 | -0.1275 | 0.0346 | 0.0002 | Only significant in EUR |
| T2D | HbA1c | 0.9456 | 0.1709 | 3.1753e-8 | 1.5124 | 0.1133 | 1.1318e-40 | Significant in both populations |
| T2D | HDL cholesterol | -0.079 | 0.1229 | 0.5200 | -0.5633 | 0.1391 | 5.1463e-5 | Only significant in EUR |
| T2D | SHBG | -0.323 | 0.1609 | 0.0447 | -0.7584 | 0.0898 | 2.9125e-17 | Only significant in EUR |
| Disorders of lipoid metabolism | Vitamin D | 0.0198 | 0.1515 | 0.8960 | -0.3566 | 0.0796 | 7.5259e-6 | Only significant in EUR |
| HbA1c | Hypertension | 0.1348 | 0.0414 | 0.0011 | 0.0458 | 0.0135 | 0.0007 | Significant in both populations |
| HbA1c | T2D | 0.4216 | 0.0453 | 1.3714e-30 | 0.2009 | 0.0140 | 1.2739e-46 | Significant in both populations |
| Height | Disorders of lipoid metabolism | -0.0007 | 0.0051 | 0.8882 | -0.0229 | 0.0047 | 8.9485e-7 | Only significant in EUR |
| Chronic ischaemic heart disease | Hypertension | 0.047 | 0.1270 | 0.7113 | 0.4539 | 0.0909 | 5.9216e-7 | Only significant in EUR |

Table S2: **Heterogeneous trait pairs between CSA and EUR populations.** $\beta$, $SE$, and $P$ denote the estimated causal effect size, standard error, and $P$ value, respectively. Significance is determined after BH correction. Abbreviations: T2D, type 2 diabetes; SHBG, sex hormone-binding globulin; HDL cholesterol, high-density lipoprotein cholesterol; HbA1c, glycated hemoglobin.

#### 3.3 Heterogeneous effects between EUR and AFR

Table S3 lists the trait pairs that achieved statistical significance in at least one of the EUR and African populations (after BH correction) and that showed statistically significant heterogeneity in causal effect size between the two groups.

| Exposure | Outcome | $\beta_{AFR}$ | $SE_{AFR}$ | $P_{AFR}$ | $\beta_{EUR}$ | $SE_{EUR}$ | $P_{EUR}$ | Category |
|---|---|---|---|---|---|---|---|---|
| Hypertension | Chronic ischaemic heart disease | -0.0123 | 0.0559 | 0.8257 | 0.2670 | 0.0223 | 4.3567e-33 | Only significant in EUR |
| Disorders of lipoid metabolism | LDL cholesterol | 0.8112 | 0.3337 | 0.0151 | 1.6601 | 0.2037 | 3.6483e-16 | Only significant in EUR |
| Disorders of lipoid metabolism | Chronic ischaemic heart disease | 0.2857 | 0.1289 | 0.0267 | 0.6361 | 0.0845 | 5.1389e-14 | Only significant in EUR |
| C-reactive protein | Hypertension | -0.0661 | 0.0508 | 0.1932 | 0.0471 | 0.0204 | 0.0207 | Only significant in EUR |
| LDL cholesterol | Disorders of lipoid metabolism | 0.0816 | 0.0603 | 0.1765 | 0.3131 | 0.0225 | 7.1635e-44 | Only significant in EUR |
| Albumin | Hypertension | 0.3746 | 0.1057 | 0.0004 | 0.0464 | 0.0210 | 0.0270 | Only significant in AFR |

Table S3: **Heterogeneous trait pairs between AFR and EUR populations.** $\beta$, $SE$, and $P$ denote the estimated causal effect size, standard error, and $P$ value, respectively. Significance is determined after BH correction. Abbreviations: LDL cholesterol, low-density lipoprotein cholesterol.

### 4 Supplementary figures

#### 4.1 Simulation results in null cases for additional methods

We extended our simulation studies to evaluate the performance of additional MR methods under the null hypothesis ($\beta = 0$), focusing on methods that were not included in the main-text comparisons. The methods evaluated here include ConMix, dIVW, IVW-fe, MR-Robust, MR-PRESSO, and MR-Lasso. To ensure a sufficient number of IVs for causal estimation and to align with the strategy adopted in our real-data analyses, we used $P \leq 5 \times 10^{-5}$ as the IV selection threshold for all methods in these simulations.

As shown in Fig. S2, all evaluated methods exhibited severe inflation, regardless of the genetic correlation ($\rho$) between the populations. Notably, MR-Lasso occasionally classified all IVs as invalid outliers, resulting in the method failing to produce a causal estimate in those instances.

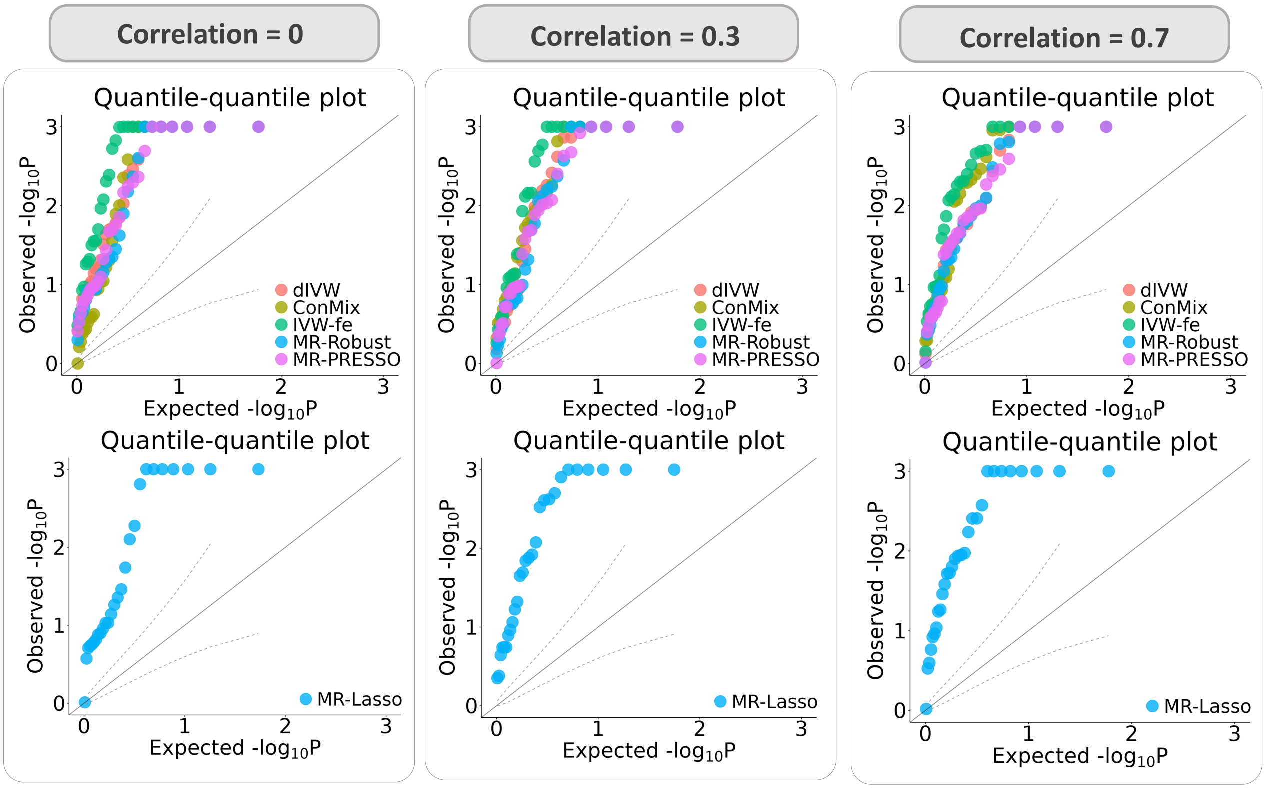

Figure S2: **Negative control simulations for additional MR methods.** QQ plots of $-\log_{10}(p)$ values from each method are displayed under the null scenario ($\beta = 0$), where no causal effect exists. The genetic correlation parameter $\rho$ between the two populations varies from 0 to 0.7.

#### 4.2 Ablation study of XMR components under null simulations

To systematically evaluate the contribution of each component within the XMR framework to confounding correction, we conducted an ablation study under the null hypothesis ($\beta = 0$). We examined the performance of three variants, each omitting a specific modeling assumption:

- **XMR ($\Omega = 0$):** The variance components for pleiotropic effects ($u_{1,j}, u_{2,j}, v_{2,j}$) were fixed at zero.

- **XMR ($C = I$):** The sample structure was ignored by enforcing a diagonal covariance matrix (i.e., zero off-diagonal elements).

- **XMR (not correct bias):** The selection bias adjustment was omitted by setting the conditional probability threshold to $P \leq 1$ (equivalently, $t = 0$ in Eq. (S4)).

As shown in Fig. S3, none of the three variants maintained well-calibrated $P$ values across all genetic correlation settings. These results underscore the necessity and effectiveness of each component in the XMR framework for controlling false positives.

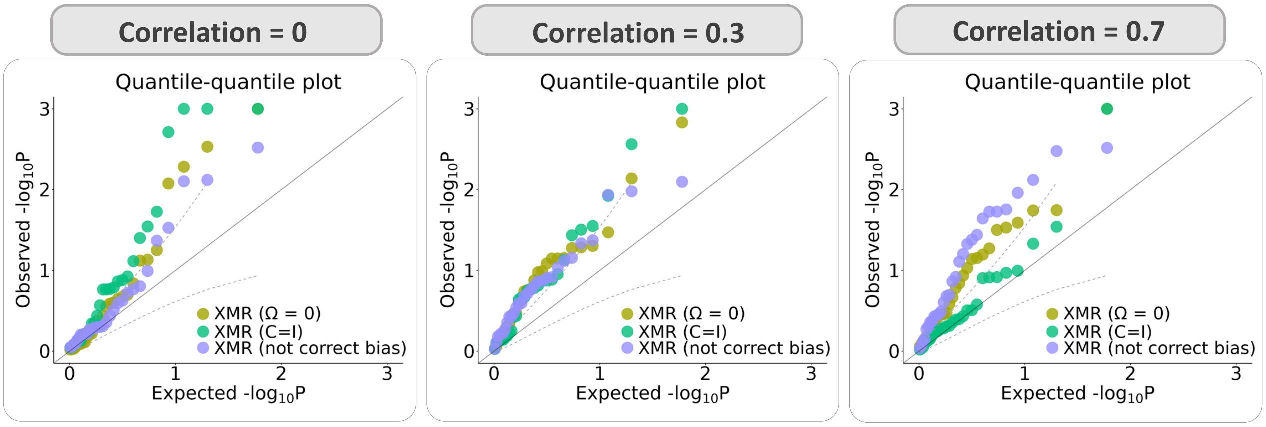

Figure S3: **XMR ablation study under null simulations.** The distribution of $-\log_{10}(p)$ values is shown from the three variants of the XMR model under the null scenario ($\beta = 0$). The genetic correlation parameter $\rho$ between the two populations varies from 0 to 0.7.

#### 4.3 Extended simulation results under alternative hypotheses

This section presents the complete causal effect estimates for the five methods that demonstrated well-controlled false positive rates in the null simulations: XMR, MRAPSS, Egger, Weighted-mode, and CAUSE. The true causal effect $\beta$ was varied across the set $\{0.05, 0.1, 0.15, 0.2, 0.25, 0.3\}$, and the genetic correlation $\rho$ was set to 0, 0.3, and 0.7.

XMR consistently achieved high estimation accuracy across all simulation settings. Notably, its performance advantage over other methods became increasingly pronounced as the genetic correlation $\rho$ and the causal effect size $\beta$ increased.

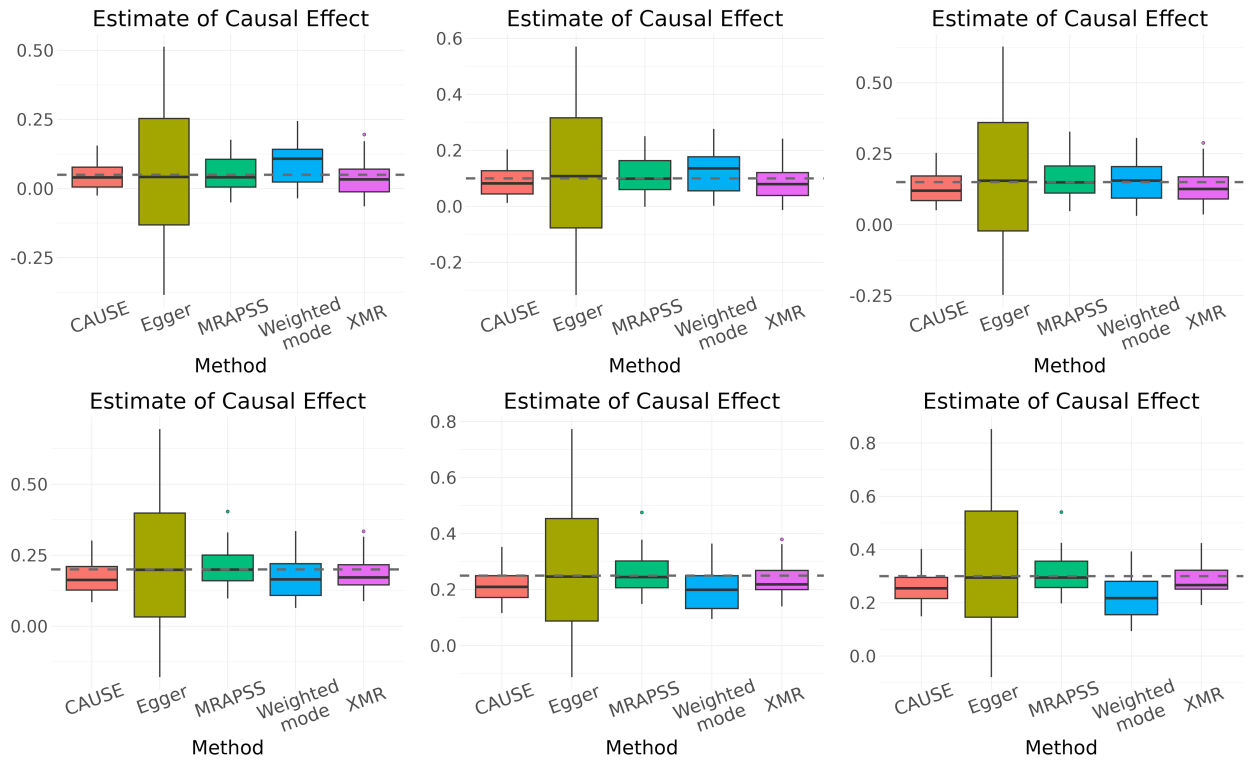

Figure S4: **Causal effect estimates under genetic correlation $\rho = 0$.** Boxplots representing the distribution of estimates from 30 independent experiments by five methods. The dashed horizontal lines indicate the true causal effect size $\beta \in \{0.05, 0.1, 0.15, 0.2, 0.25, 0.3\}$.

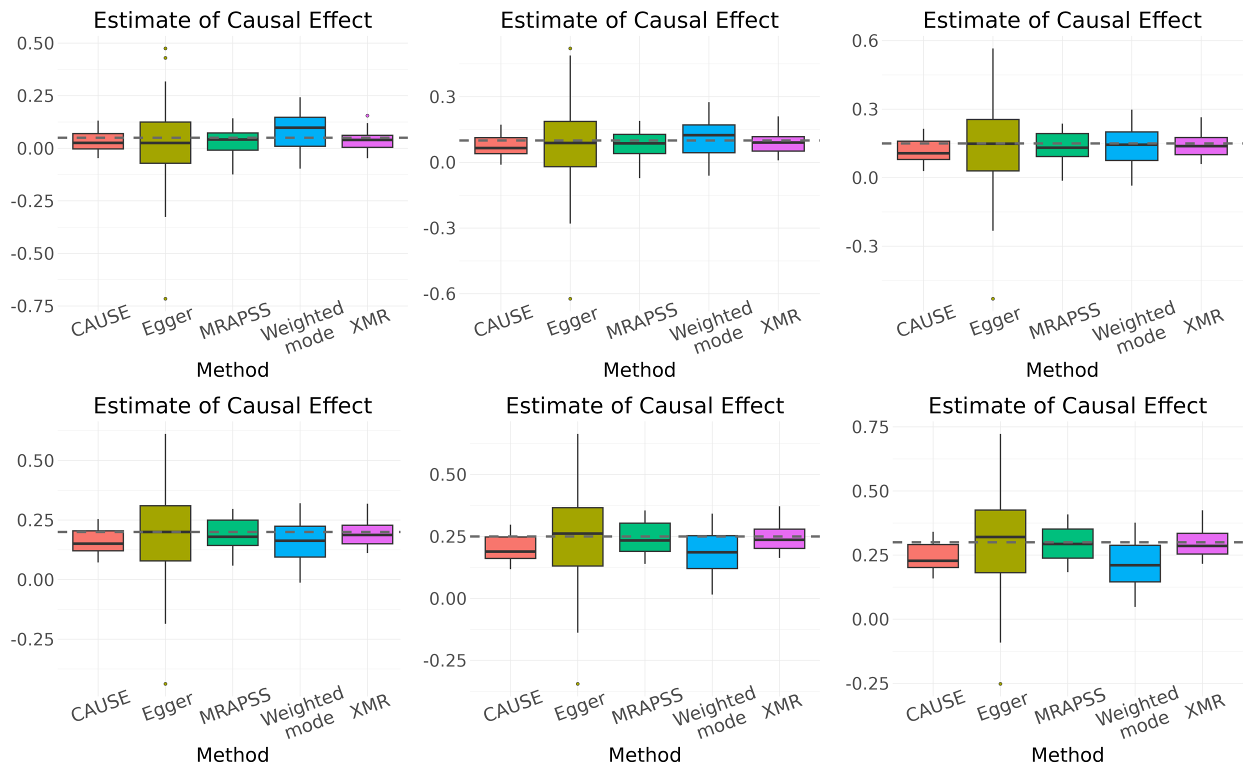

Figure S5: **Causal effect estimates under genetic correlation $\rho = 0.3$.** Boxplots representing the distribution of estimates from 30 independent experiments by five methods. The dashed horizontal lines indicate the true causal effect size $\beta \in \{0.05, 0.1, 0.15, 0.2, 0.25, 0.3\}$.

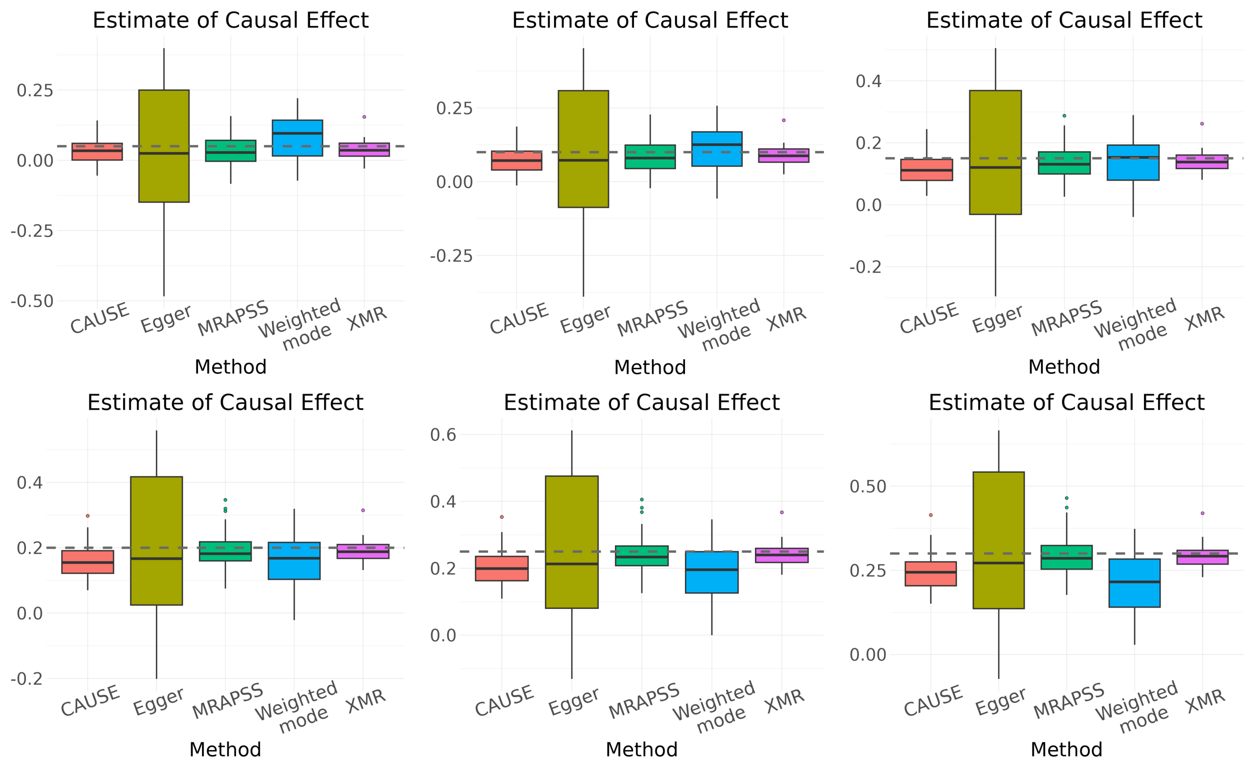

Figure S6: **Causal effect estimates under genetic correlation $\rho = 0.7$.** Boxplots representing the distribution of estimates from 30 independent experiments by five methods. The dashed horizontal lines indicate the true causal effect size $\beta \in \{0.05, 0.1, 0.15, 0.2, 0.25, 0.3\}$.

#### 4.4 Performance of additional methods in real-data negative control studies

We extended the real-data negative control analysis to include additional single-population MR methods. Because the available datasets for the AFR and CSA populations have limited sample sizes (below 10,000), the standard genome-wide significance threshold ($5 \times 10^{-8}$) yielded an insufficient number of IVs. We therefore relaxed the inclusion threshold and evaluated each method's performance under three distinct $P$ value cutoffs.

Among the tested methods, only Egger, Weighted-mode, CAUSE, and Weighted-median avoided severe inflation in both AFR and CSA populations. The remaining methods faced a trade-off, suffering from significant inflation when the IV threshold was relaxed to include more variants, consistent with the findings discussed in the main text.

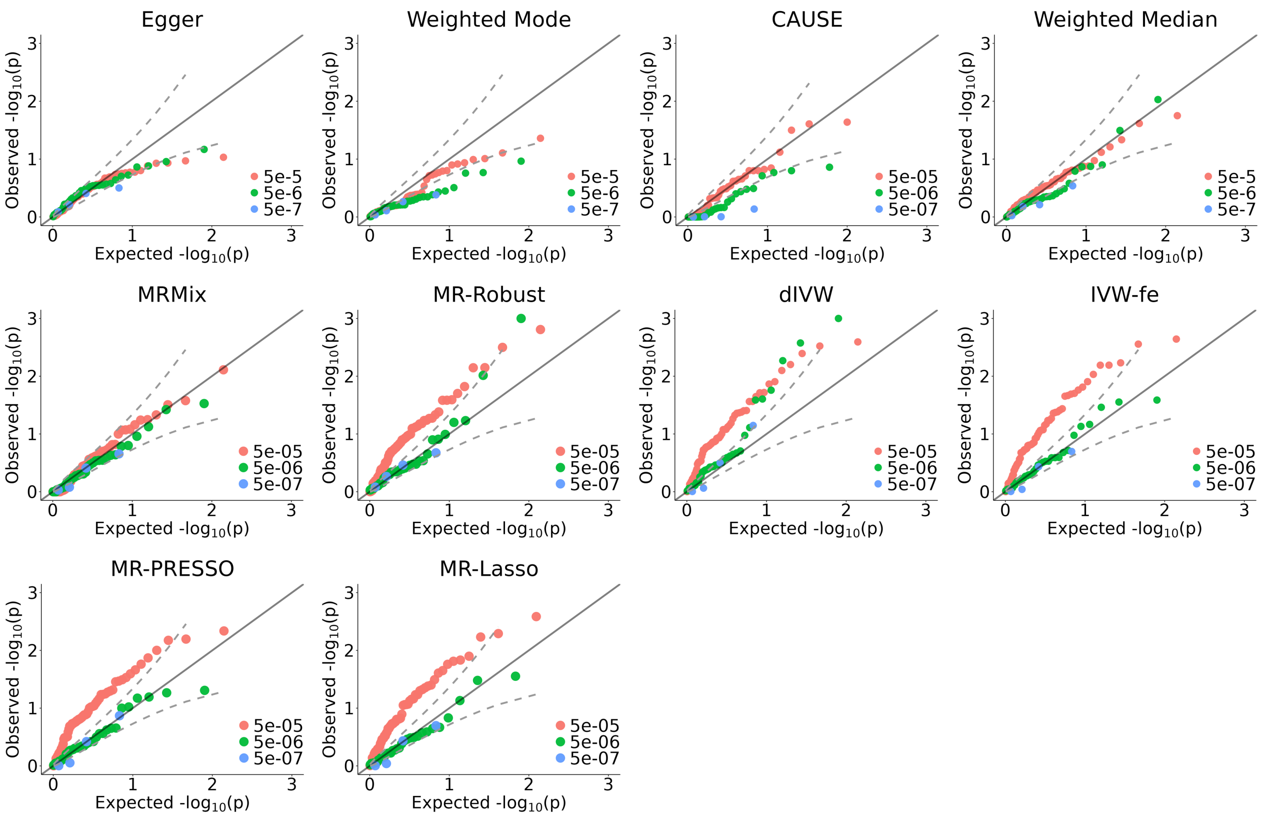

Figure S7: **Real-data negative control studies in the AFR population.** QQ plots of $-\log_{10}(p)$ values from various single-population MR methods, evaluated under different IV selection thresholds.

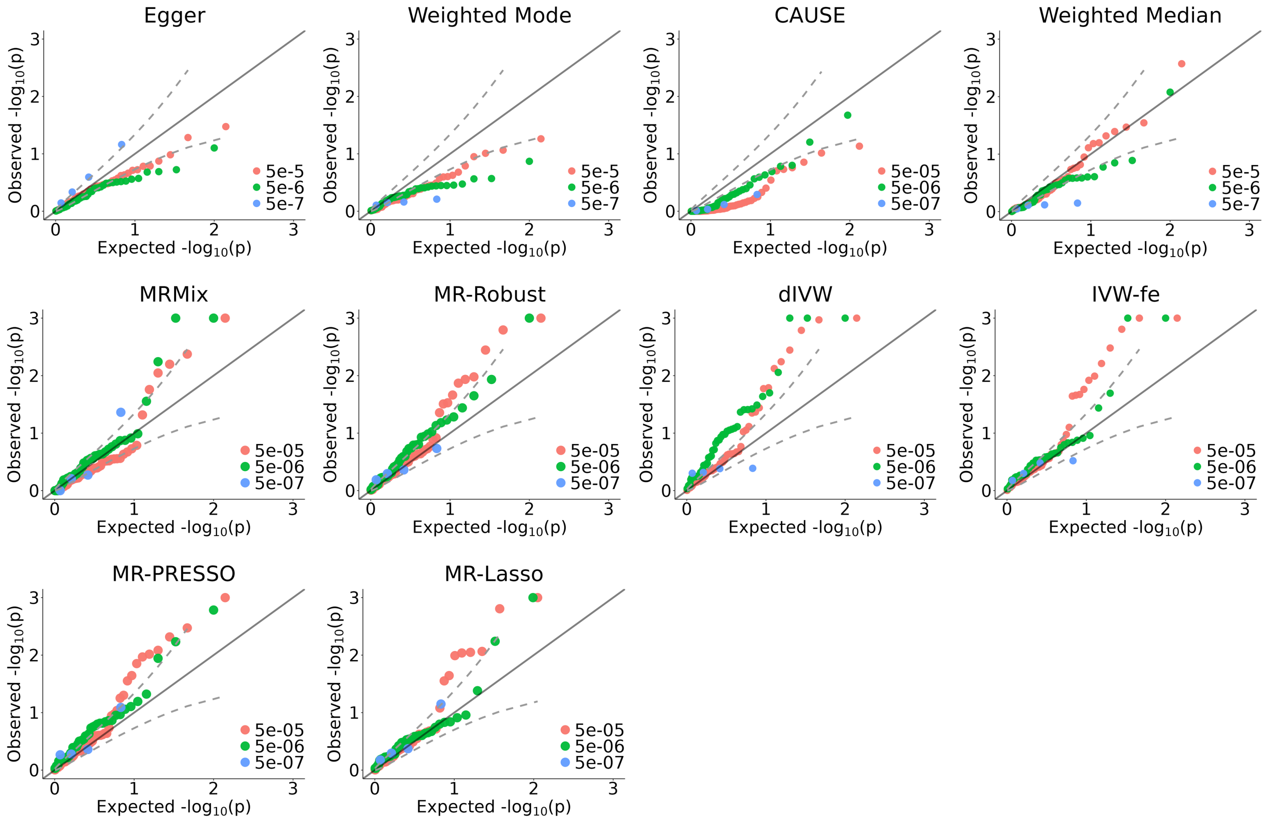

Figure S8: **Real-data negative control studies in the CSA population.** QQ plots of $-\log_{10}(p)$ values from various single-population MR methods, evaluated under different IV selection thresholds.

#### 4.5 Ablation study of XMR variants in real-data negative controls

We validated the ablation study findings using real-data negative controls. We compared the full XMR model against its three variants: XMR ($\boldsymbol{\Omega} = 0$), XMR ($\boldsymbol{C} = I$), and XMR (not correct bias), using an IV selection threshold of $P \leq 5 \times 10^{-5}$.

Fig. S9 illustrates the critical role of each model component. Consistent with the simulation results, only the full version of XMR produced well-calibrated $P$ values across both populations, whereas the variants exhibited varying degrees of inflation. Detailed causal effect estimates from the three XMR variants are listed in Supplementary Tables S7 and S12.

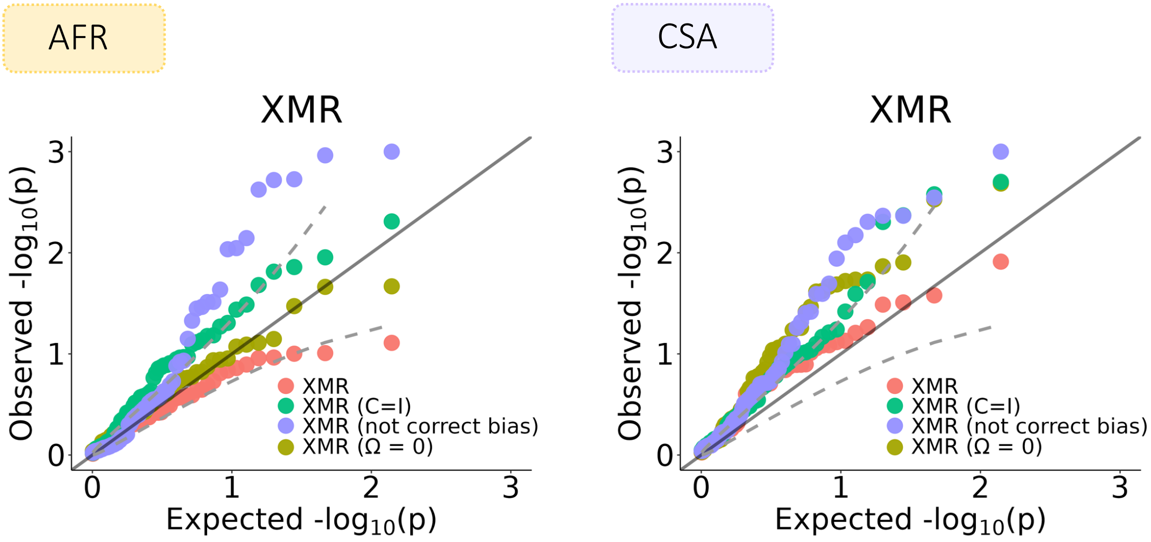

Figure S9: **XMR ablation study in real-data negative control studies.** The distribution of $-\log_{10}(p)$ values is shown from the full XMR model and its three variants in both AFR and CSA populations.

##### 4.6 Real-data causal relationship exploration in East Asians

This section presents supplementary results from the real-data analysis focused on the EAS population, using the BioBank Japan (BBJ) and Taiwan Precision Medicine Initiative (TPMI) datasets specifically.

###### 4.6.1 Comparison of discovery numbers in BBJ across methods

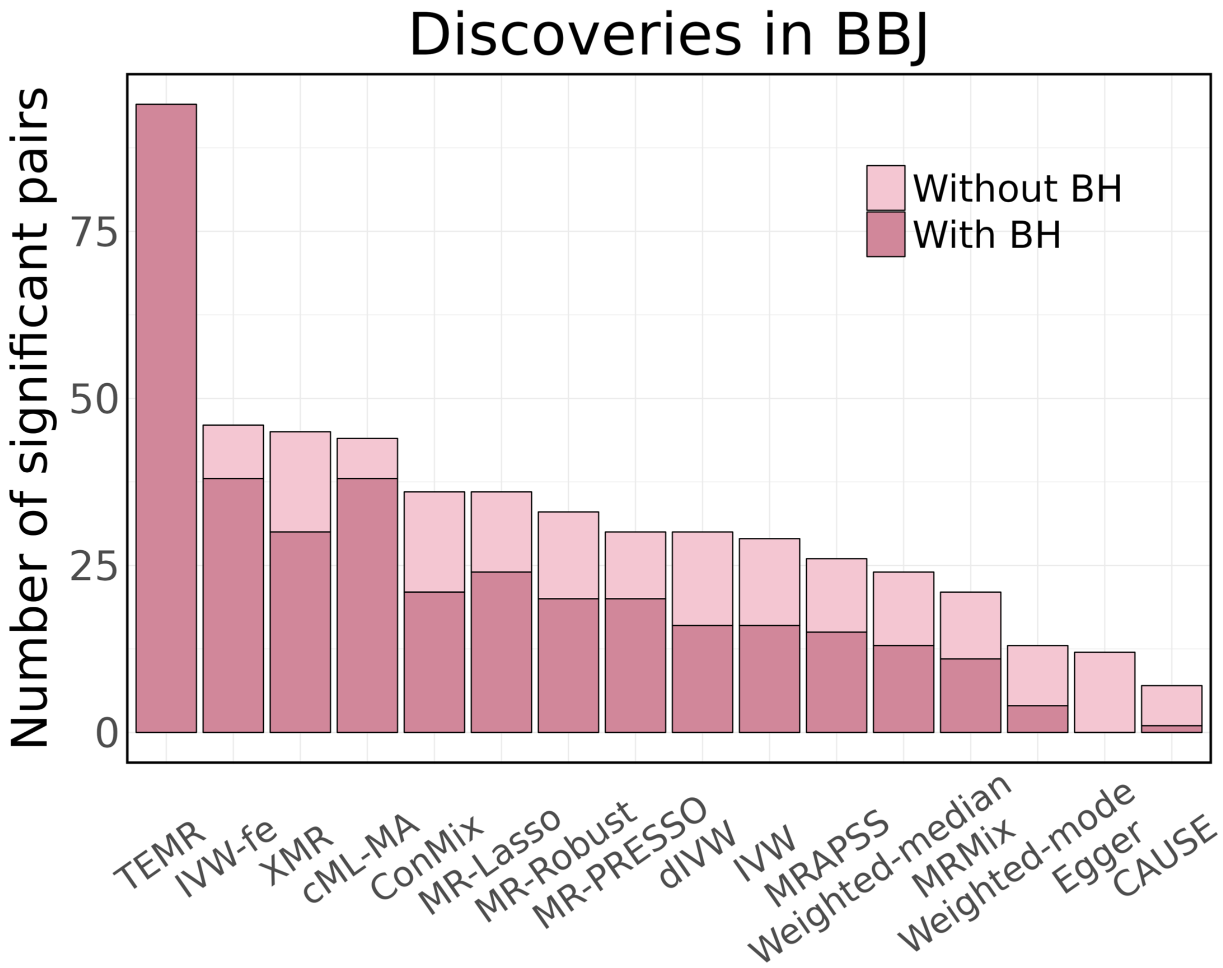

Figure S10: **Number of significant discoveries in the BBJ cohort.** The number of significant causal pairs identified by each of the 16 tested methods is shown. Light pink bars represent discoveries before multiple-testing correction, and dark pink bars represent discoveries after BH correction.

##### 4.6.2 Extended causal effect estimates in BBJ

Fig. S11 presents the causal discoveries in the BBJ cohort using MRAPSS, Egger, Weighted-mode, and CAUSE. Fig. S12 displays results of the remaining methods, which exhibited inflation in either the simulation studies or the negative control analyses.

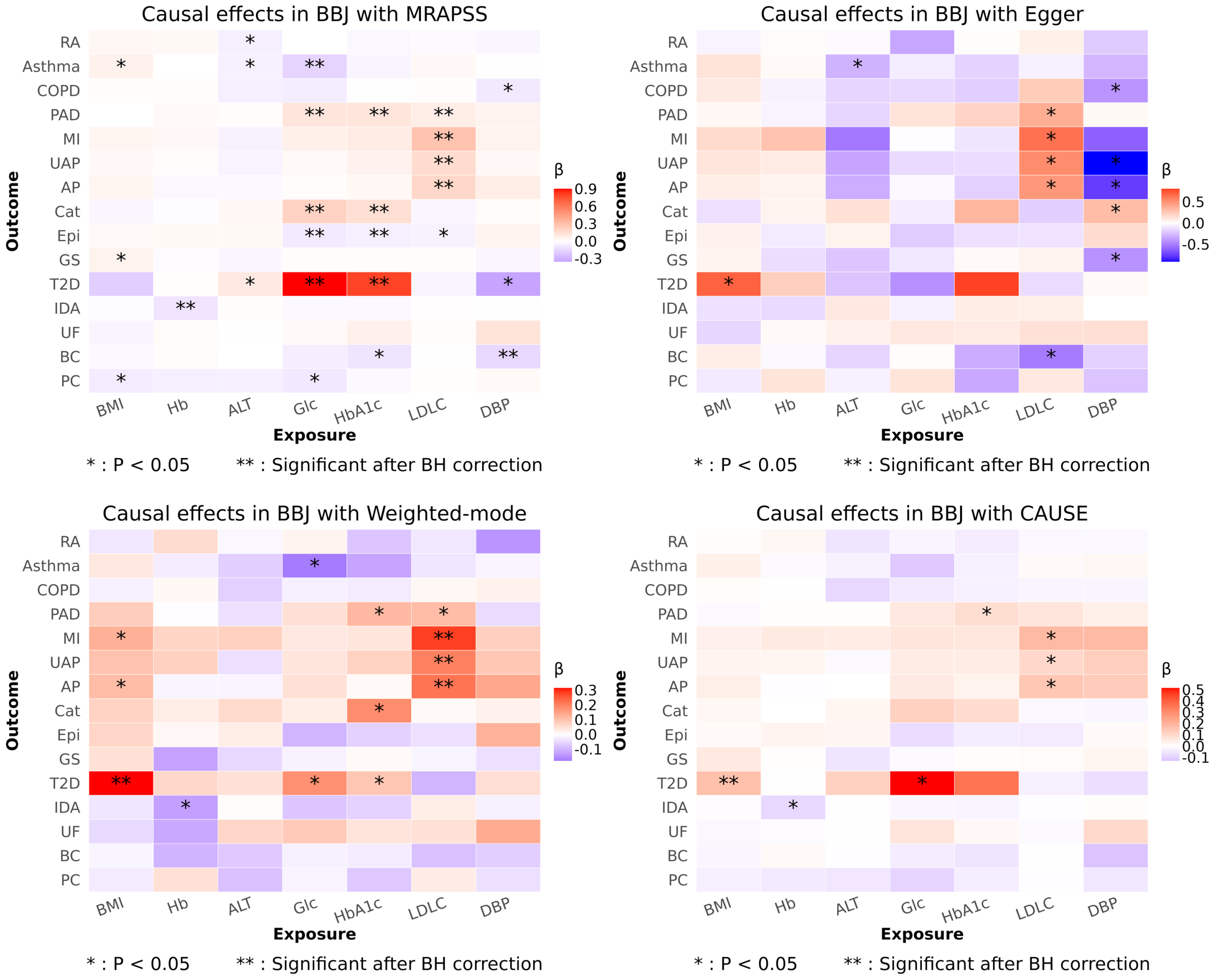

Figure S11: **Causal effect estimates in BBJ.** Causal effect estimates ($\beta$) are shown across MRAPSS, Egger, Weighted-mode, and CAUSE. Red indicates positive effects, while purple indicates negative effects. Statistical significance is denoted by * ($P < 0.05$) and ** (significant after BH correction).

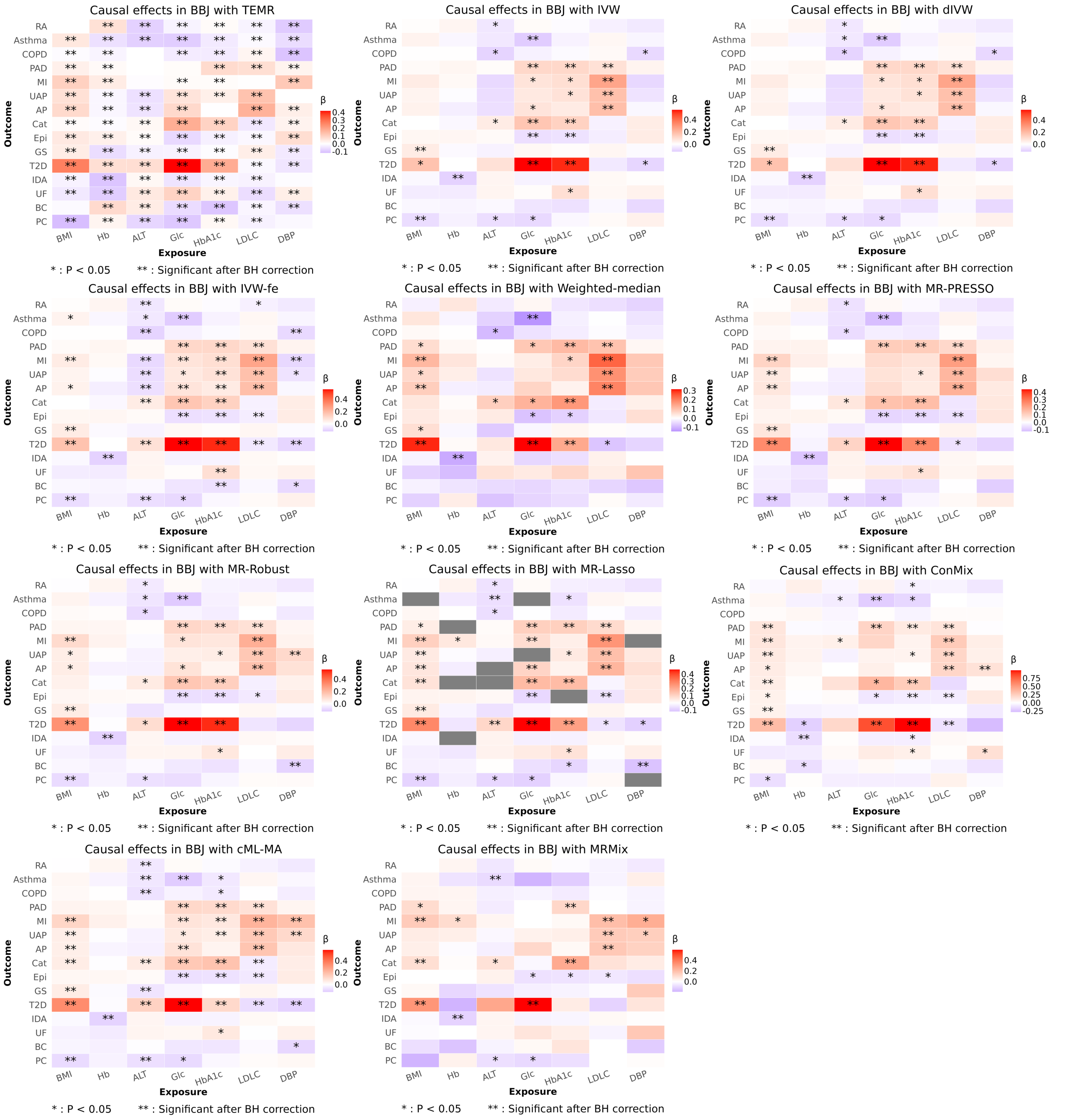

Figure S12: **Causal effect estimates in BBJ.** Causal effect estimates ($\beta$) are shown across the remaining methods. Red indicates positive effects, while purple indicates negative effects. Statistical significance is denoted by * ($P < 0.05$) and ** (significant after BH correction).

###### 4.6.3 Replication consistency between two East Asian cohorts

We evaluated the consistency of causal effect estimates between the BBJ and TPMI cohorts. Fig. S13 compares the estimates from MRAPSS, Egger, Weighted-mode, and CAUSE, while Fig. S14 provides the comparison of the other 10 methods.

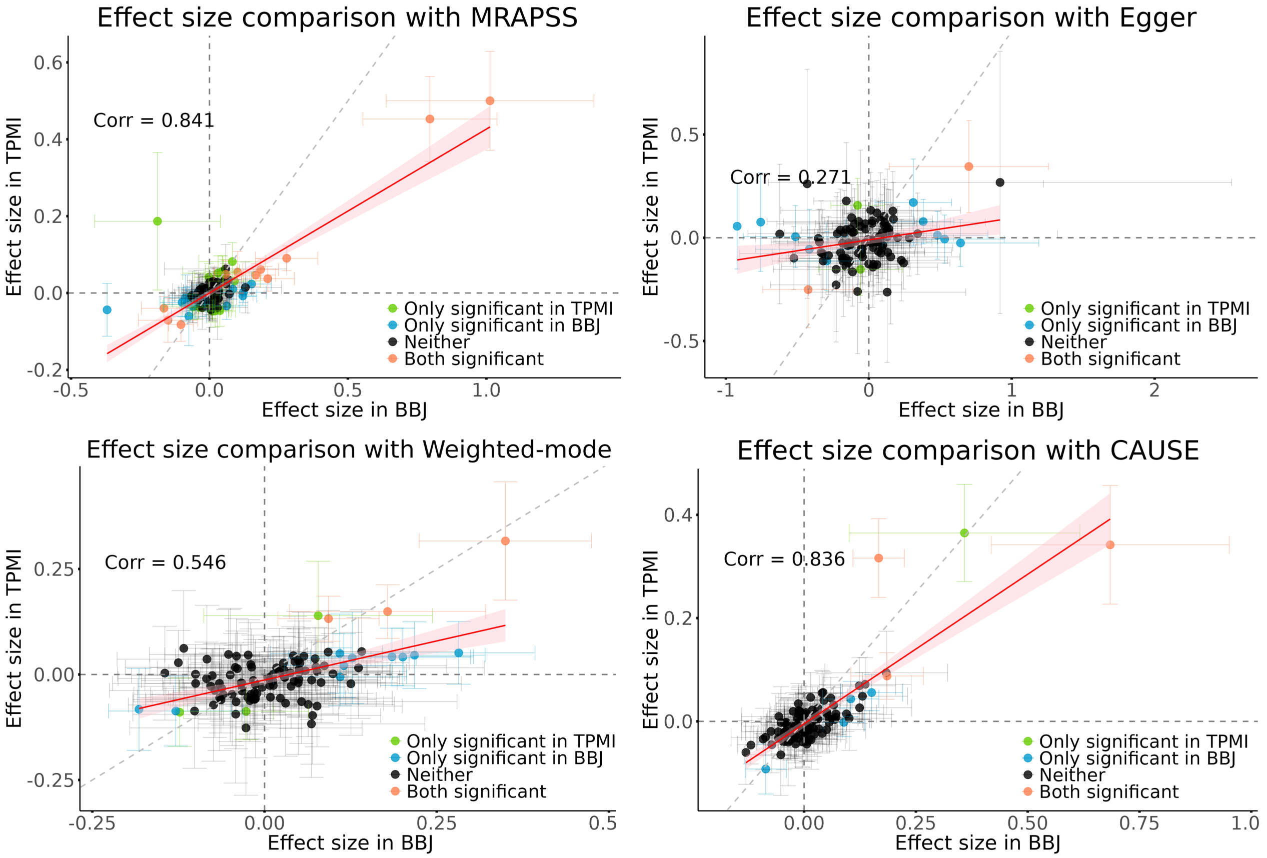

Figure S13: **Consistency of causal effect estimates between BBJ and TPMI.** Scatter plots comparing the effect sizes estimated in BBJ versus TPMI from MRAPSS, Egger, Weighted-mode, and CAUSE. Error bars represent 95% confidence intervals. Point colors indicate significance status across the two datasets. The red line represents the linear regression fit with a 95% confidence interval (shaded area), and the grey dashed line indicates the identity line ($y = x$).

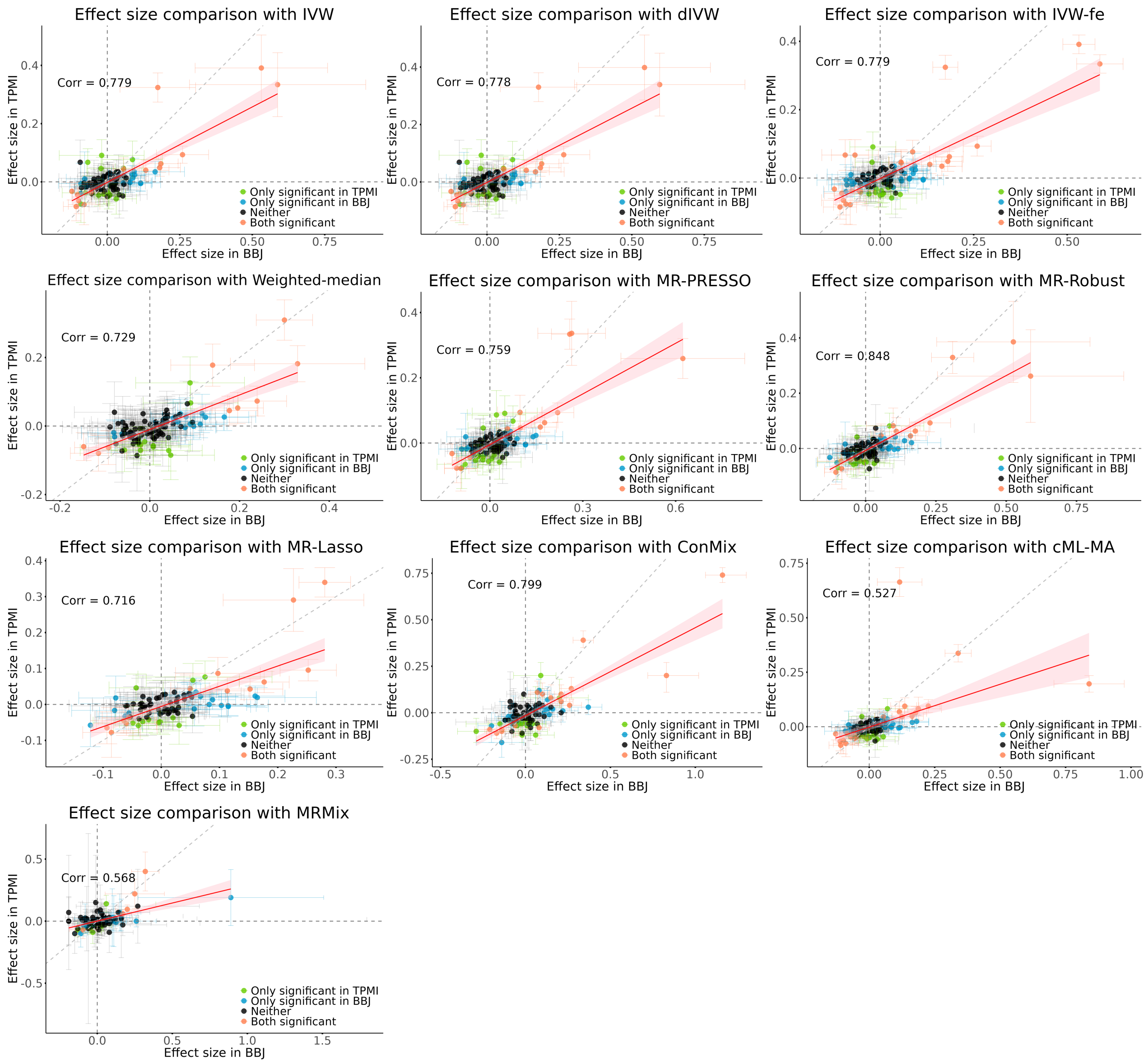

Figure S14: **Consistency of causal effect estimates between BBJ and TPMI.** Scatter plots comparing the effect sizes estimated in BBJ versus TPMI from the other 10 methods. Error bars represent 95% confidence intervals. Point colors indicate significance status across the two datasets. The red line represents the linear regression fit with a 95% confidence interval (shaded area), and the grey dashed line indicates the identity line ($y = x$).

##### 4.6.4 Identification of heterogeneous causal pairs via meta-analysis

Using estimates refined by meta-analysis ($\hat{\beta}^{meta}$), we highlighted causal pairs that exhibit heterogeneity compared to the European population. Fig. S15 displays the results of XMR, and Fig. S16 displays the results of MRAPSS, Egger, Weighted-mode, and CAUSE.

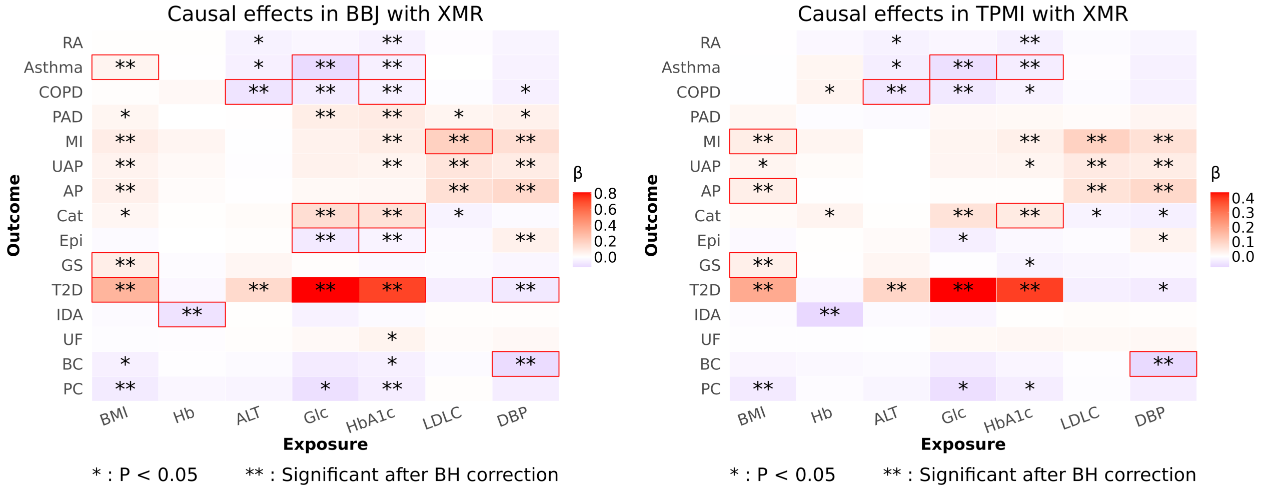

Figure S15: **Meta-analyzed causal effect estimates in EAS cohorts.** Heatmaps displaying the meta-analyzed causal effect estimates ($\hat{\beta}^{meta}$) from XMR in both BBJ and TPMI. Red indicates positive effects, and purple indicates negative effects. Significance is marked by * ($P < 0.05$) and ** (significant after BH correction). Among the significant pairs (**), red frames highlight those showing statistically significant heterogeneity compared to the EUR population.

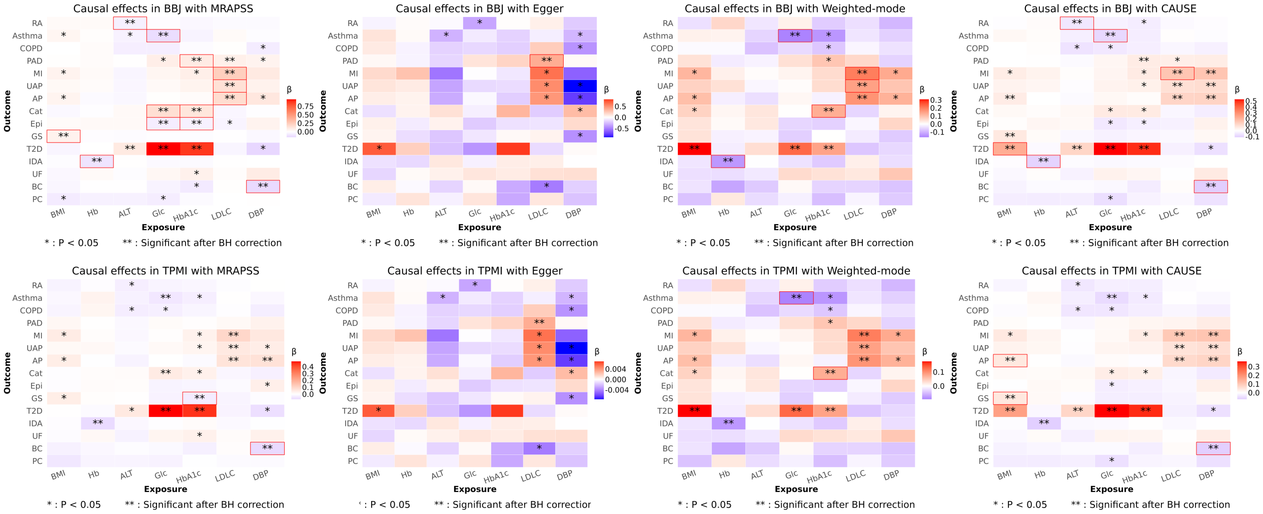

Figure S16: **Meta-analyzed causal effect estimates in EAS cohorts.** Heatmaps displaying the meta-analyzed causal effect estimates ($\hat{\beta}^{meta}$) from MRAPSS, Egger, Weighted-mode, and CAUSE in both BBJ and TPMI. Red indicates positive effects, and purple indicates negative effects. Significance is marked by * ($P < 0.05$) and ** (significant after BH correction). Among the significant pairs (**), red frames highlight those showing statistically significant heterogeneity compared to the EUR population.

##### 4.6.5 Comparison of effect sizes between EAS and EUR populations

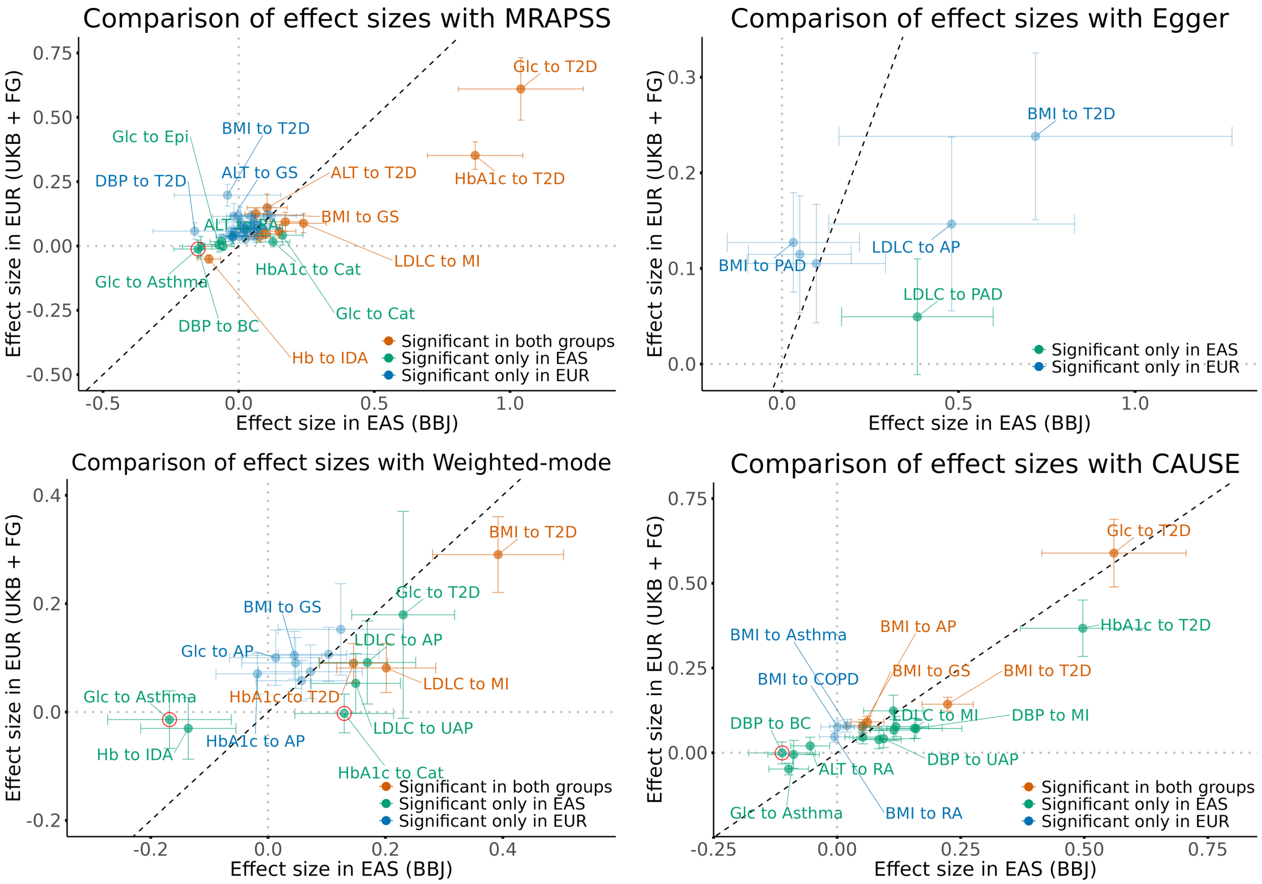

Figure S17: **Comparison of causal effect estimates between EAS and EUR.** Scatter plots comparing effect sizes in the EAS (BBJ) cohort (meta-analyzed) versus the EUR cohort from MRAPSS, Egger, Weighted-mode, and CAUSE. Error bars represent 95% confidence intervals. Point colors indicate significance status in the two populations. The black dashed line represents the identity line ($y = x$). Red circles highlight heterogeneous pairs that were validated by the TPMI dataset.

#### 4.7 Investigation of causal relationships in the Central/South Asian population

##### 4.7.1 Comparison of effect sizes between CSA and EUR populations

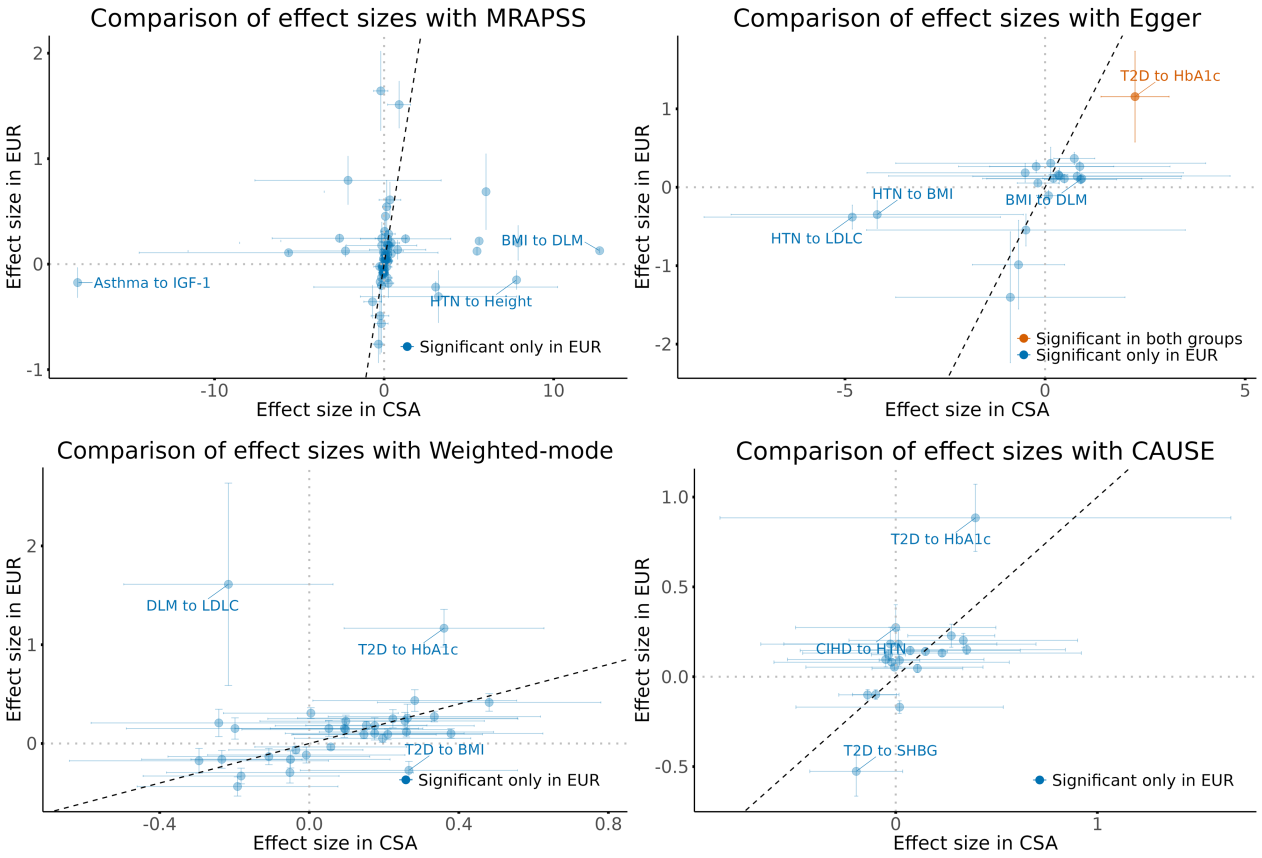

Figure S18: **Comparison of causal effect estimates between CSA and EUR.** Scatter plots comparing effect sizes in the CSA cohort versus the EUR cohort from MRAPSS, Egger, Weighted-mode, and CAUSE. Error bars represent 95% confidence intervals. Point colors indicate significance status in the two populations. The black dashed line represents the identity line ($y = x$).

#### 4.8 Investigation of causal relationships in the African population

##### 4.8.1 Comparison of effect sizes between AFR and EUR populations

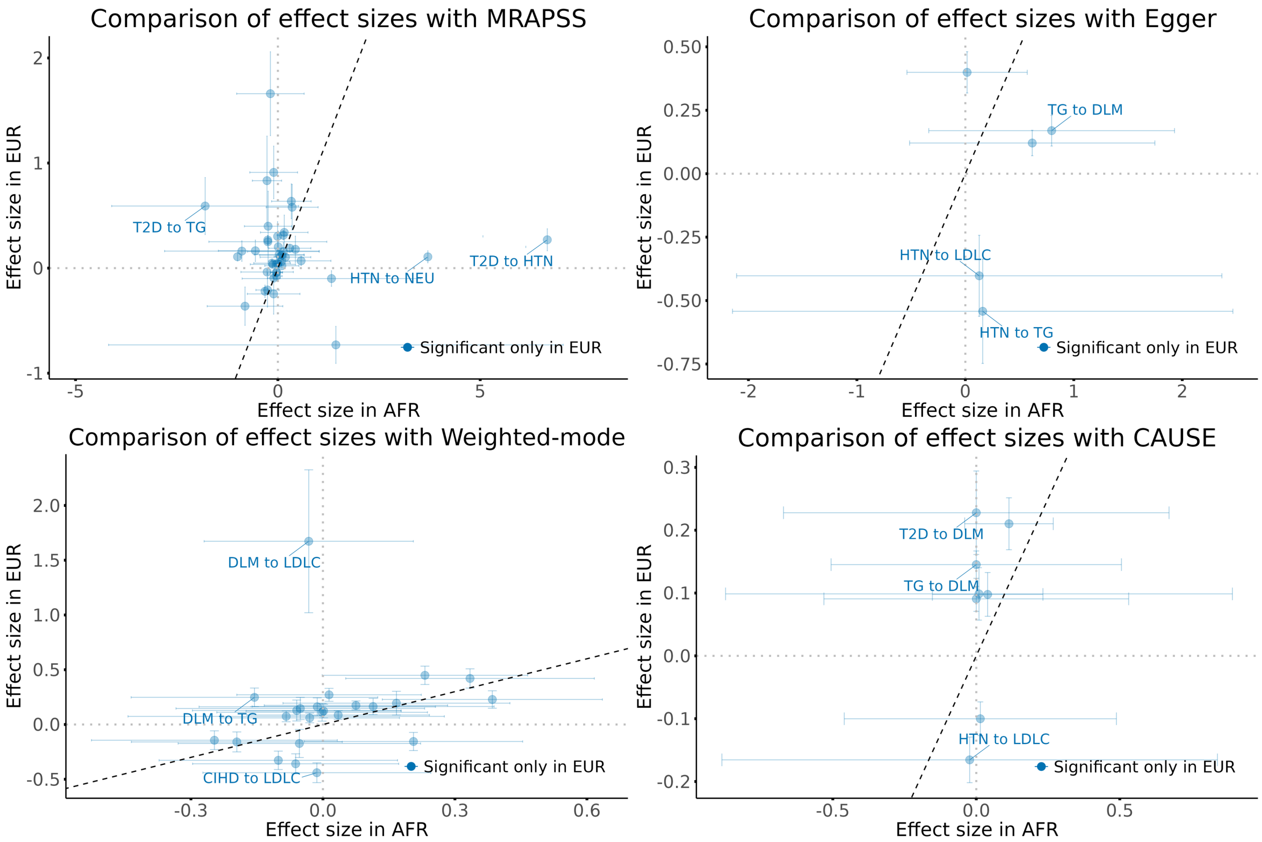

Figure S19: **Comparison of causal effect estimates between AFR and EUR.** Scatter plots comparing effect sizes in the AFR cohort versus the EUR cohort for MRAPSS, Egger, Weighted-mode, and CAUSE. Error bars represent 95% confidence intervals. Point colors indicate significance status in the two populations. The black dashed line represents the identity line ($y = x$).

#### 4.9 Sensitivity analysis of CAUSE with the default IV selection threshold $P \leq 1 \times 10^{-3}$

This section evaluates the sensitivity of CAUSE to its IV selection threshold by comparing the default threshold ($P \leq 1 \times 10^{-3}$) with the stricter threshold ($P \leq 5 \times 10^{-5}$) adopted in our main analyses.

In the BBJ cohort, where sample sizes generally exceed 100,000, the default threshold yields results comparable to the stricter threshold (Fig. S20). However, in the CSA and AFR populations, where sample sizes are typically below 10,000, the default threshold produces unstable estimates: a substantial fraction cluster near zero yet attain significant $P$ values, suggesting numerical instability rather than genuine causal signals (Fig. S21). This motivated our adoption of the stricter threshold for the primary analyses, while we systematically compare both versions across all experiments. Detailed causal effect estimates from both CAUSE versions are provided in Supplementary Tables S16–S19, S21–S22, and S24–S25.

In simulation studies (Figs. S22–S25; see also Supplementary Tables S8 and S13), both versions demonstrated well-controlled type I error rates under the null hypothesis. However, CAUSE (5e-5) exhibited slightly higher statistical power. Although the relaxed threshold (1e-3) incorporates more IVs—thereby reducing standard errors—it tends to underestimate causal effects more severely, likely because the additional weak instruments introduce bias. Real-data negative control results under three thresholds near $1 \times 10^{-3}$ (Fig. S26) further support the validity of using CAUSE (5e-5) as the primary analysis threshold.

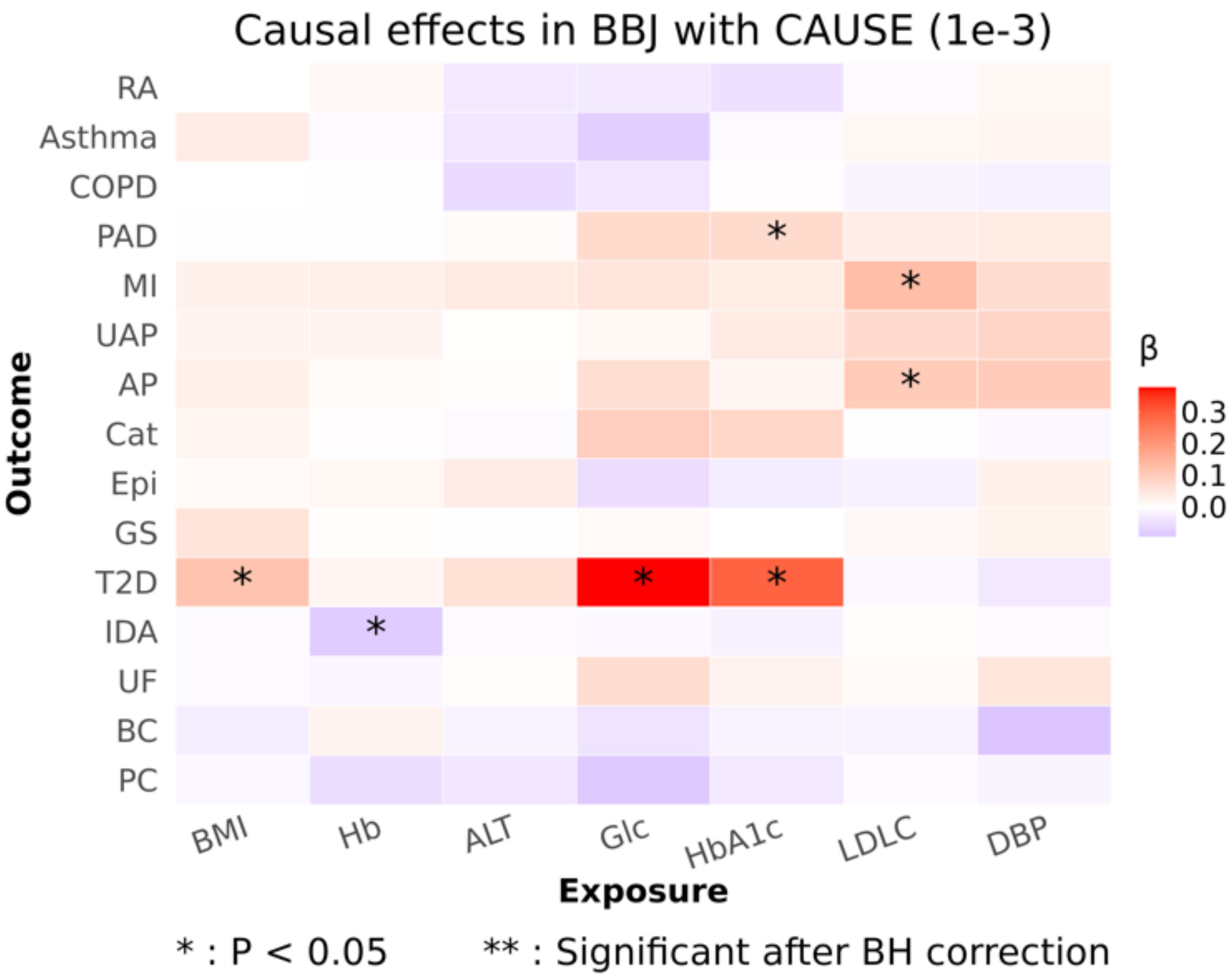

Figure S20: **Causal effect estimates in BBJ.** Heatmap displaying causal effect estimates ($\beta$) from CAUSE in the BBJ cohort using the default IV selection threshold ($P \leq 1 \times 10^{-3}$). Red indicates positive effects, and purple indicates negative effects. Statistical significance is denoted by * ($P < 0.05$) and ** (significant after BH correction).

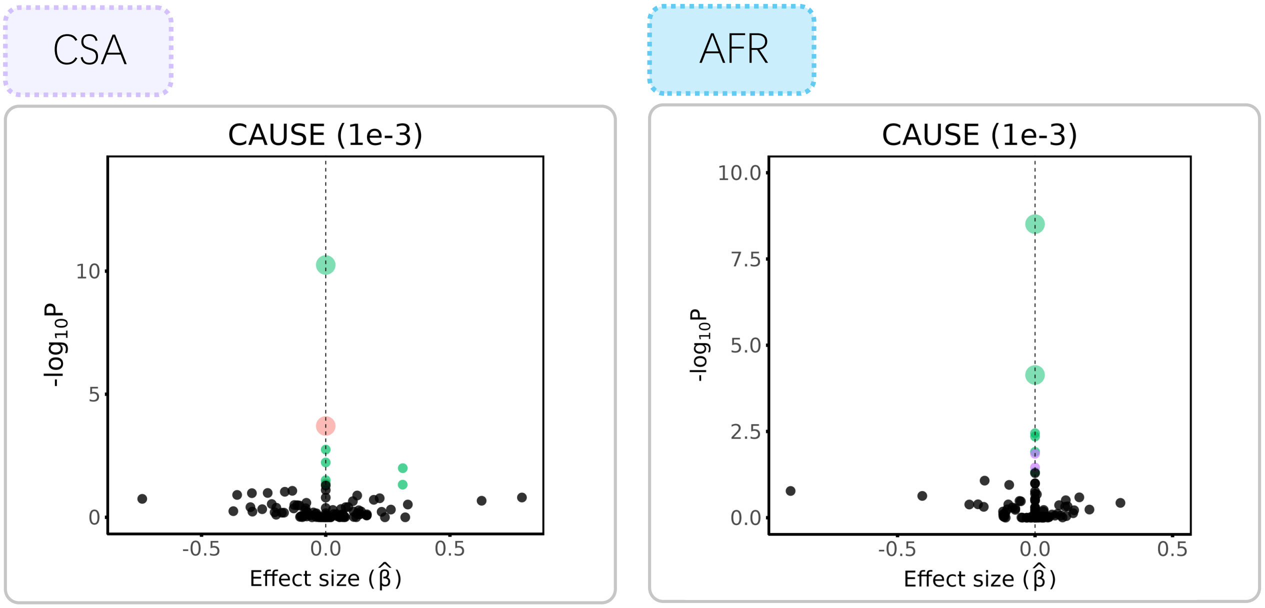

Figure S21: **Causal effect estimates in CSA and AFR.** Scatter plots showing the estimated effect size ($\hat{\beta}$) against the $-\log_{10}(p)$ value of each trait pair in CSA and AFR, using CAUSE with the default IV selection threshold ($P \leq 1 \times 10^{-3}$). Points with nominal significance ($P < 0.05$) are colored by exposure category, with shapes indicating significance after BH correction.

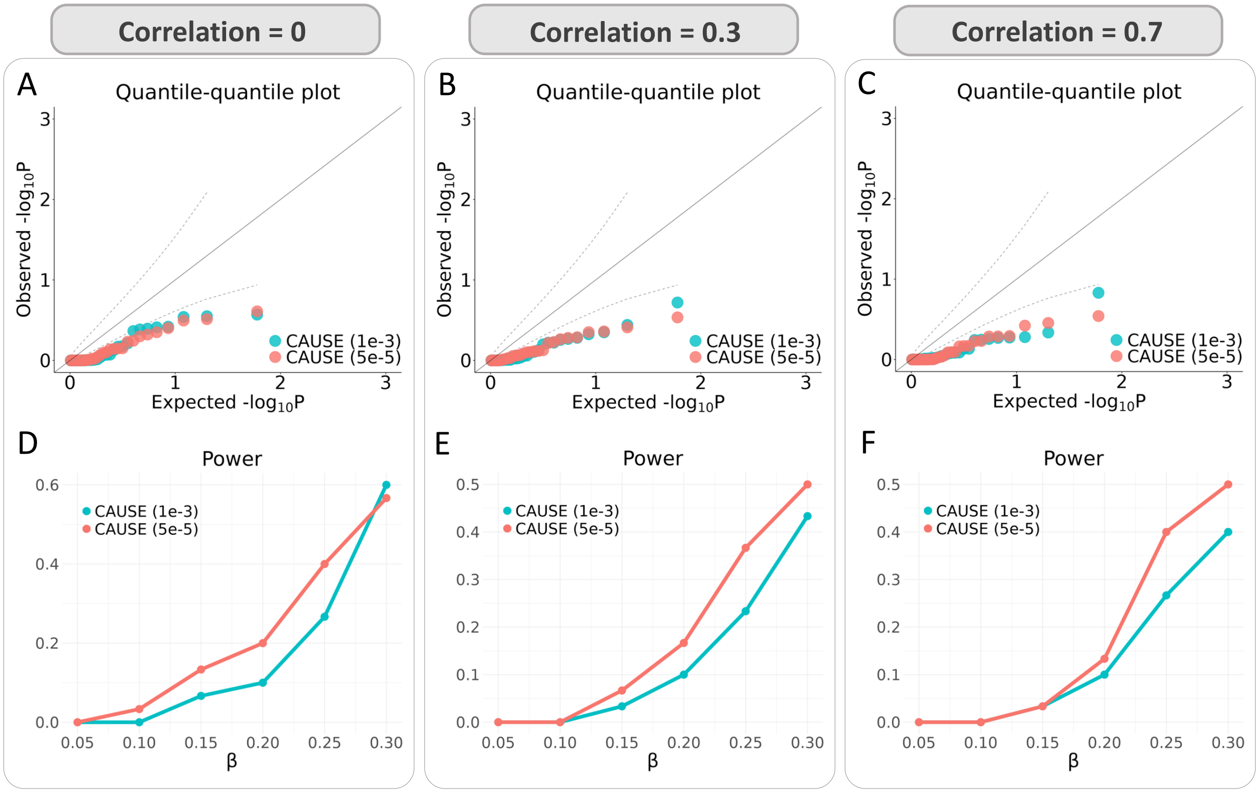

Figure S22: **Simulation performance of CAUSE using different thresholds. (A–C)** QQ plots of $-\log_{10}(p)$ values under the null scenario ($\beta = 0$, no causal effect). Results are shown from CAUSE with thresholds $P \leq 1 \times 10^{-3}$ (CAUSE 1e-3) and $P \leq 5 \times 10^{-5}$ (CAUSE 5e-5), with genetic correlation $\rho$ between populations set to 0, 0.3, and 0.7, respectively. **(D–F)** Comparison of statistical power between the two CAUSE versions under alternative simulations with varying causal effect sizes ($\beta$). Genetic correlation $\rho$ corresponds to 0, 0.3, and 0.7, respectively.

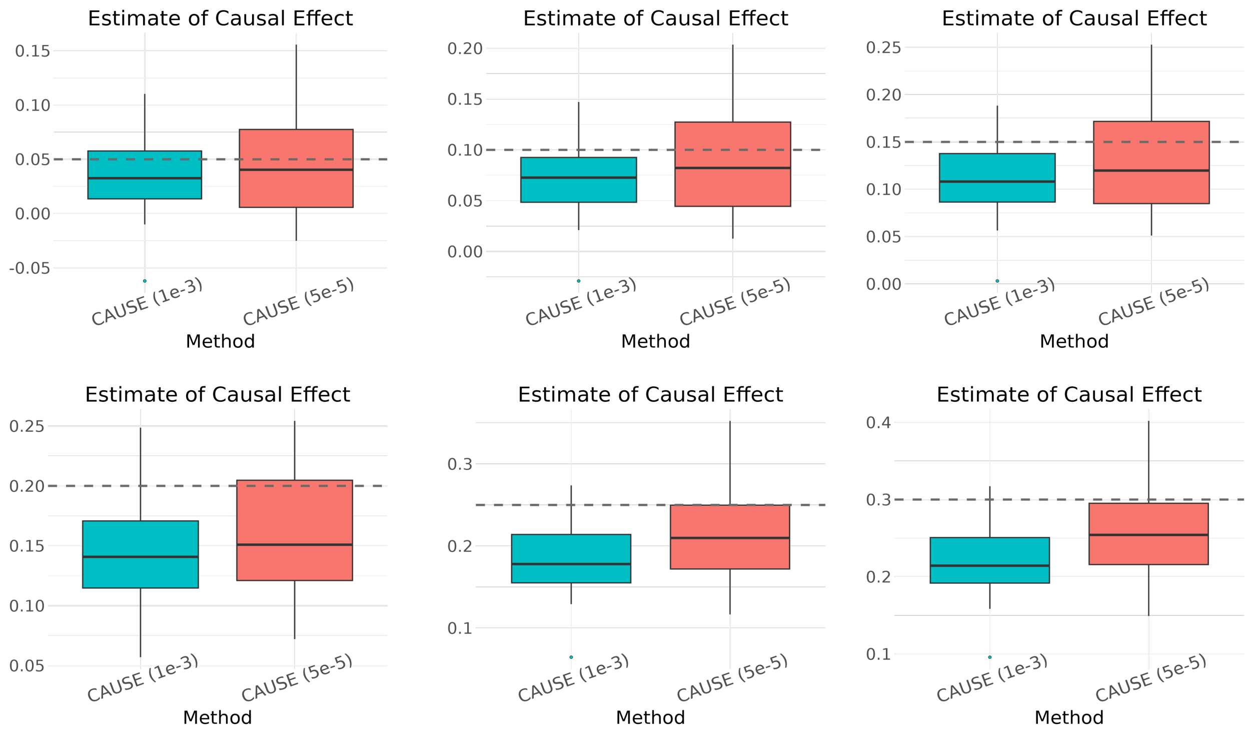

Figure S23: **Causal effect estimates under genetic correlation $\rho = 0$.** Boxplots comparing estimates from the two versions of CAUSE across 30 simulation replicates. The dashed lines represent the true causal effect size ($\beta$), ranging from 0.05 to 0.3.

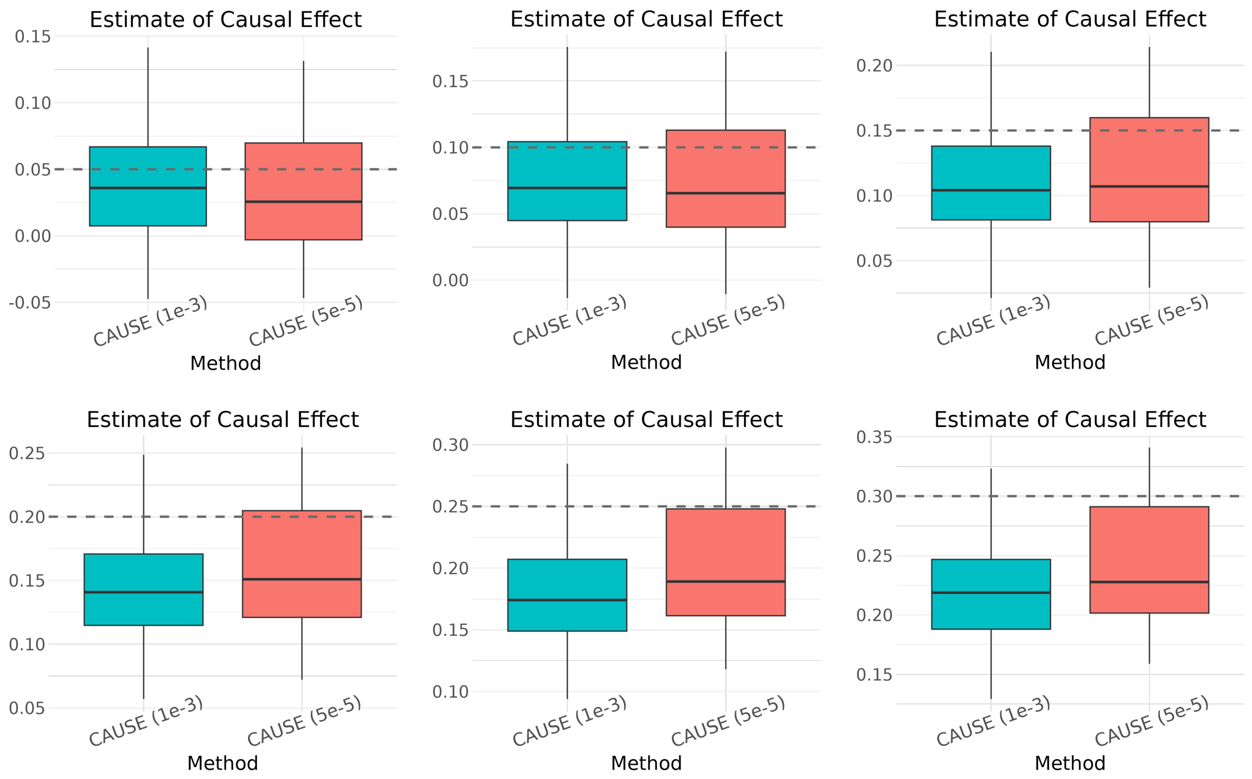

Figure S24: **Causal effect estimates under genetic correlation $\rho = 0.3$.** Boxplots comparing estimates from the two versions of CAUSE across 30 simulation replicates. The dashed lines represent the true causal effect size ($\beta$), ranging from 0.05 to 0.3.

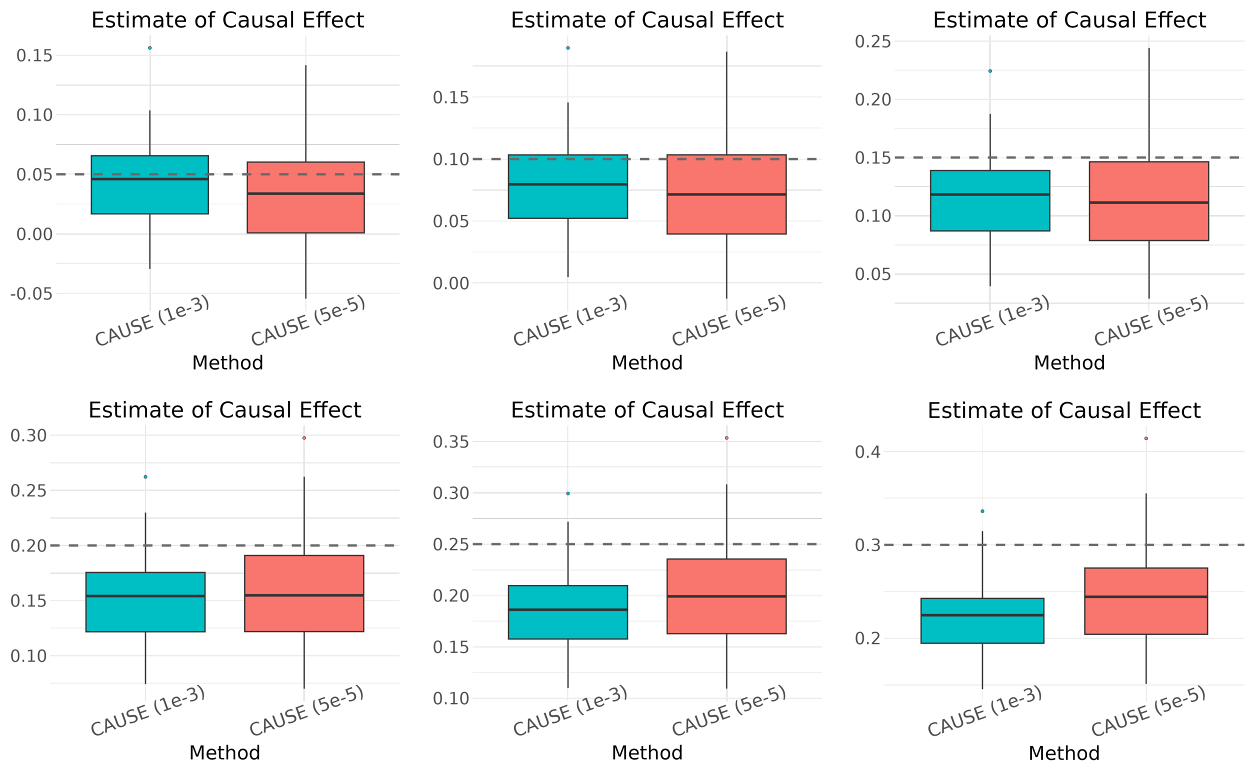

Figure S25: **Causal effect estimates under genetic correlation $\rho = 0.7$.** Boxplots comparing estimates from the two versions of CAUSE across 30 simulation replicates. The dashed lines represent the true causal effect size ($\beta$), ranging from 0.05 to 0.3.

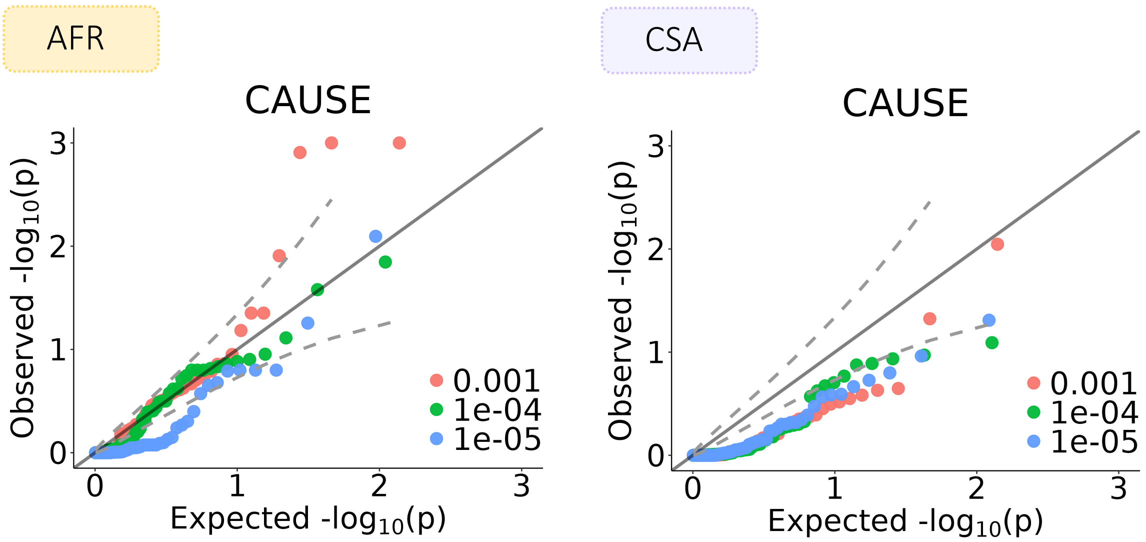

Figure S26: **Negative control studies for CAUSE in AFR and CSA.** QQ plots of $-\log_{10}(p)$ values derived from CAUSE applied to real-data negative controls, using three different IV selection thresholds near the default value.

#### 4.10 Distinctions between XMR and MRAPSS

To rigorously assess the contribution of XMR's key modeling innovations relative to MRAPSS, we designed an ablation study examining three simplified variants of XMR, each targeting a specific component that differentiates the two methods.

The first variant, **XMR** ($\sigma_{12,f} = 0$), evaluates the necessity of explicitly modeling the genetic correlation between the target and auxiliary populations. This variant fixes $\sigma_{12,f} = 0$ during the estimation of $\mathbf{\Sigma}$, effectively ignoring the shared genetic architecture between populations.

The second and third variants address XMR's correction term $\Delta t$, which accounts for selection bias arising from LD clumping across populations. Instead of the principled correction used in XMR, these variants adopt a simpler threshold adjustment analogous to that used in MRAPSS:

$$P \text{ value threshold} \leftarrow \text{IV threshold} \times \min\left(\frac{\text{median}(P \text{ values}_{\text{after}})}{\text{median}(P \text{ values}_{\text{before}})}, 1\right), \quad \text{(S38)}$$

where $P$ values$_{\text{before}}$ and $P$ values$_{\text{after}}$ represent the $P$ values of IVs before and after LD clumping, respectively. **XMR ($\Delta t$ unmodeled 1)** computes the median $P$ value ratio from the auxiliary population's IVs, while **XMR ($\Delta t$ unmodeled 2)** computes it from the target population's IVs.

As illustrated in Fig. S27, both $\Delta t$ variants failed to maintain well-calibrated $P$ values in the real-data negative control study. This inflation underscores the critical role of XMR's principled modeling of $\Delta t$ in correcting selection bias; the simple threshold adjustment used in MRAPSS does not adequately capture this effect in the cross-population setting.

The XMR ($\sigma_{12,f} = 0$) variant maintained reasonable false positive rates but suffered from reduced statistical power compared to the full XMR model (Fig. S28). When the true genetic correlation was null ($\rho = 0$), this variant and the full model yielded nearly identical results (Figs. S28 and S29), confirming that the full model correctly adapts when no cross-population correlation exists. However, when a true genetic correlation was present ($\rho > 0$), XMR ($\sigma_{12,f} = 0$) systematically underestimated the causal effect $\beta$ (Figs. S30 and S31). This downward bias directly explains the observed loss of power and demonstrates that leveraging the shared genetic architecture between populations through $\sigma_{12,f}$ is essential for accurate causal estimation in cross-population MR.

Detailed causal effect estimates across these three XMR variants in the real-data negative control study are presented in Supplementary Tables S9 and S14.

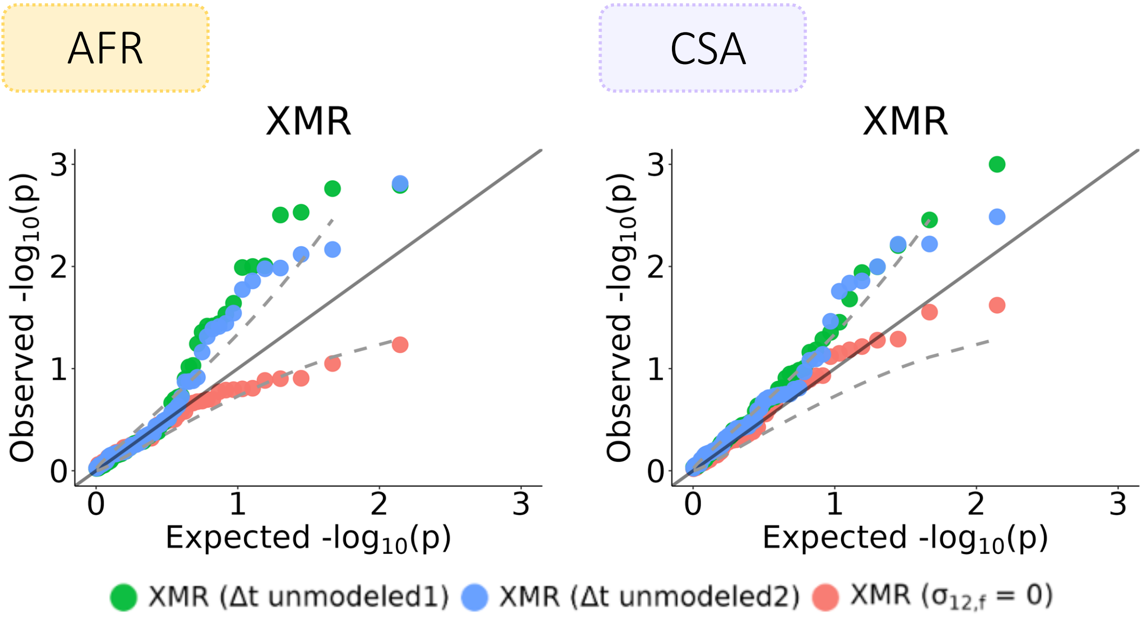

Figure S27: **XMR variants in real-data negative control studies.** The distribution of $-\log_{10}(p)$ values is shown from three variants of the XMR model in both AFR (left) and CSA (right) populations.

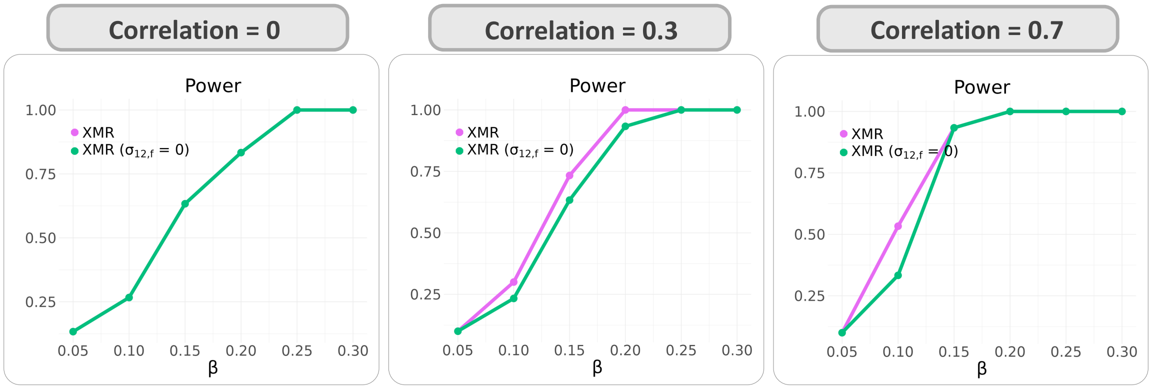

Figure S28: **Power comparison in simulations.** QQ plots of $-\log_{10}(p)$ values from 30 independent simulation experiments comparing the full XMR model against the XMR ($\sigma_{12,f} = 0$) variant. The genetic correlation parameter $\rho$ between the two populations varies from 0 to 0.7.

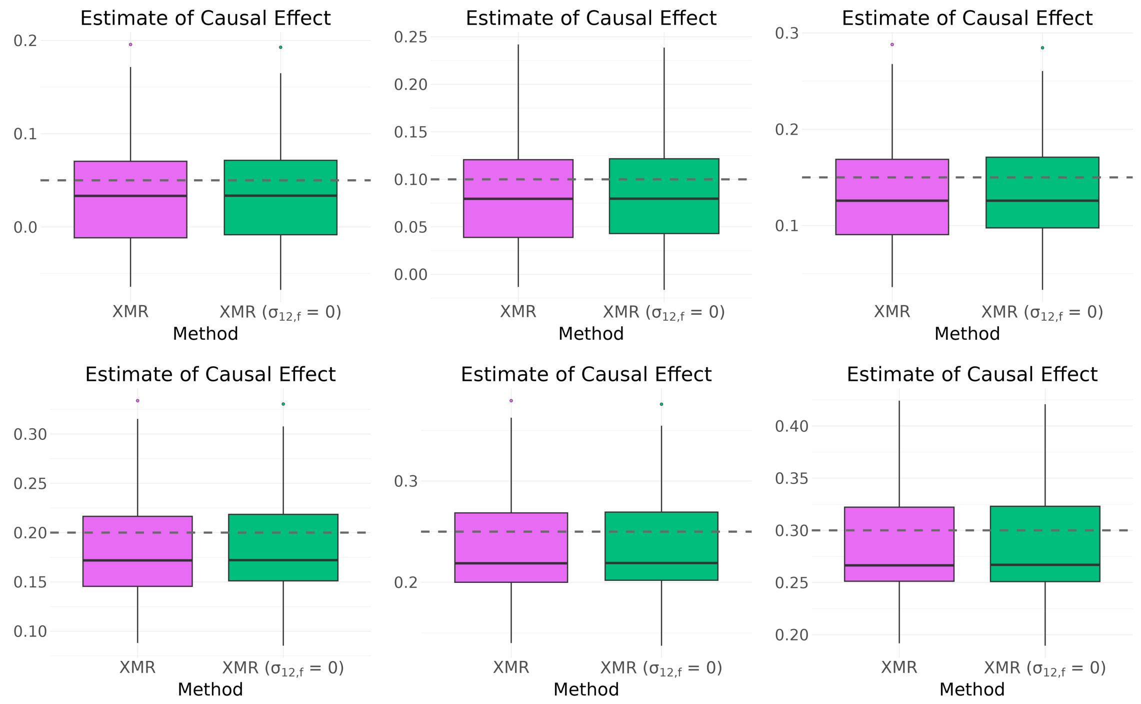

Figure S29: **Causal effect estimates under genetic correlation $\rho = 0$.** Boxplots representing the distribution of estimates from 30 independent simulation experiments by XMR and its variant with $\sigma_{12,f} = 0$. The dashed horizontal lines indicate the true causal effect size $\beta \in \{0.05, 0.1, 0.15, 0.2, 0.25, 0.3\}$.

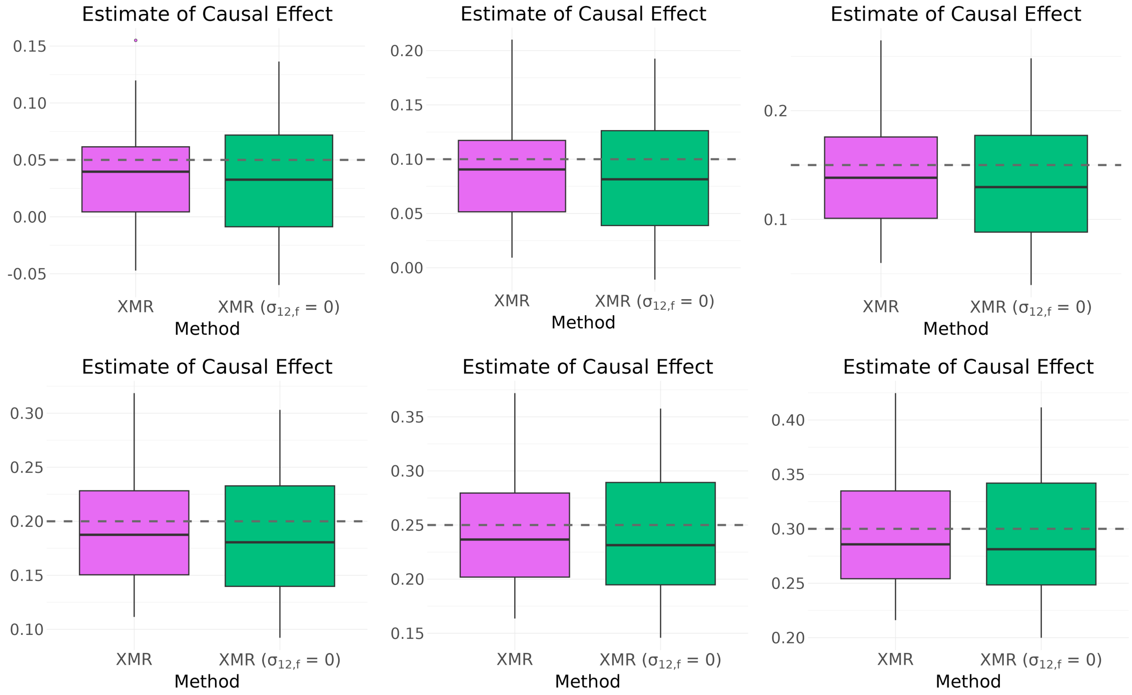

Figure S30: **Causal effect estimates under genetic correlation** $\rho = 0.3$**.** Boxplots representing the distribution of estimates from 30 independent simulation experiments by XMR and its variant with $\sigma_{12,f} = 0$. The dashed horizontal lines indicate the true causal effect size $\beta \in \{0.05, 0.1, 0.15, 0.2, 0.25, 0.3\}$.

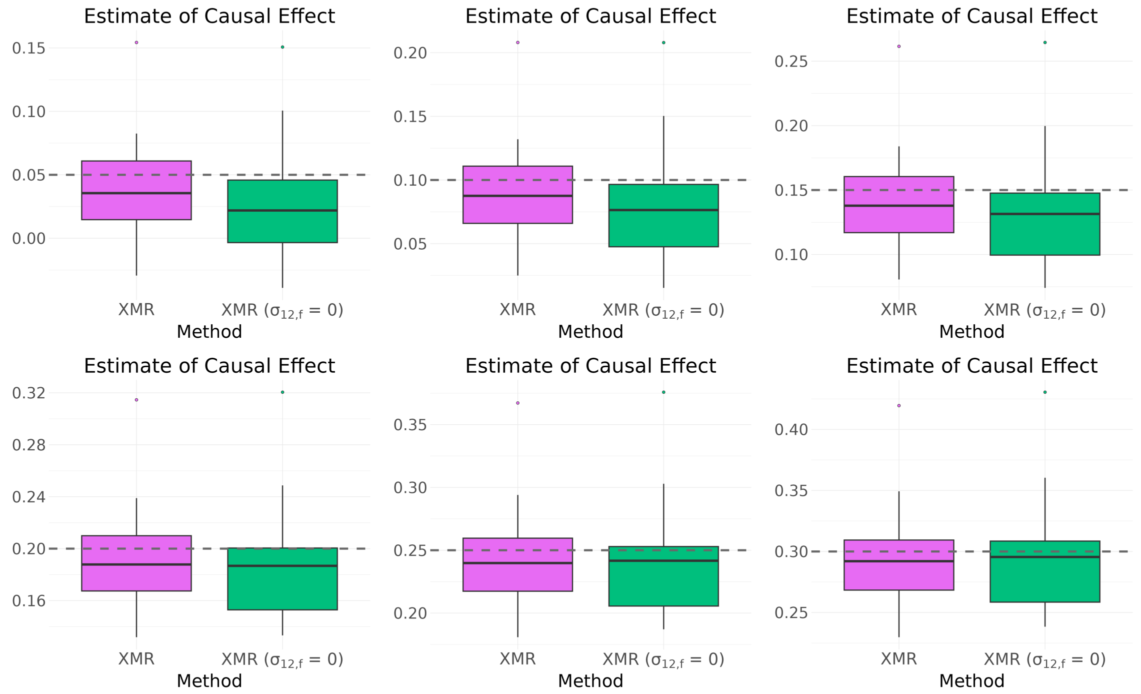

Figure S31: **Causal effect estimates under genetic correlation $\rho = 0.7$.** Boxplots representing the distribution of estimates from 30 independent simulation experiments by XMR and its variant with $\sigma_{12,f} = 0$. The dashed horizontal lines indicate the true causal effect size $\beta \in \{0.05, 0.1, 0.15, 0.2, 0.25, 0.3\}$.
